# Stroke Burden Attributable to Risk Factors in the Americas, 1990–2021: A Temporal Trends Analysis from the Global Burden of Disease Study 2021

**DOI:** 10.1101/2025.07.10.25331329

**Authors:** Felipe Fregni, Bassel Almarie, Ramón Martinez, Verónica V. Olavarría, Carlos Abanto, Matias Alet, Tony Fabián Alvarez, Pablo Amaya, Sebastian Ameriso, Antonio Arauz, Miguel A. Barboza, Hernán Bayona, Antonio Bernabé-Ortiz, Juan Calleja, Vanessa Cano-Nigenda, Leonardo Augusto Carbonera, Rodrigo M. Carrillo-Larco, Angel Corredor, Ana Cláudia de Souza, Jose D Diestro, Rodrigo Guerrero, Claudio Jimenez, Fernando Lanas, Sheila Martins, Paula Muñoz-Venturelli, Victor Navia, Nelson Novarro-Escudero, Bruce Ovbiagele, Octavio Pontes-Neto, Pedro Ordunez, Virginia Pujol, Alejandro A. Rabinstein, Julieta Rosales, Andres Rosende, Gisele Sampaio Silva, Gustavo Saposnik, Souvik Sen, Luciano Sposato, Fernando D. Testai, Victor Urrutia, Craig S. Anderson, Pablo M. Lavados

## Abstract

**Background:** Stroke remains a major public health concern with marked disparities across populations, particularly in the Americas. Despite documented declines in overall burden, limited data exist on how modifiable risk factors contribute to contemporary stroke trends across the continent.

**Methods:** Using data from the Global Burden of Disease 2021 study, we analyzed age-standardized mortality rates and age-standardized disability-adjusted life-years (DALYs) rates from stroke attributable to 23 modifiable risk factors across 39 countries and territories in the Americas. Estimates were stratified by age, sex, stroke subtype, and Socio-Demographic Index (SDI). Temporal trends from 1990 to 2021 were assessed using the average annual percent change (AAPC), which was estimated applying Joinpoint regression analysis.

**Results:** In 2021, 78% of stroke deaths and 77% of stroke DALYs were attributable to modifiable risk factors in the Americas. High systolic blood pressure was the leading risk factor (20.87 deaths [95% uncertainty interval: 15.17–25.8] and 453.30 DALYs [333.16–556.41] per 100,000), contributing over half of stroke burden. From 1990–2021, the largest reductions occurred in household air pollution (deaths AAPC −5.09% [−5.17–−5.02]; DALYs AAPC −4.92% [−4.98–−4.86]) and secondhand smoke (deaths AAPC −3.45% [−3.49–−3.41]; DALYs AAPC −3.37% [−3.41–−3.33]). In contrast, several risk factors showed minimal progress: high body-mass index (deaths AAPC −0.54% [−0.59–−0.49]; DALYs AAPC −0.42% [−0.46–−0.37]) and high fasting glucose (deaths AAPC −1.10% [−1.14–−1.06]; DALYs AAPC −0.99% [−1.05–−0.93]). High temperature demonstrated increases, with AAPCs exceeding +3.5% in select countries. High systolic blood pressure declined moderately (deaths AAPC −1.89% [−1.93–−1.85]; DALYs AAPC −1.84% [−1.89–−1.80]), with stroke burden remaining disproportionately higher in low-SDI countries.

**Conclusion:** Although encouraging declines in stroke burden attributable to risk factors have been observed over three decades, substantial variation persists by geography and SDI. High systolic blood pressure remains the leading modifiable risk factor. Minimal progress in obesity and metabolic factors highlights the need for equity-focused strategies to address persistent and emerging stroke risks across the Americas.

## Introduction

Stroke remains a leading cause of disability and mortality worldwide, with substantial regional variations. In 2021, stroke accounted for 7.3 million deaths (10.7% of all deaths) globally, making it the third most common cause of death. It also contributed to 160.5 million disability-adjusted life years (DALYs), or 5.6% of total DALYs.^1^ In the Americas, stroke was responsible for 0.5 million deaths (38.1 deaths per 100 000 population), and 11.4 million DALYs or 869.2 DALYs per 100 000 population in 2021.^2^ The burden of stroke and its subtypes varies substantially across countries and stroke subtypes in the Americas. ^4^

A high proportion of this burden is attributable to behavioral, metabolic, dietary, and environmental risk factors, making stroke a target for prevention-oriented public health strategies.^1,3^ Several factors contribute to stroke. Previous studies underscore the impact of socioeconomic status on stroke, showing an inverse relationship between income and stroke risk.^5^ Another study, focusing on countries in the Americas, showed an inverse association between population-level hypertension control and stroke mortality, highlighting how many stroke deaths could be prevented for each unit increase in population-level hypertension control.^6^ Critical questions persist regarding the timing and relative contribution of modifiable risk factors for stroke, as well as the impact of public health efforts aimed at modifying behaviour (e.g., smoking cessation, decreasing alcohol use), and reducing environmental (e.g., air pollution), dietary (e.g., high sodium intake), and metabolic (e.g., high systolic blood pressure, high low density lipoprotein [LDL]-cholesterol) contributors. To date, no study has systematically and comprehensively quantified long-term trends in stroke burden attributable to risk factors across the Americas at this level of geographic and risk-specific resolution.

This study aimed to assess the levels and temporal trends of mortality and DALYs due to stroke and stroke major types attributable to risk factors by sex, age, and socio-demographic index (SDI) in the Americas from 1990 to 2021. It includes analysis at regional and national (38 countries and territories) levels.

## Methods

### Study Design, Setting, and Data Sources

The study investigates the mortality and disease burden―measured in DALYs―due to stroke, and stroke type (ischemic stroke, intracerebral haemorrhage, and subarachnoid haemorrhage) attributable to 23 risk factors and six risk factor clusters by age, sex at the population level in the Region of the Americas from 1990 to 2021, using data from the Global Burden of Disease (GBD) Study 2021.

According to the GBD location hierarchy, the Americas comprise the super-region Latin America and Caribbean, which is further subdivided into four regions (Andean Latin America, Central Latin America, Caribbean, and Tropical Latin America) and two additional regions (High-income North America, and Southern Latin America).^7^ The Americas encompasses 39 countries and territories, with a combined population exceeding two billion.^8,9^ (appendix pp xx) All the data used in this article are freely accessible after registration via the GBD Results Tool (https://vizhub.healthdata.org/gbd-results/). As all the data included were aggregated and not identifiable, this study was exempt from institutional review board evaluation.

### Analytical Approach

The methodology for estimating stroke mortality and DALYs attributable to risk factors in the GBD 2021 aligns with previously established methodologies.^1,10–12^ Stroke was categorised into three main subtypes: ischaemic stroke, intracerebral haemorrhage, and subarachnoid haemorrhage, following the clinical WHO criteria.^13^ Cause of Death Ensemble Modelling (CODEm) was used to estimate death-related stroke, while the DisMod-MR 2.1 Bayesian tool was used for non-fatal disease parameters, such as incidence and prevalence. Stroke burden estimates are presented as age-standardised rates per 100,000 population, stratified by sex and geographic location within the Americas. Additionally, burden estimates were stratified by age group from 25 to 94 at five-year intervals and 95+.

We estimated the population attributable fraction (PAF) to quantify the proportion of stroke deaths and DALYs attributable to leading risk factors, using standard GBD comparative risk assessment methods, as described elsewhere.^10^ PAF was calculated by integrating risk factor exposure levels, relative risks from meta-analyses, and theoretical minimum risk exposure levels.^14^ PAF estimates were generated separately for deaths and DALYs and stratified by geographic region to assess variations in risk factor contributions across the Americas.

Summary Exposure Values (SEVs) were also used to visualise population-level exposure to each risk factor. SEV is a GBD metric ranging from 0% (no excess risk) to 100% (entire population exposed to the maximum level of risk), reflecting both the prevalence of exposure and the magnitude of risk associated with each level of exposure. SEV values for each risk factor were obtained from the GBD 2021 results and incorporated into our visualizations. Detailed methods for SEV calculation are published elsewhere.^15^

### Attributable Risk Factors

Our analysis evaluated 23 risk factors grouped into four clusters: Environmental, dietary, behavioral, and metabolic. Environmental risks included ambient particulate matter pollution, household air pollution from solid fuels, low ambient temperature, high ambient temperature, and lead exposure. Dietary risks included high sodium intake, high red meat consumption, high processed meat consumption, low fruit intake, low vegetable intake, low whole grain intake, high sugar-sweetened beverage consumption, low fiber intake, and low omega-6 polyunsaturated fatty acid intake.

Behavioral risks included low physical activity, alcohol use, smoking, and second-hand smoke exposure. Metabolic risks included high body mass index, high fasting plasma glucose, high systolic blood pressure, high LDL cholesterol, and kidney dysfunction.

### Burden According to Socio-Demographic Index (SDI)

Countries and territories in the Americas were grouped by SDI levels. The SDI is a composite measure of income, education, and fertility rates, providing a standardized development assessment across locations.^16^ The SDI measure ranges from 0 to 1. Higher values indicate higher levels of development. The SDI scale is divided into five categories: low (<0.45), low-middle (≥0.45 to <0.60), middle (≥0.60 to <0.75), high-middle (≥0.75 to <0.85), and high (≥0.85).

Based on this classification, regions were assigned to SDI levels as follows: high SDI included high-income North America (mean SDI = 0.864); high-middle SDI included Southern Latin America (SDI = 0.743); and middle SDI included the Caribbean (0.642), Andean Latin America (0.654), Central Latin America (0.642), and Tropical Latin America (0.649).

Within each SDI category, countries and territories were grouped based on their 2021 SDI values. The high SDI group, comprising approximately 372 million people (36.5% of the regional population), included Canada, the United States of America, Bermuda, Greenland, Puerto Rico, and the United States Virgin Islands (n = 6). The high-middle SDI group represented around 77 million people (7.6%) and included Argentina, Chile, Uruguay, Antigua and Barbuda, the Bahamas, Barbados, Dominica, Trinidad and Tobago, and Saint Kitts and Nevis (n = 9). The middle SDI category encompassed the largest population share, with approximately 512 million people (50.2%) across 16 countries, including Brazil, Cuba, Colombia, Costa Rica, the Dominican Republic, Ecuador, Jamaica, Mexico, Panama, Paraguay, Peru, and others. Countries classified as low-middle SDI—Belize, Bolivia (Plurinational State of), El Salvador, Guatemala, Honduras, Nicaragua, and Venezuela (n = 7)—accounted for approximately 47.4 million people (4.6%). Finally, Haiti was the only country in the low SDI category, representing 11 million people (1.1%).

The burden of stroke-related deaths and DALYs, presented for both sexes as age-standardised rates per 100,000, was stratified by SDI category across all locations and risk factors to identify patterns of stroke burden attributable to risk factors across varying levels of development. An inverse variance-weighted mean approach was applied to calculate aggregated prevalence rates for each classification. Weighted mean deaths and DALYs were computed with 95% uncertainty intervals and reported as age-standardized rates per 100,000 population.

### Temporal Trends

We analyzed temporal trends in stroke rates using a log-linear joinpoint regression model with year (1990-2021) as the independent variable and age-standardized stroke deaths or DALY rates (per 100,000) as the dependent variable. Negative or zero rates were excluded to allow log transformation of the dependent variable.

Models with zero to four joinpoints were tested, requiring a minimum of two years between joinpoints and at least two data points from each joinpoint to the ends of the time series. The Weighted Bayesian Information Criterion was used to guide selection of the optimal model, balancing fit with parsimony.

Only the Average Annual Percentage Change (AAPC) was presented to summarize overall trends. AAPC was calculated with 95% CI derived via the empirical quantile method with 5,000 resamples. If the AAPC is above 0 or below 0, and the p-value is less than 0.05, it suggests a statistically significant upward or downward trend, respectively.

Diagnostic outputs, including regression coefficients, covariance matrices, and model fit statistics were examined to ensure the robustness of the selected models. We set variance to be constant (homoscedasticity), and residuals uncorrelated as a result of the visual inspection of the plot of residuals against fitted values.

All temporal change analyses were performed in the Joinpoint Regression Program, version 5.2.0.0 (December 2024), developed by the National Cancer Institute (NCI), Bethesda, MD, USA. The statistical significance of joinpoints was evaluated using the Monte Carlo permutation test with a threshold of 0.05. Analyses were stratified by sex and geographic location, with each risk factor independently assessed. The AAPC was labelled as “not available” in the results table for excluded data.

All statistical analyses were conducted using R version 4.3.1 (2023-06-16) with (RStudio version 2023.06.1+524), with a significance level of 0.05 applied throughout. Data visualizations were performed using Python version 3.12.7 (JupyterLab version 4.2.5).

## Results

### 1. Burden of Stroke Attributable to Risk Factors

In the Americas, the total number of stroke DALYs due to risk factors increased by 0.16% (0.11 to 0.22) from 7.5 million (95% UI 8.0 to 6.9) in 1990 to 8.8 million (9.8 – 7.7) in 2021, but the corresponding age-standardized stroke DALYs attributable to risk factors declined by 4.8% [2.4 to 7.9) (Table 3).

In 2021, 78.1% (70.0 - 84.9) of deaths from stroke and 75.8 (67.3 - 82.4) of DALYs from stroke were attributable to 23 modifiable risk factors, with the largest proportions of attributable risks for total stroke observed in Southern Latin America, and Caribbean (Table 1, Supplementary Figure 56-69)

**Table 1:**
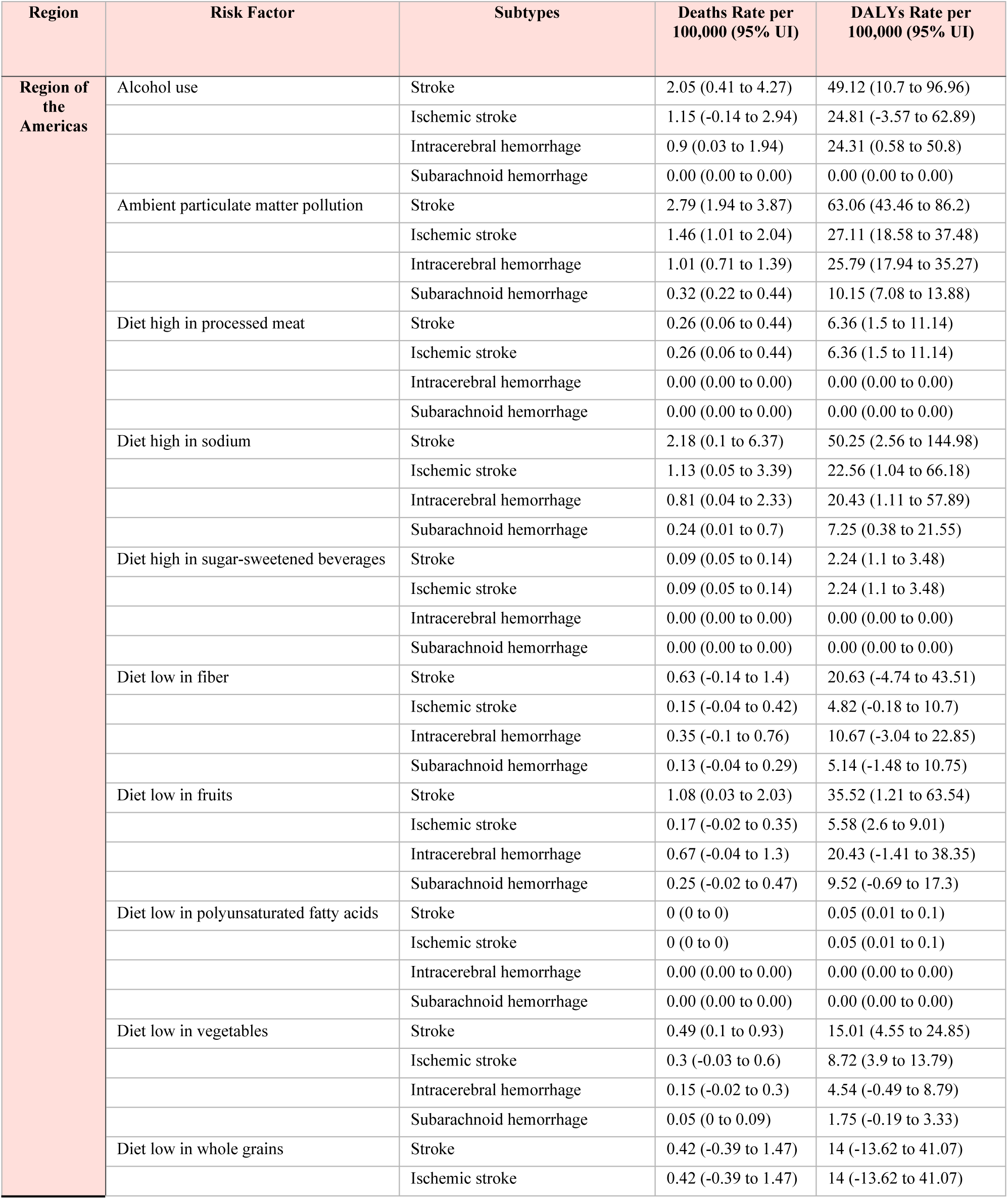
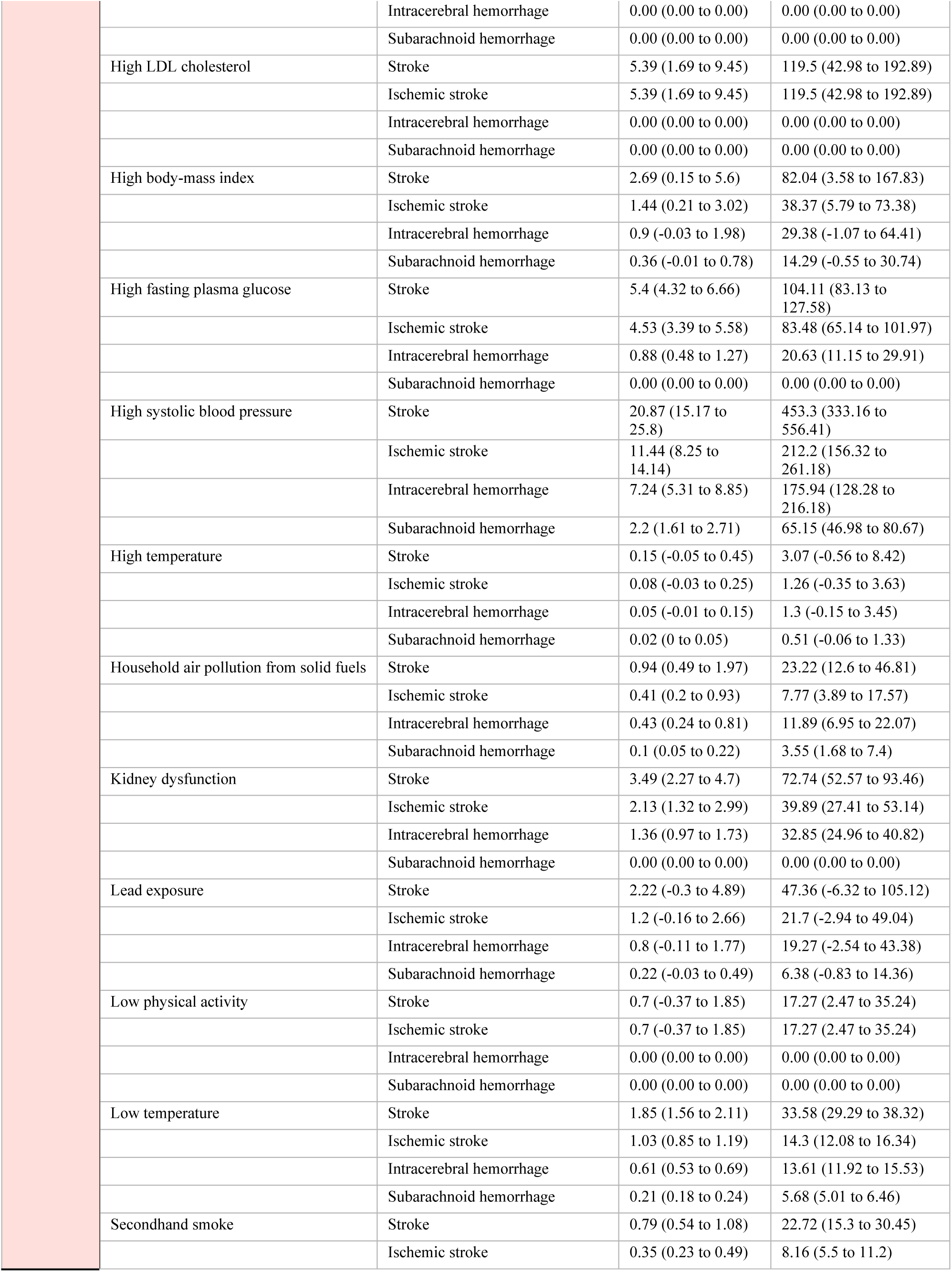
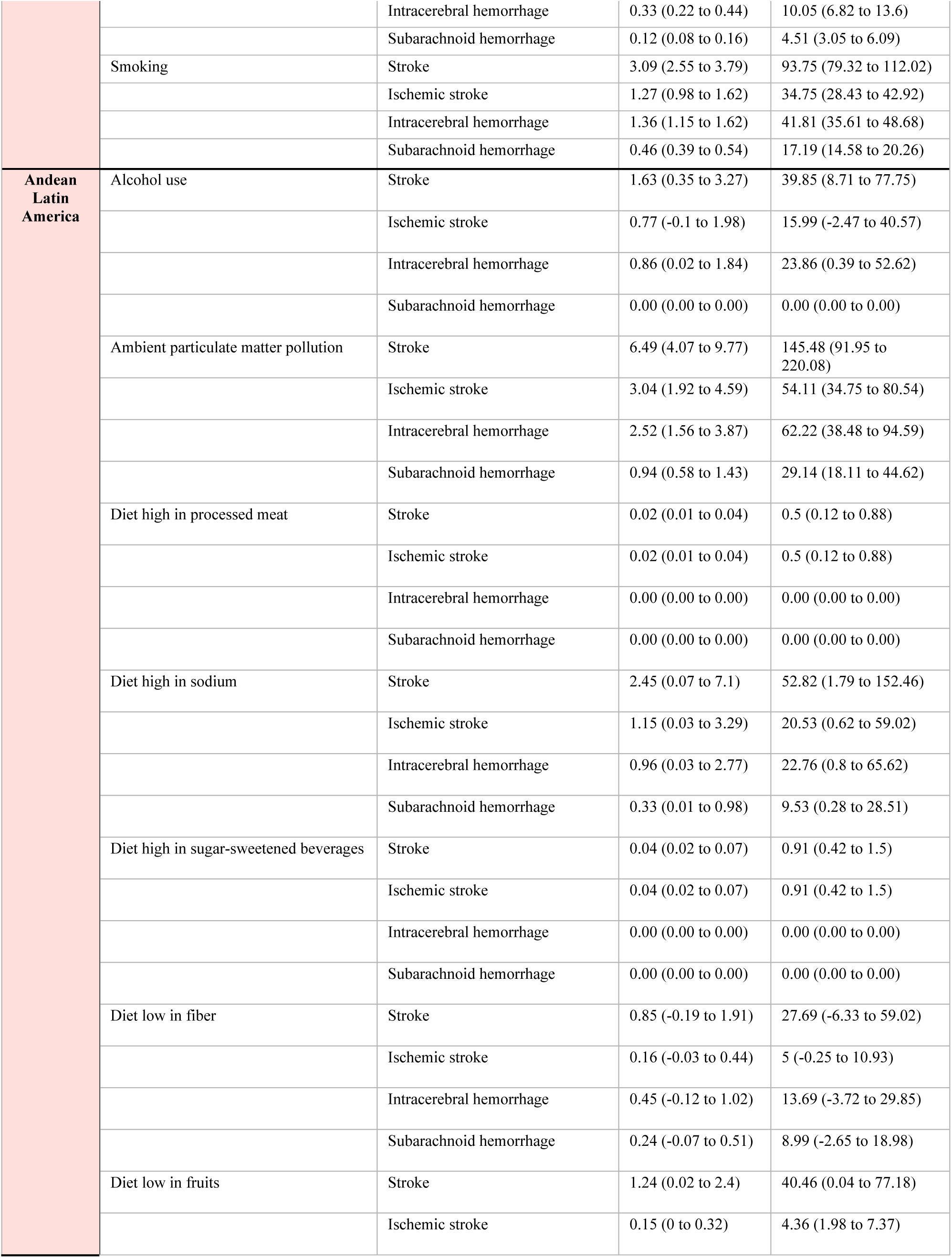
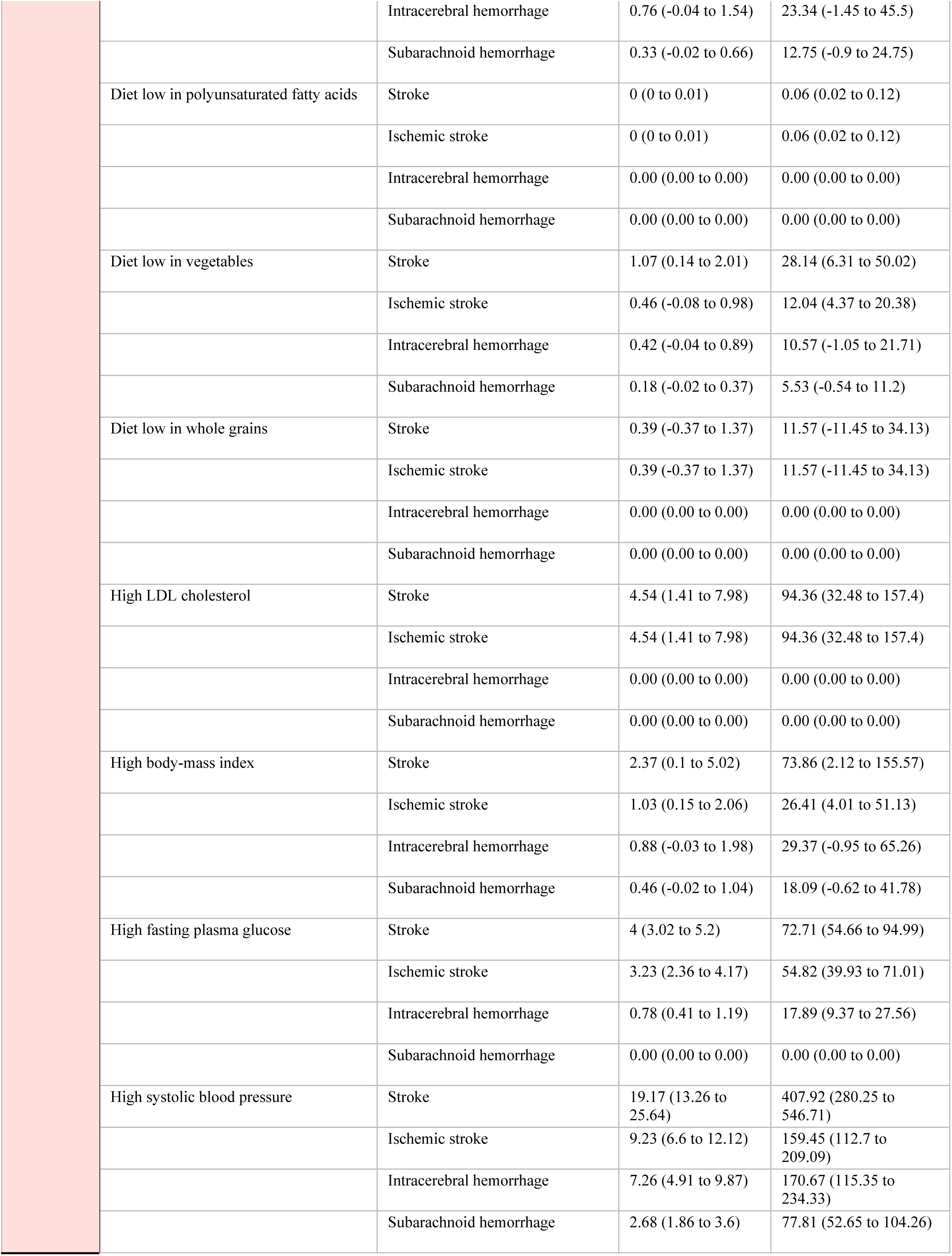
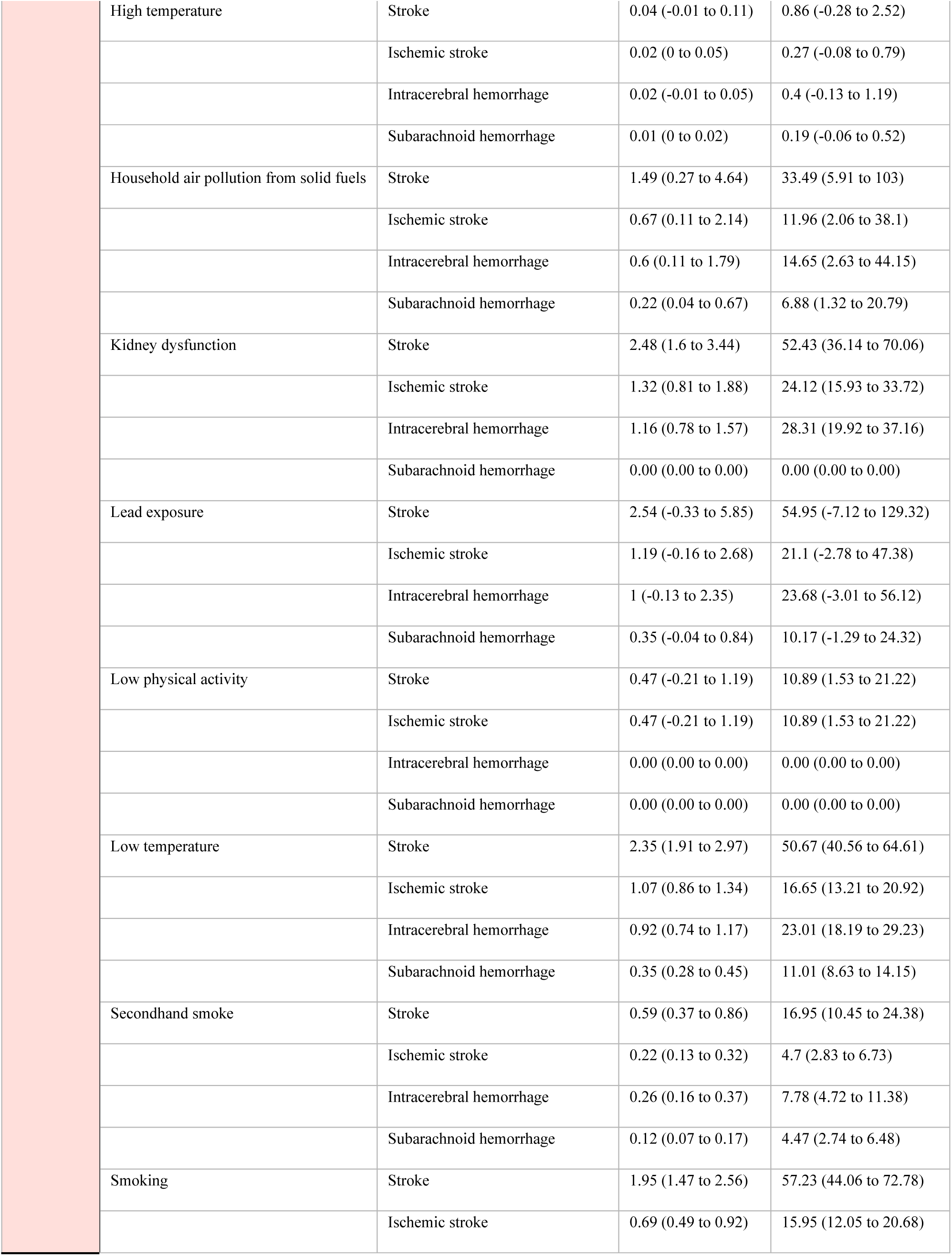
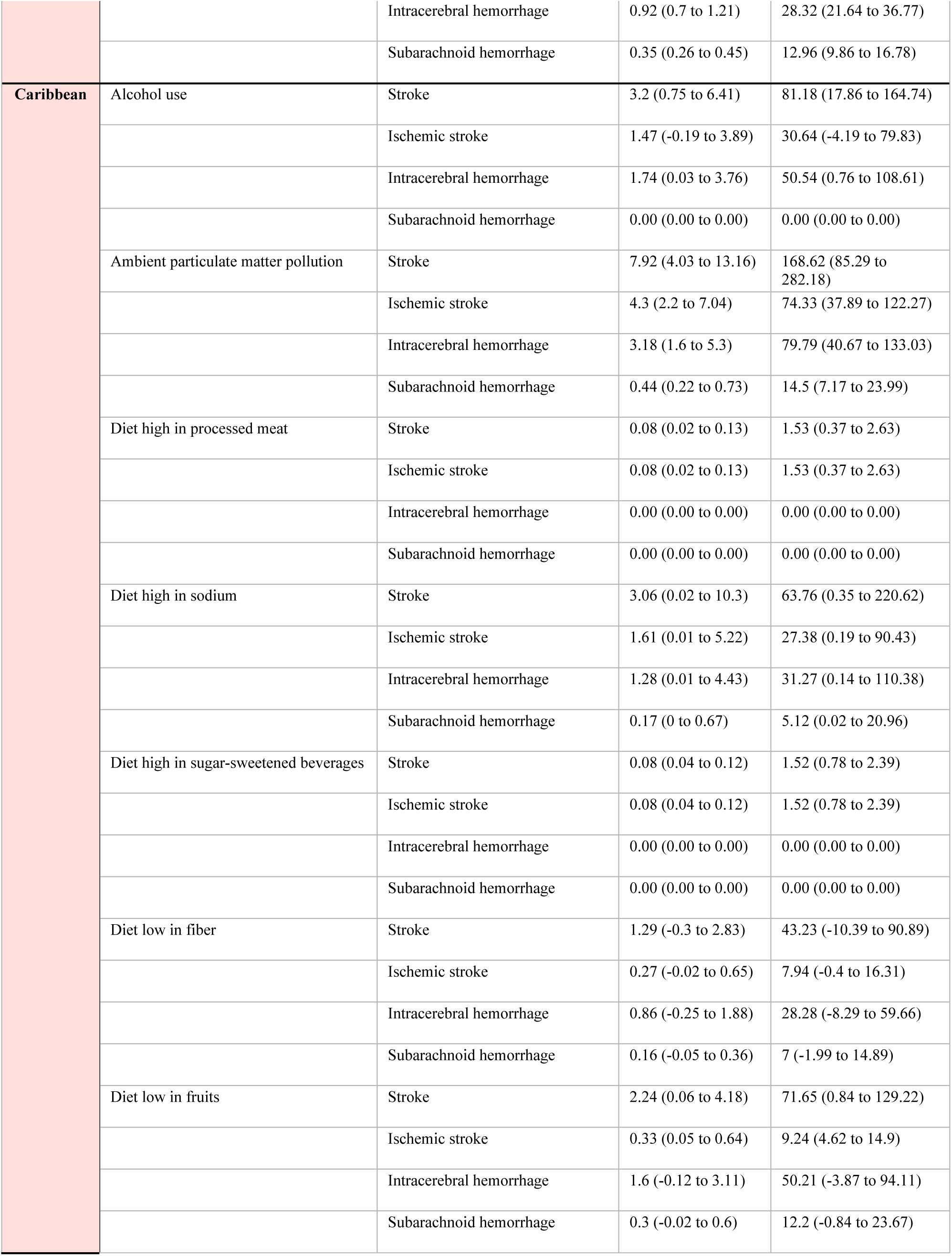
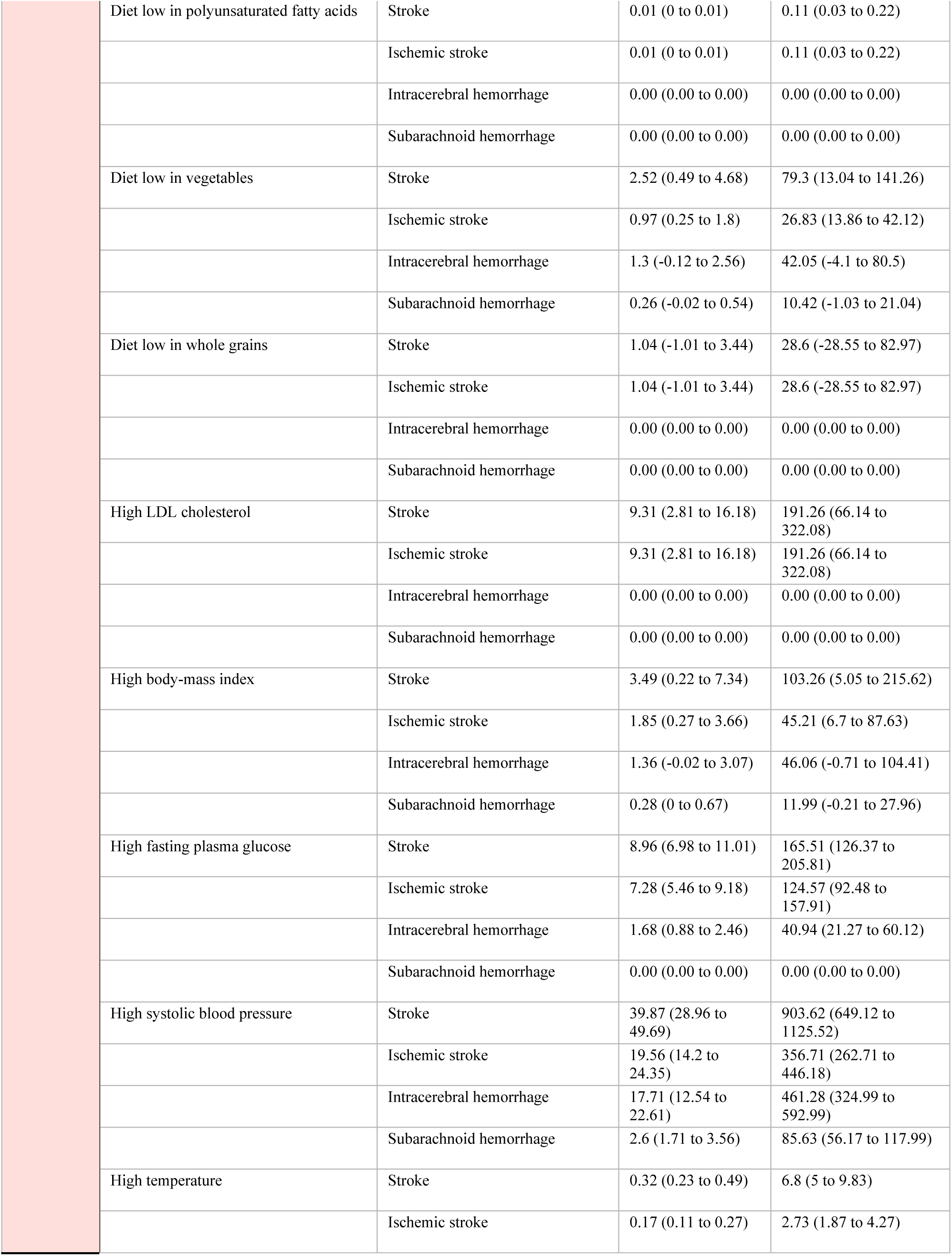
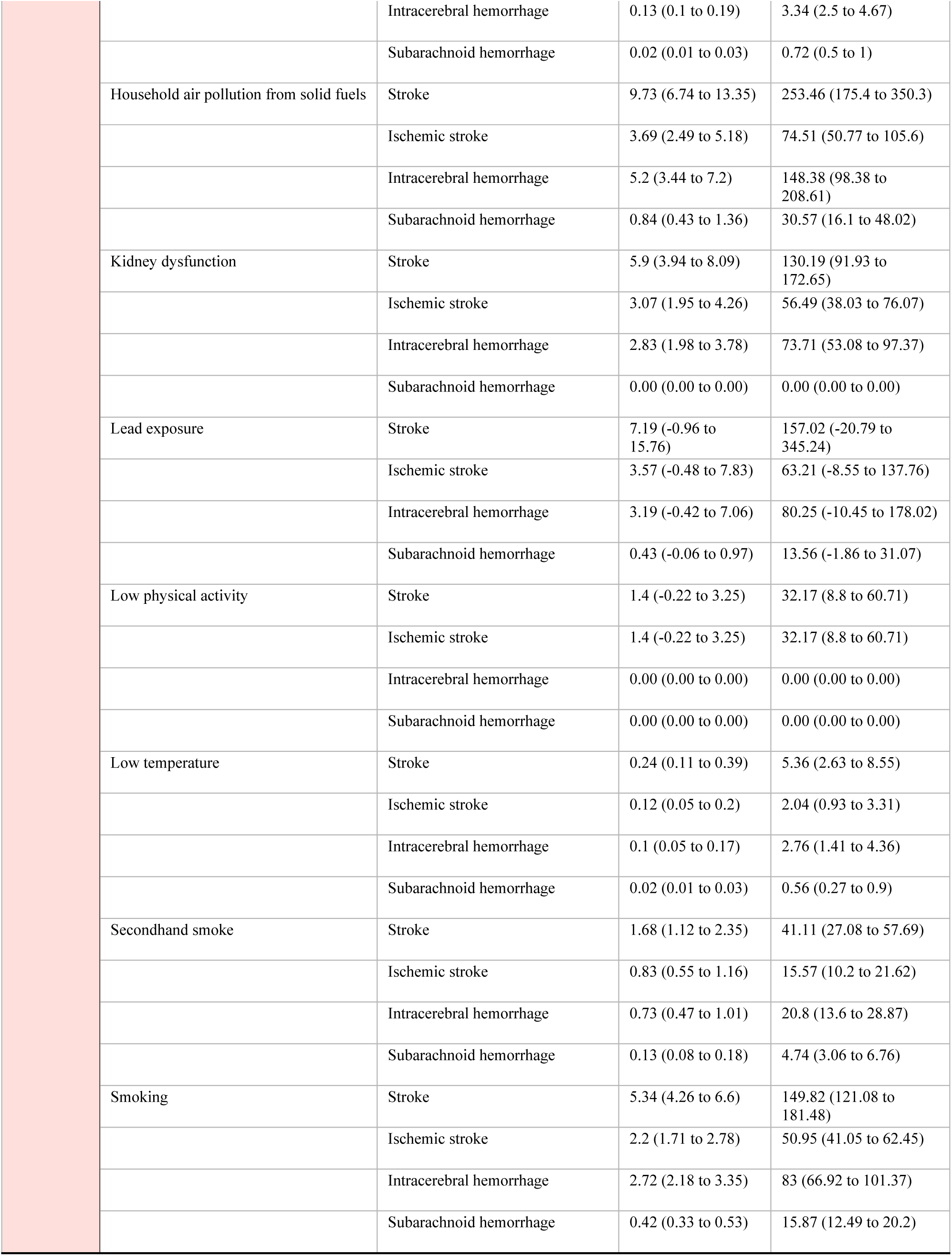
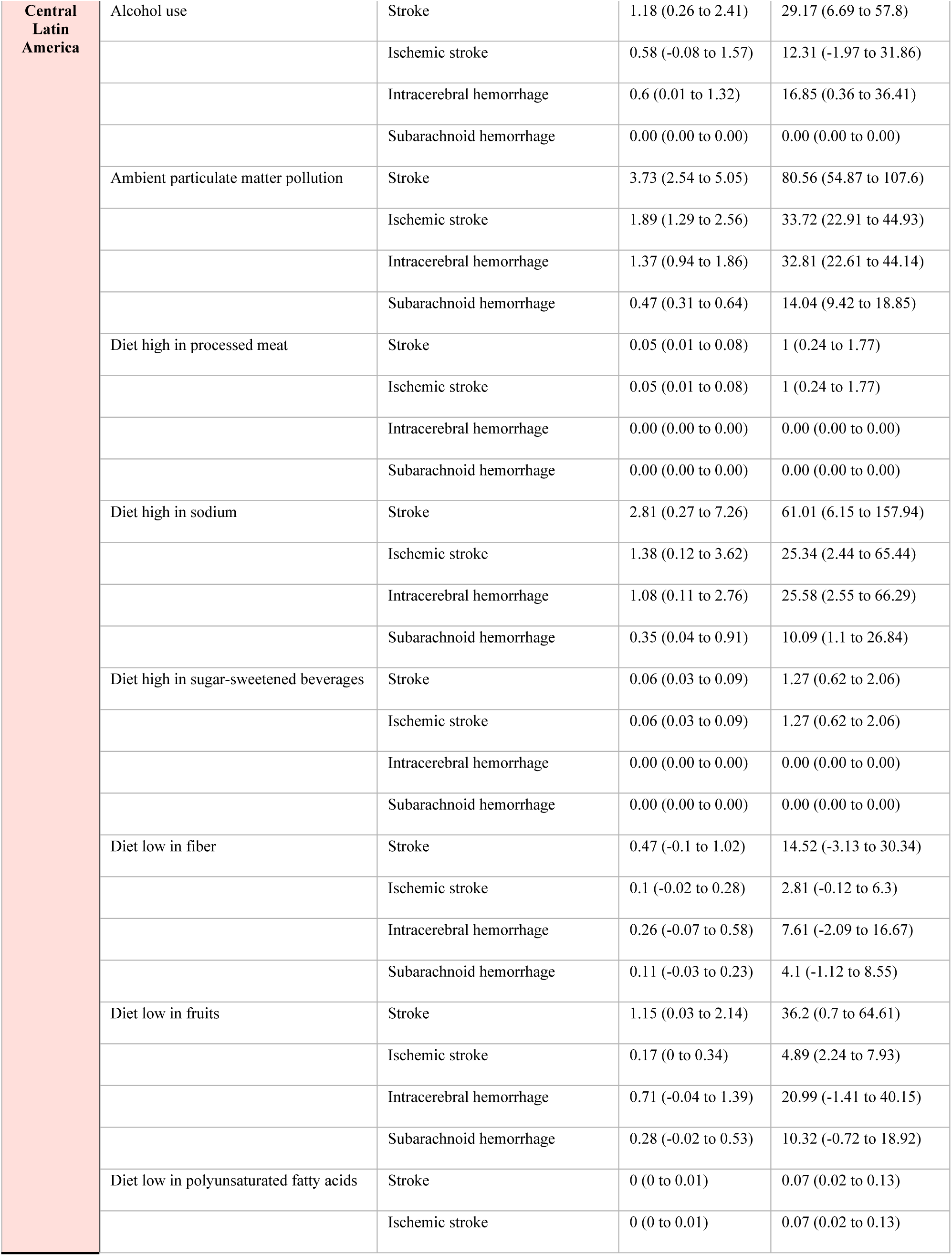
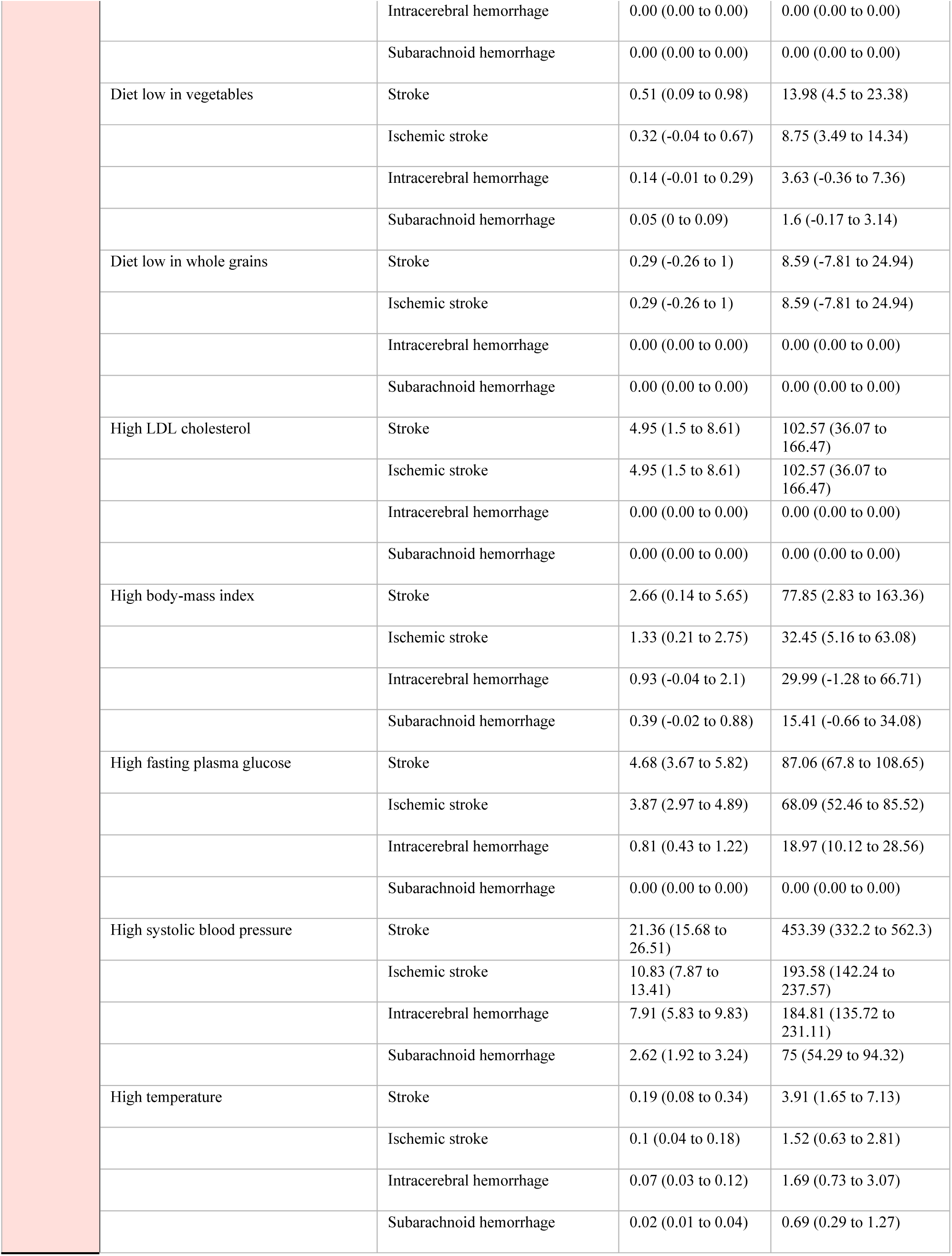
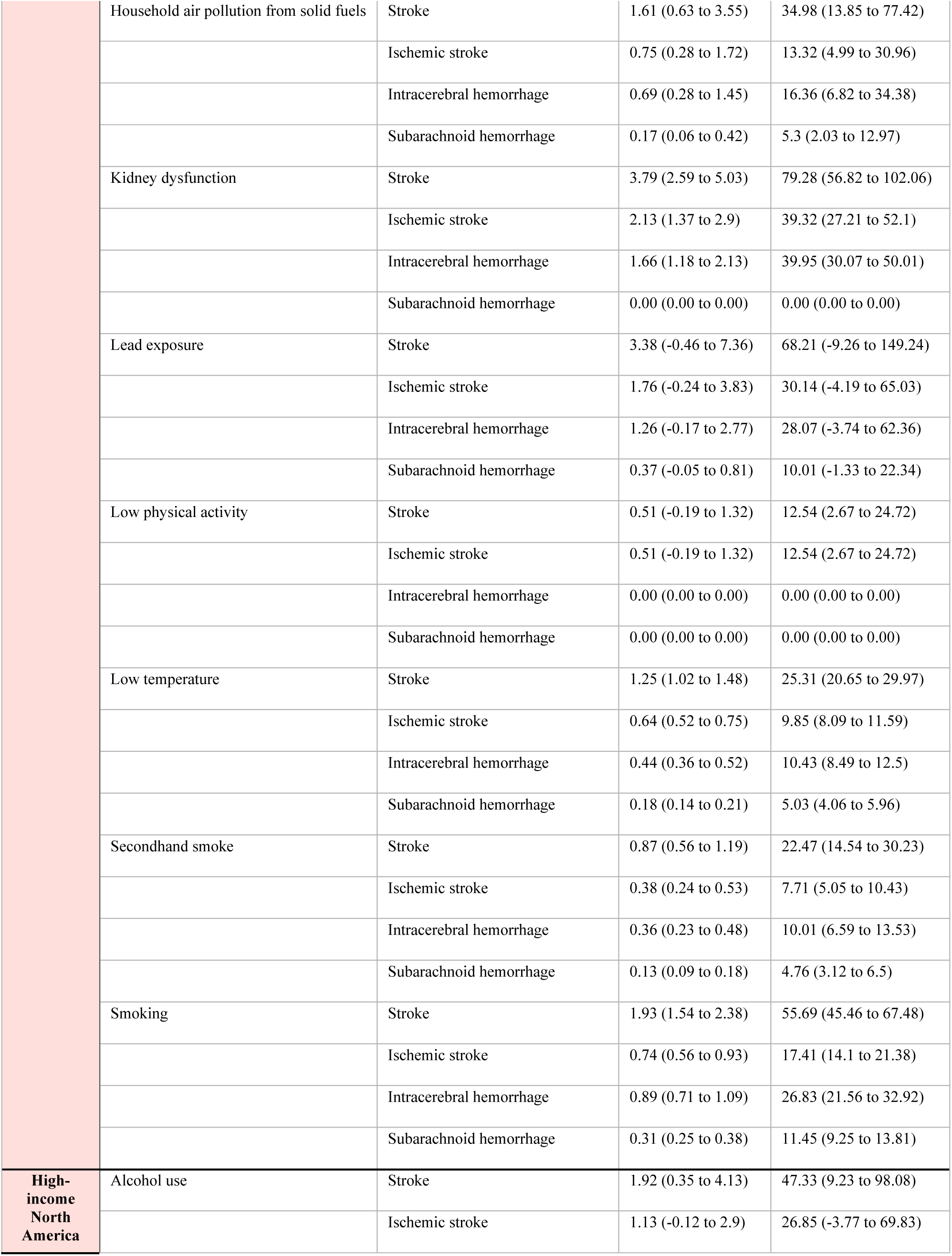
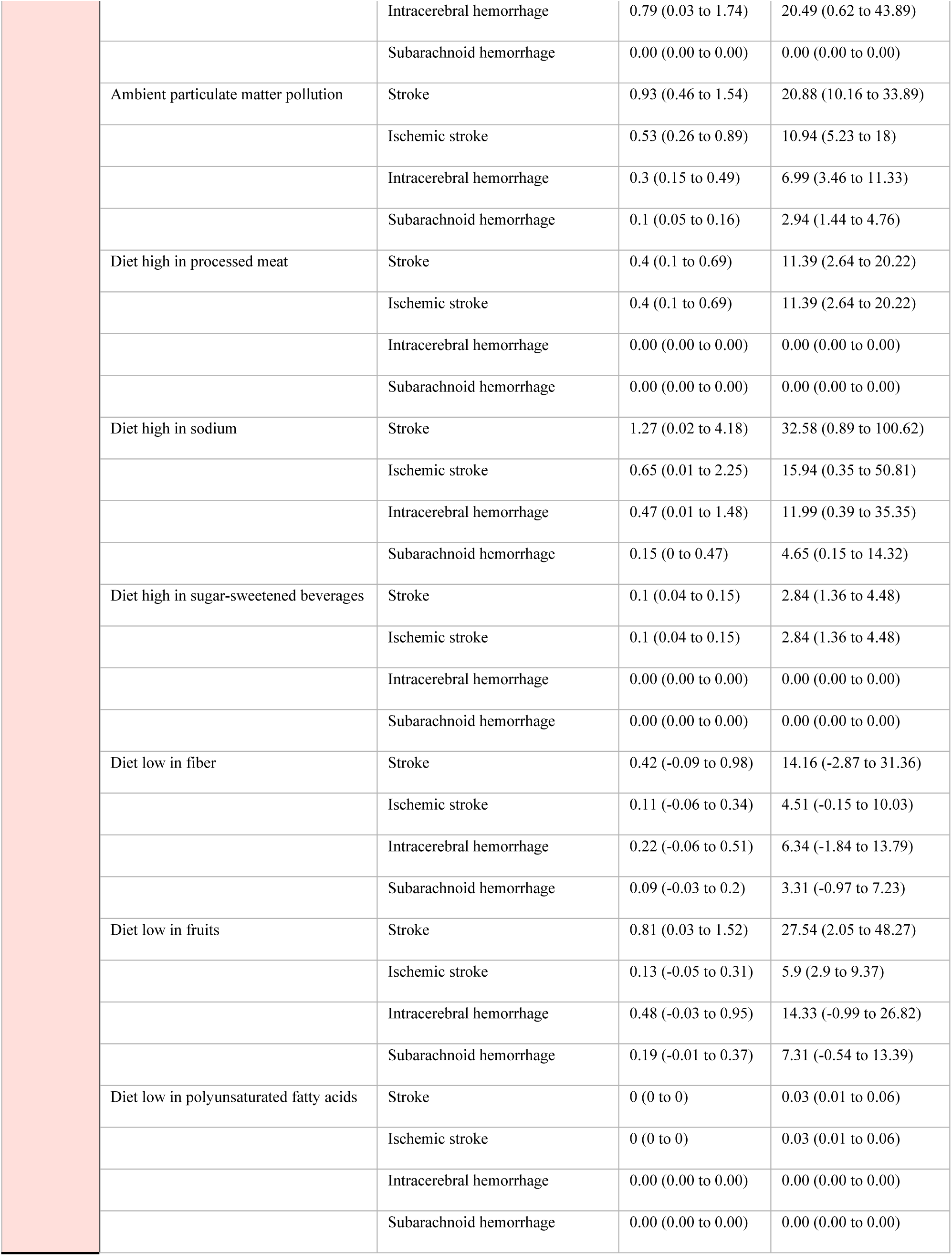
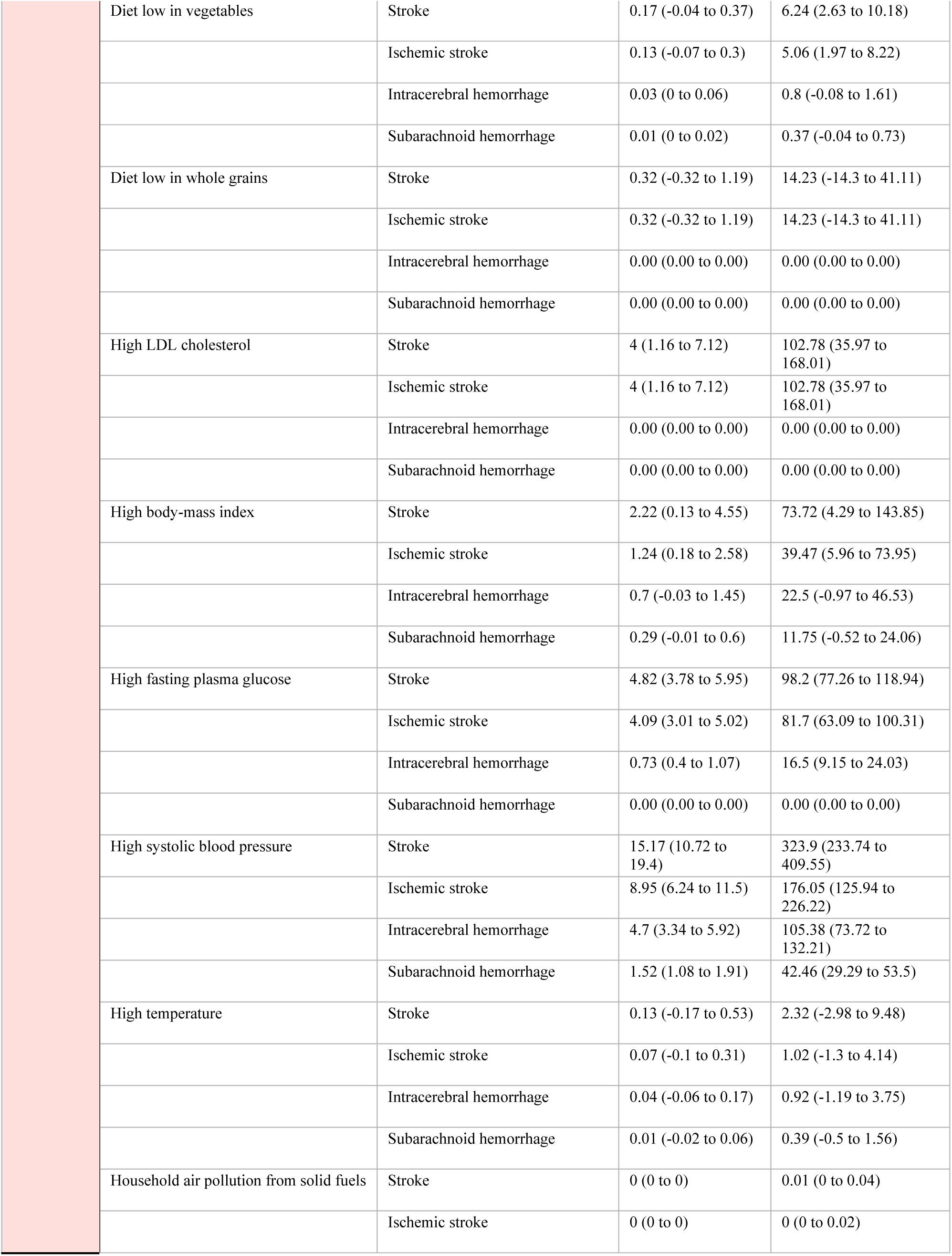
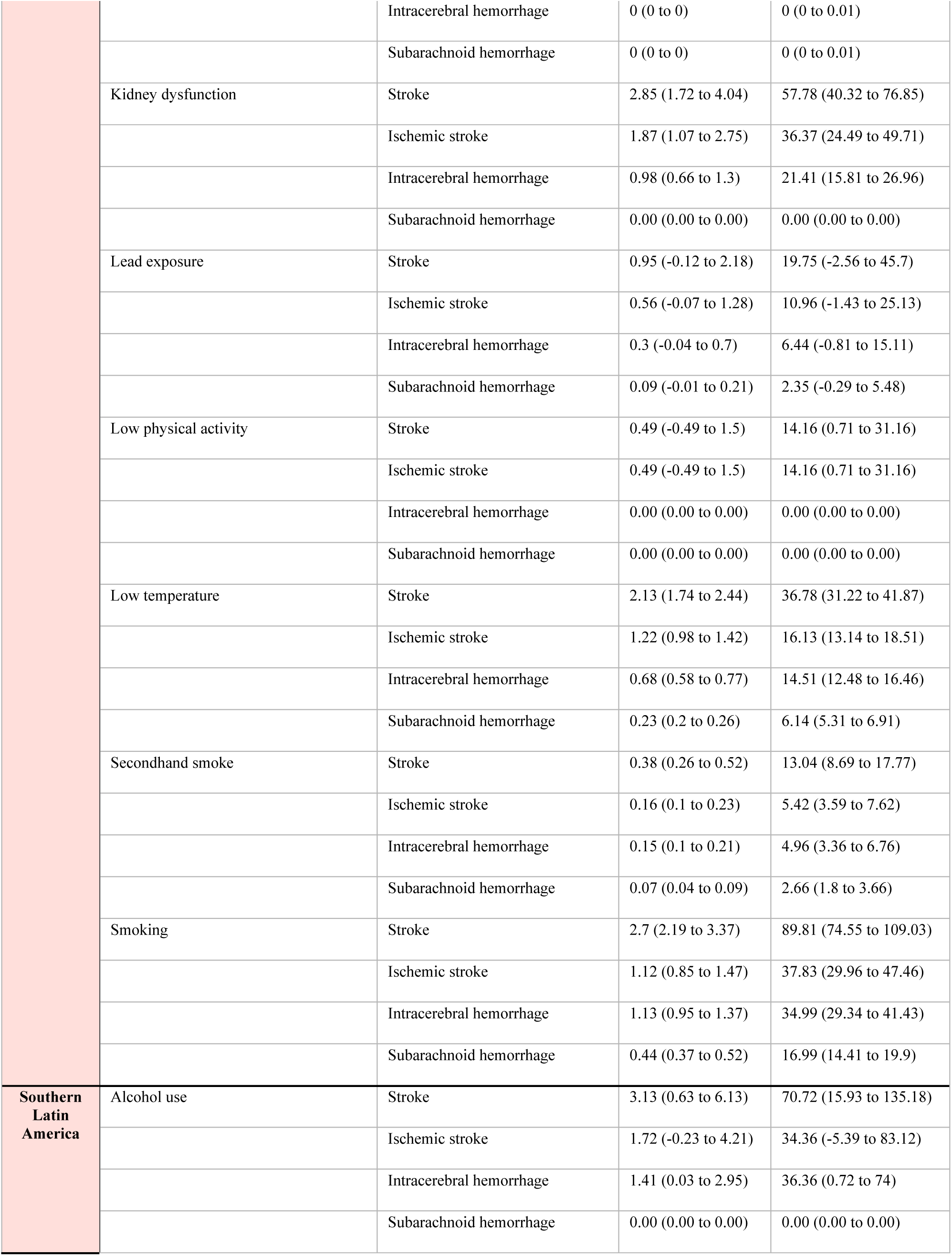
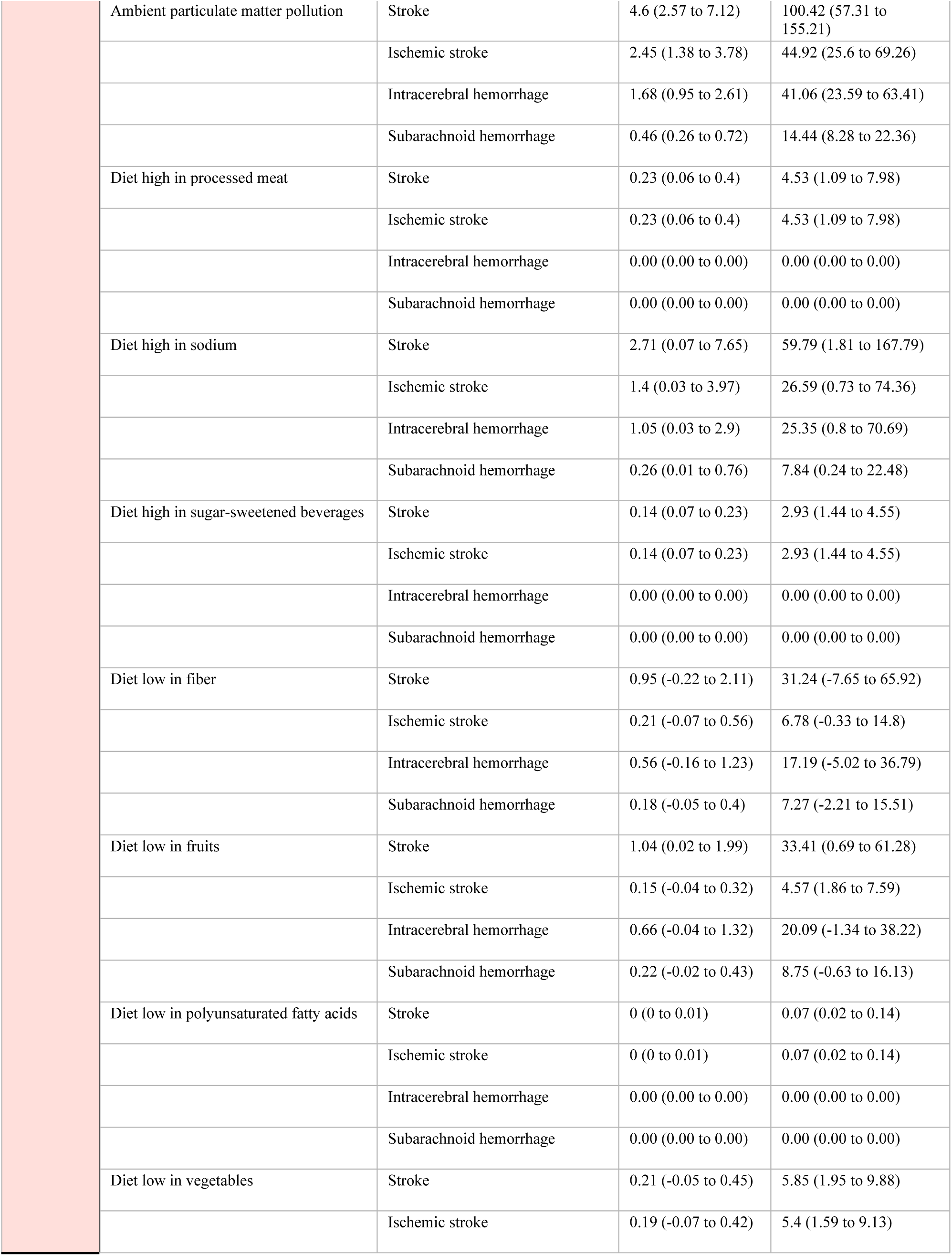
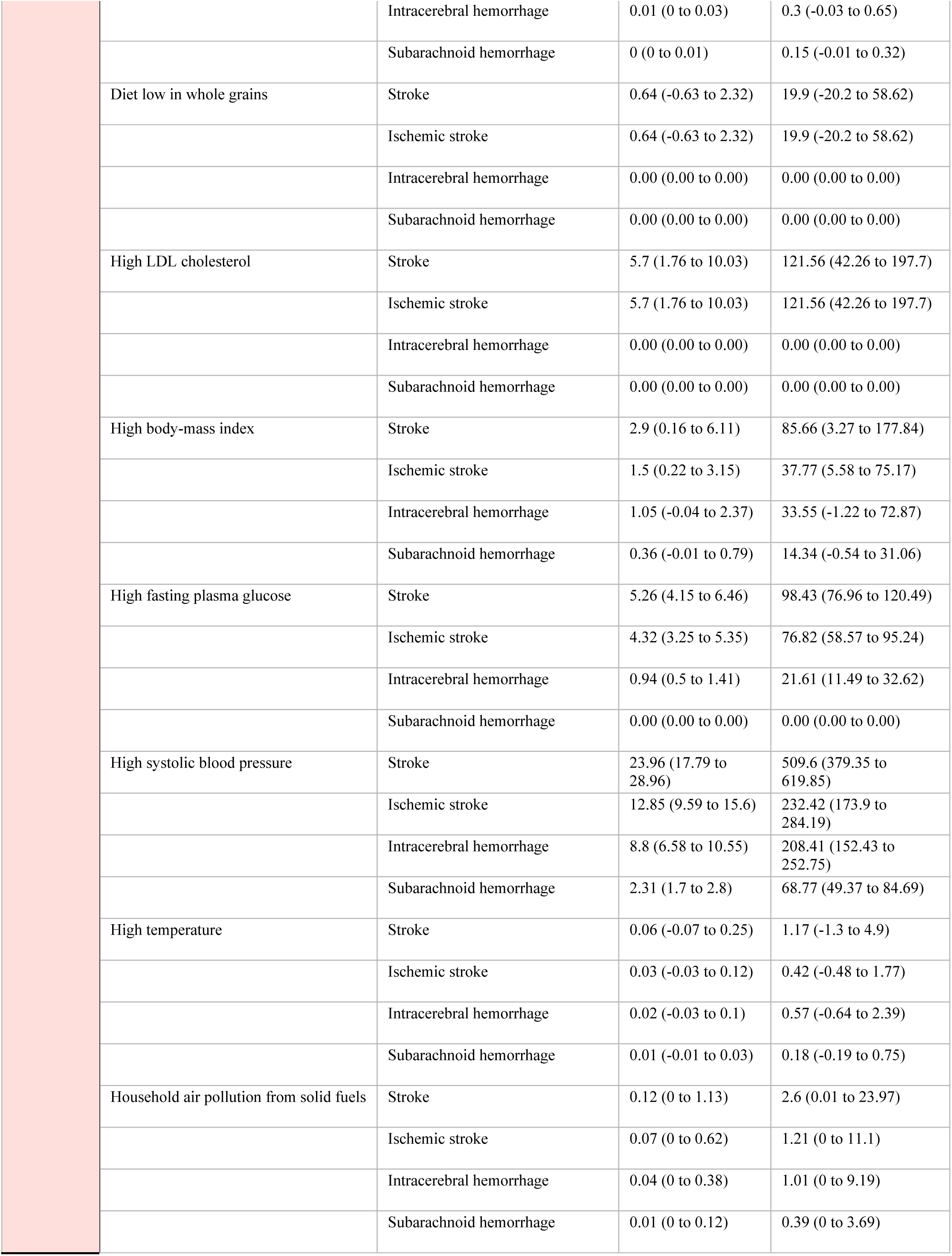
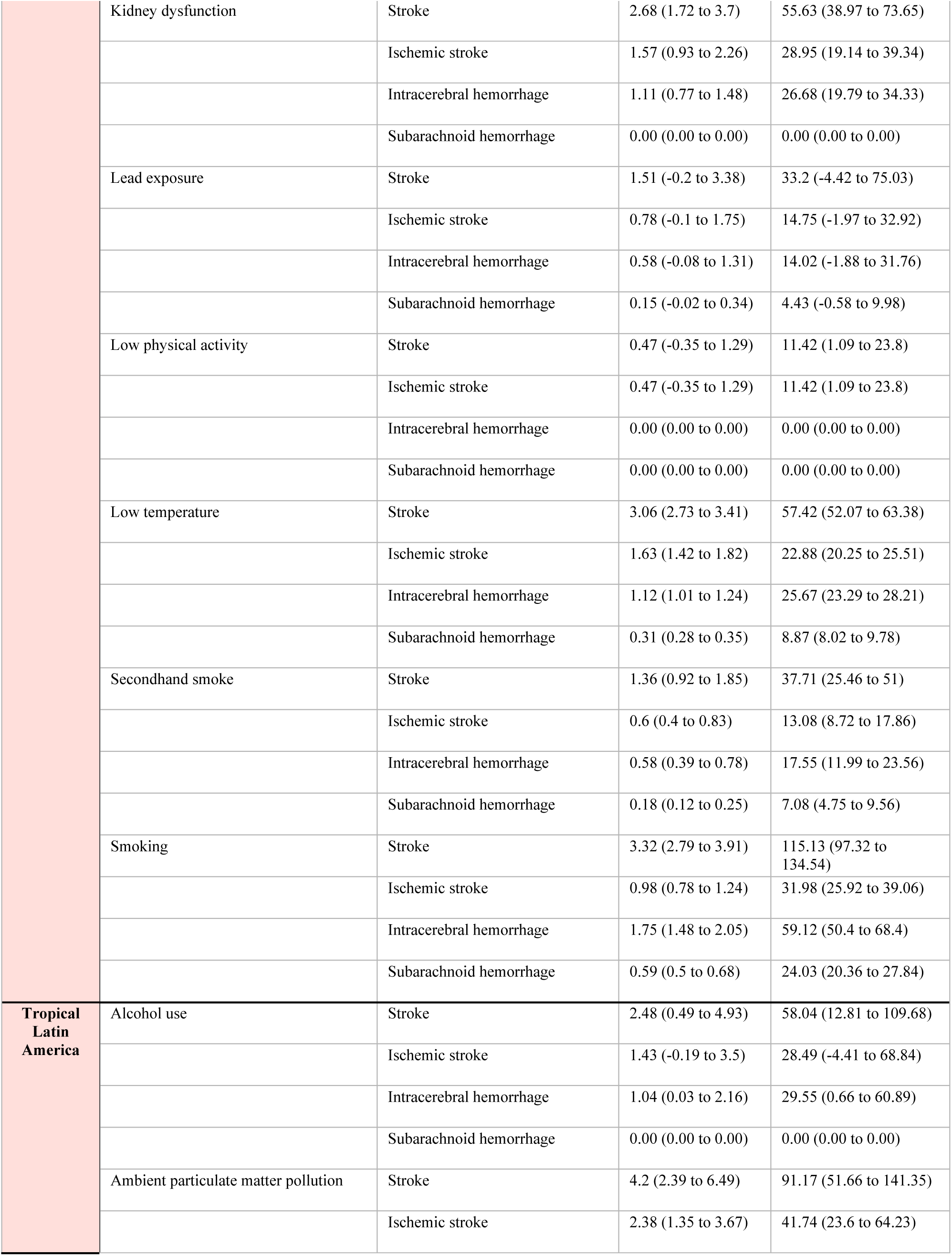
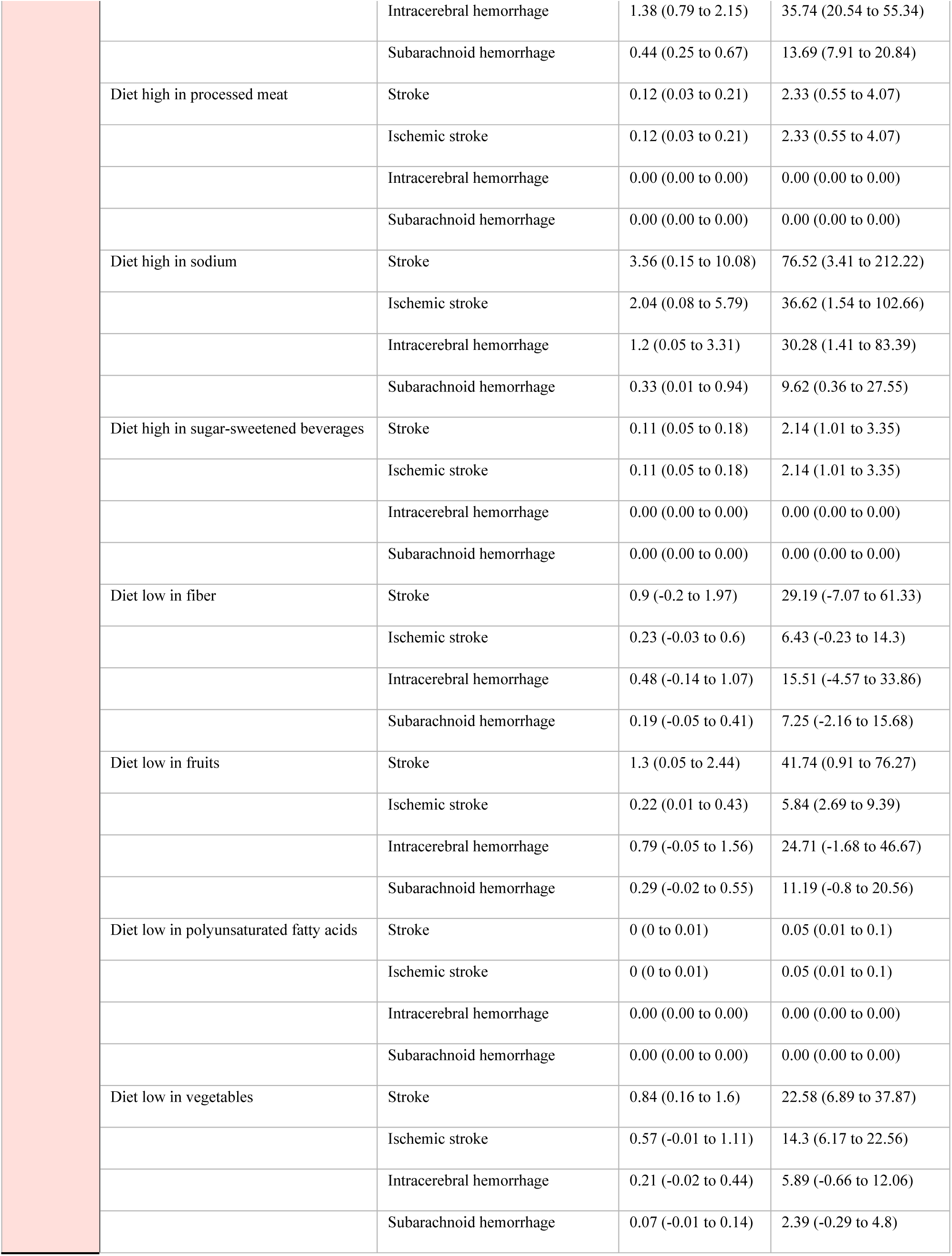
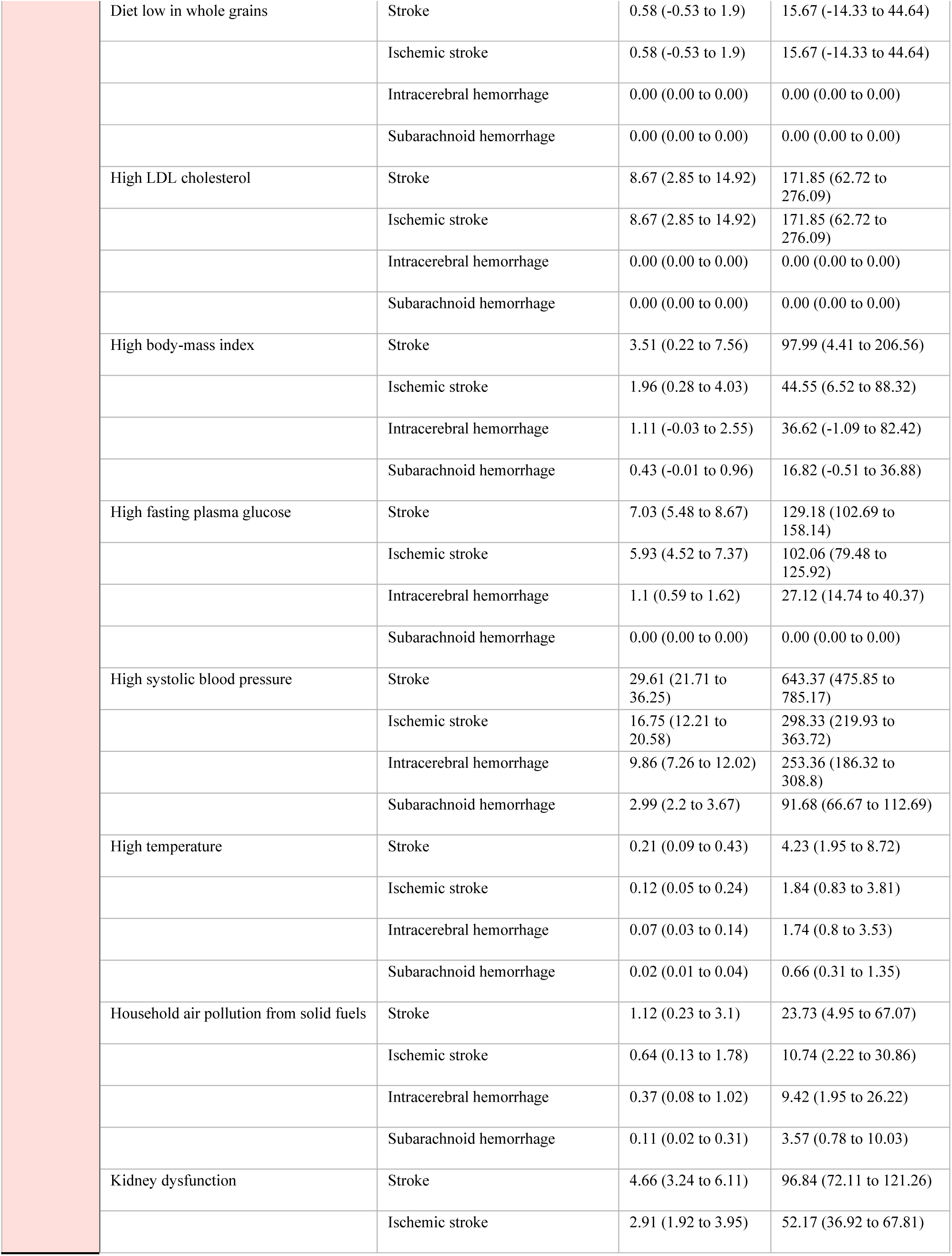
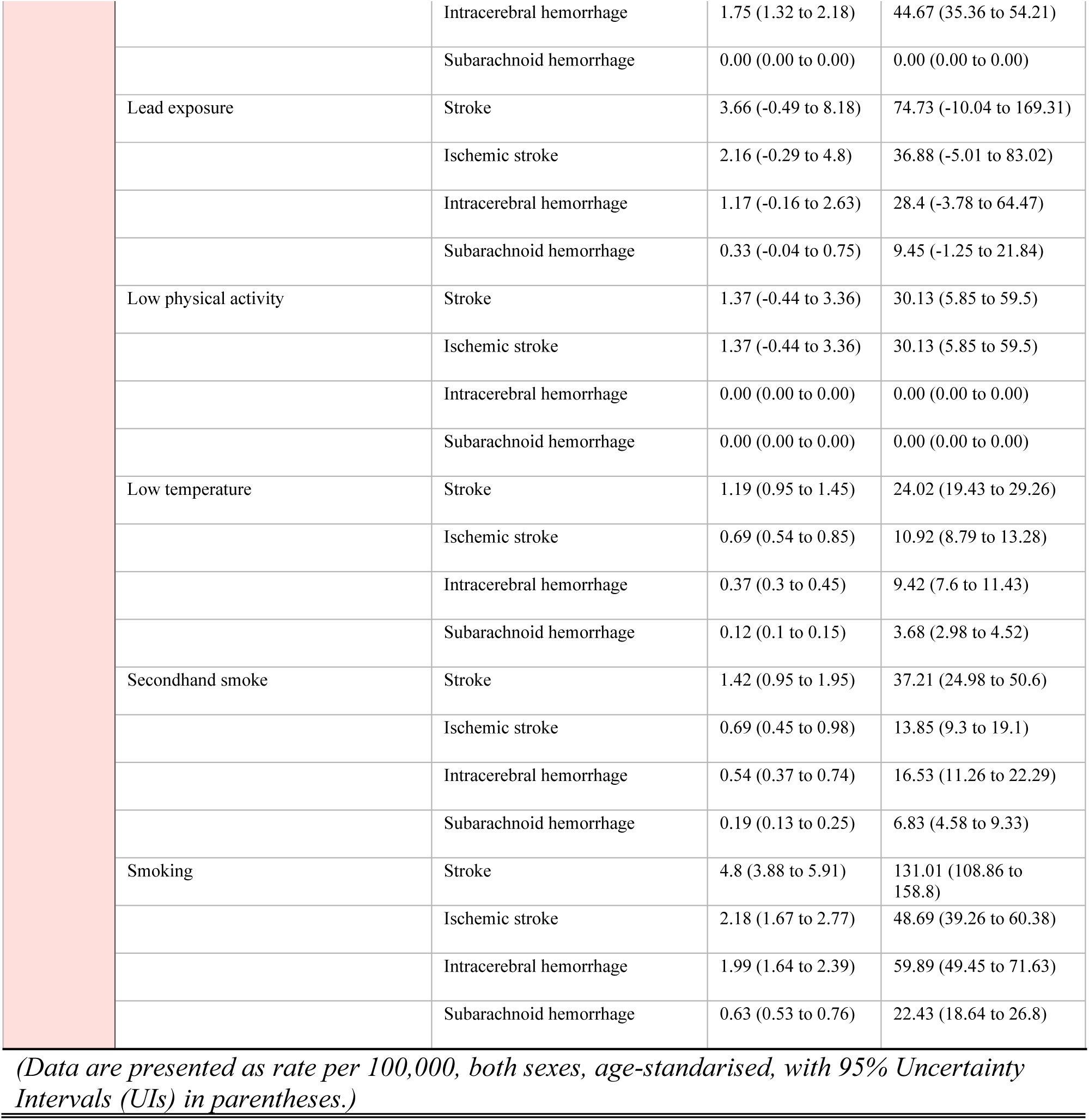
Age-standardized mortality and Disability-Adjusted Life Year (DALY) rates per 100,000 population due to stroke and stroke subtypes attributable to risk factors in the Americas and its six regions, 2021. This table presents the stroke-related mortality and Disability-Adjusted Life Year (DALY) rates per 100,000 population for key risk factors in the Americas in 2021. The data are stratified by stroke subtypes, including ischemic stroke, intracerebral hemorrhage, and subarachnoid hemorrhage. Mortality and DALY rates are reported with 95% Uncertainty Intervals (UIs) in parentheses. DALY rates represent the sum of years of life lost due to premature mortality and years lived with disability. UI = Uncertainty Interval.

#### 1.1. Leading Risk Factors Across the Americas

In 2021, high systolic blood pressure was the leading risk factor attributing stroke in both sexes region-wide, accounting for 20.87 deaths per 100,000 (95% UI 15.17–25.80) and 453.30 DALYs per 100,000 (95% UI 333.16–556.41), contributing to more than half of all stroke deaths (54.8% [95% UI 40.8–65.8]) and DALYs (52.2% [95% UI 38.4–63.1]) (Supplementary Figure 56). High fasting plasma glucose (5.40 [95% UI 4.32–6.66] deaths and 104.11 [95% UI 83.13–127.58] DALYs per 100,000) and high LDL cholesterol (5.39 [95% UI 1.69–9.45] deaths and 119.50 [95% UI 42.98–192.89] DALYs per 100,000) were also major contributors, accounting for 14.2% (95% UI 11.5–17.0) and 12.0% (95% UI 9.6–14.3) of the total burden from high fasting plasma glucose and 14.1% (95% UI 4.4–24.5) and 13.7% (95% UI 4.8–22.2) from high LDL cholesterol, respectively. Among behavioral risk factors, smoking contributed to 3.09 deaths per 100,000 (95% UI 2.55–3.79) and 93.75 DALYs per 100,000 (95% UI 79.32–112.02), accounting for 8.1% (95% UI 6.7–9.7) of stroke-related deaths and 10.8% (95% UI 9.1–12.5) of stroke-related DALYs. Alcohol use accounted for 2.05 deaths per 100,000 (95% UI 0.41–4.27) and 49.12 DALYs per 100,000 (95% UI 10.70–96.96), making up 5.4% (95% UI 1.2–11.2) of total stroke deaths and 5.7% (95% UI 1.2–11.2) of stroke-related DALYs.

Environmental risks, including ambient particulate matter pollution (2.79 [95% UI 1.94–3.87] deaths and 63.06 [95% UI 43.46–86.20] DALYs per 100,000), contributed to the overall stroke burden but to a lesser extent compared to metabolic and behavioral factors. Diet high in red meat or low in fiber had negligible or even negative associations (Table 1, Supplementary Table 1, Supplementary Table 2).

#### 1.2. Variations by Stroke Subtypes

Ischemic stroke accounted for the highest proportion of stroke burden attributable to risk factors, largely driven by high systolic blood pressure (11.44 deaths [95% UI 8.25–14.14]; 212.20 DALYs [95% UI 156.32–261.18]), high LDL cholesterol (5.39 deaths [95% UI 1.69–9.45]; 119.5 DALYs [95% 42.98–192.89]), and high fasting plasma glucose (4.53 deaths [95% UI 3.39–5.58]; 83.48 DALYs [95% UI 65.14–101.97]). Intracerebral hemorrhage was predominantly associated with high systolic blood pressure (7.24 deaths [95% UI 5.31–8.85]; 175.94 DALYs [95% UI 128.28–216.18]) and smoking (1.36 deaths [95% UI 1.15–1.62]; 41.81 DALYs [95% UI 35.61–48.68]), with additional contributions from alcohol use (0.90 deaths [95% UI 0.03–1.94]; 24.31 DALYs [95% UI 0.58–50.8]) and high sodium intake (0.81 deaths [95% UI 0.04–2.33]; 20.43 DALYs [95% UI 1.11–57.89]). Although subarachnoid hemorrhage was less frequent, it was similarly influenced by high systolic blood pressure (2.20 deaths [95% UI 1.61–2.71]; 65.15 DALYs [95% UI 46.98–80.67]) and smoking (0.46 deaths [95% UI 0.39– 0.54]; 17.19 DALYs [95% UI 14.58–20.26]), with ambient particulate matter pollution contributed to a smaller extent (0.32 deaths [95% UI 0.22–0.44]; 10.15 DALYs [95% UI 7.08– 13.88]). (Supplementary Figure 56, Supplementary Figure 63) Across stroke subtypes, high systolic blood pressure remained consistently the principal modifiable risk factor. All estimates are age-standardized rates per 100,000 population in both sexes (Table 1, Supplementary Table 3) (Supplementary Figure 1–6, Supplementary Figure 56, Supplementary Figure 63).

#### 1.3. Subregional and Country-Level Patterns

Across subregions, high systolic blood pressure remained the leading risk factor, accounting for 29.61 deaths (95% UI 21.71–36.25) and 643.37 DALYs (95% UI 475.85–785.17) per 100,000 age-standardized individuals in Tropical Latin America; and 39.87 deaths (95% UI 28.96–49.69) and 903.62 DALYs (95% UI 649.12–1125.52) per 100,000 in the Caribbean, contributing to over half of stroke-related deaths and DALYs in some regions. Other metabolic risks (high fasting plasma glucose, high LDL cholesterol) were consistently high in Tropical Latin America, and the Caribbean. Environmental risks, specifically ambient particulate matter pollution, were prominent in the Caribbean (7.92 deaths [95% UI 4.03–13.16]; 168.62 DALYs [95% UI 85.29–282.18] per 100,000) and Andean Latin America (6.49 deaths [95% UI 4.07–9.77]; 145.48 DALYs [95% UI 91.95–220.08] per 100,000). Dietary factors (high sodium, low fruit/vegetable intake) contributed moderately across all subregions, whilst behavioral risks (smoking, alcohol use) varied but remained important, especially in the Caribbean, and Tropical Latin America (Table 1, Supplementary Table 1, Supplementary Table 2).

At the country level, high systolic blood pressure, high fasting plasma glucose, high body-mass index, high LDL cholesterol, and ambient particulate matter pollution were consistently the top contributors to stroke-related deaths and DALYs. In some countries (e.g., Guyana, Haiti, Honduras), deaths from high systolic blood pressure exceeded 66 per 100,000, with DALYs surpassing 1,300 per 100,000 (Supplementary Table 3). Household air pollution remained significant in lower-income settings (e.g., Haiti, Nicaragua), while alcohol use posed a particular burden in Guyana and Dominica. Across stroke subtypes, high systolic blood pressure consistently ranked among the leading modifiable risk factors, accounting for more than 9.5 deaths per 100,000 individuals from ischemic stroke in Costa Rica, Mexico, and Nicaragua (Supplementary Table 3) (Figure 1, Figure 2).

**Figure 1:**
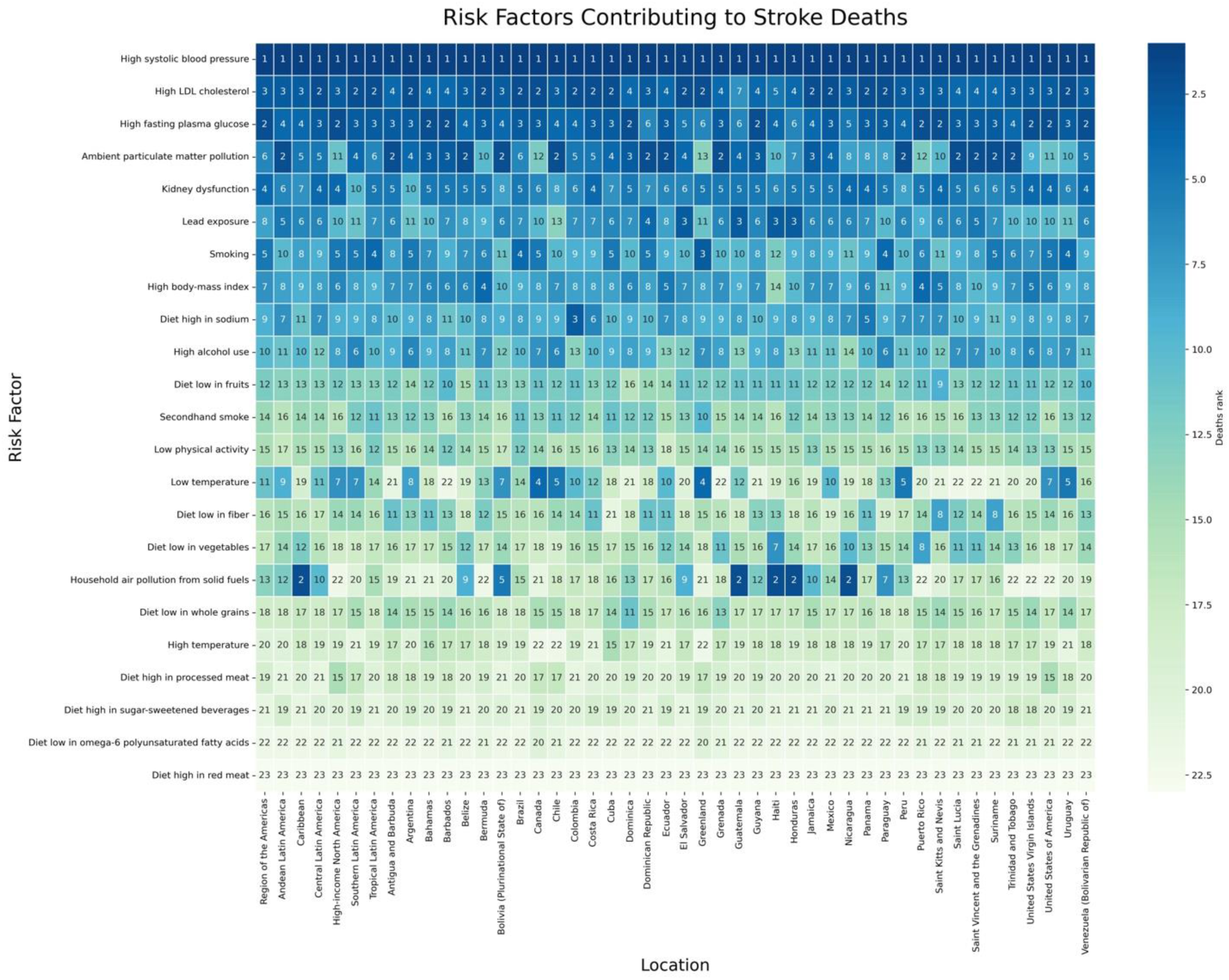
Ranking of Risk Factors Contributing to Stroke Deaths in the Americas, 2021. **Legend:** This heatmap presents the ranking of risk factors contributing to stroke deaths across regions and countries in the Americas in 2021. Each risk factor is ranked from 1 (highest contributor) to 23 (lowest contributor) for each location, with darker shades indicating higher ranks.

**Figure 2:**
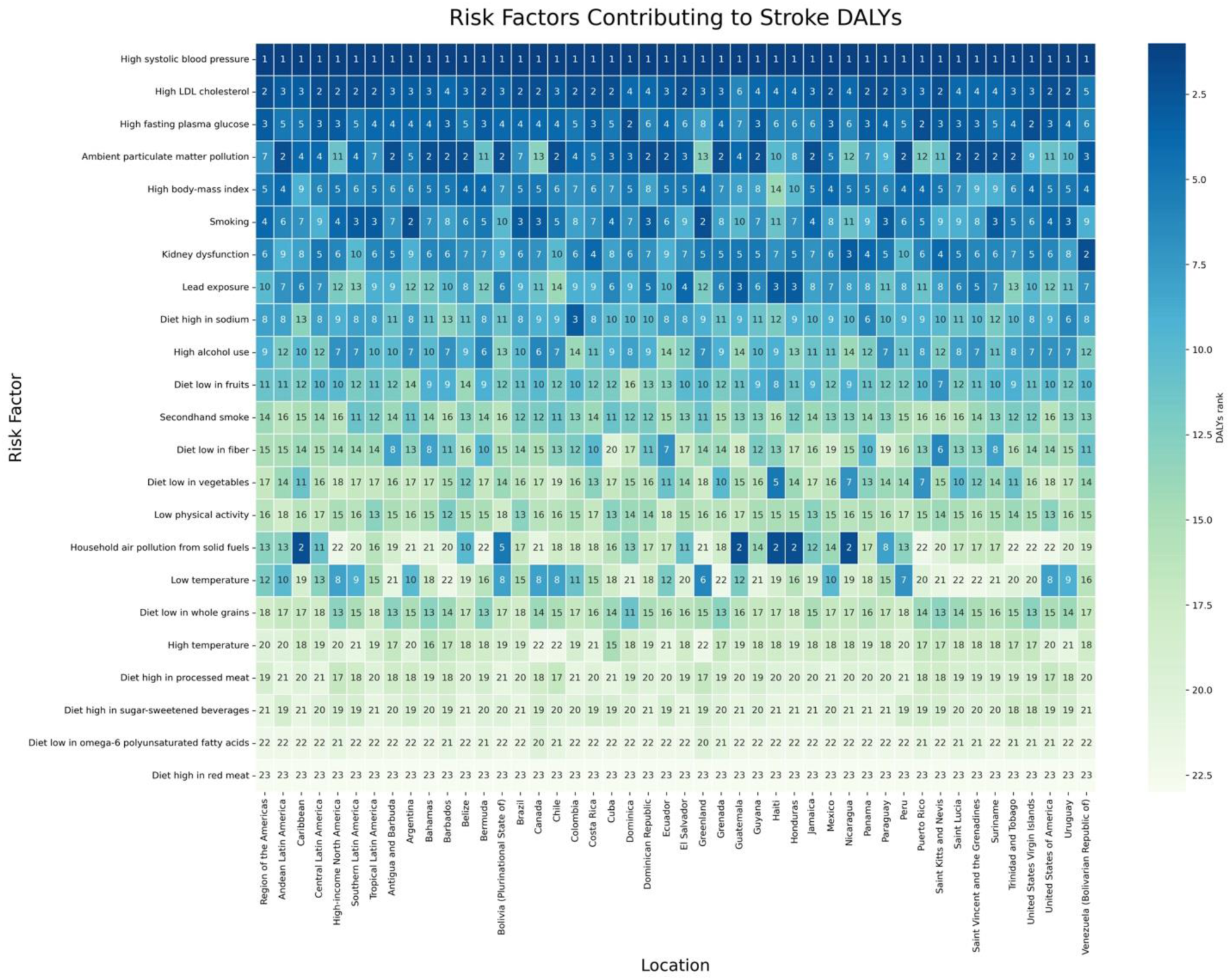
Ranking of Risk Factors Contributing to Stroke DALYs in the Americas, 2021. **Legend:** This heatmap illustrates the ranking of risk factors contributing to Disability-Adjusted Life Years (DALYs) from stroke across regions and countries in the Americas in 2021. Each risk factor is ranked from 1 (highest contributor) to 23 (lowest contributor) for each

#### 1.4. Variations by SDI

The disparity was particularly pronounced for high blood pressure, which led to 10.42 (95% UI 7.13–13.29) deaths per 100,000 and 335.94 (95% UI 241.27–424.52) DALYs per 100,000 in high-SDI regions but soared to 109.33 (95% UI 71.33–149.17) deaths per 100,000 and 2395.53 DALYs per 100,000 (95% UI 1570.32–3249.86) in low-SDI regions. High fasting plasma glucose and high LDL cholesterol similarly carried a higher burden in low- and middle-SDI areas relative to high-SDI. Alcohol use accounted for 1.99 deaths (95% UI 0.37–4.26) per 100,000 and 48.87 DALYs (95% UI 9.61–101.19) per 100,000 in high-SDI regions, compared to 8.00 deaths (95% UI 1.79–17.32) per 100,000 and 184.68 DALYs (95% UI 40.99–398.60) per 100,000 in low-SDI regions. Similarly, smoking contributed to 1.69 deaths (95% UI 1.34–2.15) per 100,000 and 93.42 DALYs (95% UI 77.59–112.83) in high-SDI areas, rising to 5.45 deaths (95% UI 3.66–7.88) per 100,000 and 2395.53 DALYs (1570.32–3249.86) in low-SDI areas.

Household air pollution had a negligible impact in high-SDI areas but significantly impacted stroke burden in lower SDI regions, whereas alcohol use was more burdensome in low-SDI regions. Dietary risk factors (low fruits/vegetables, low whole grains) influenced stroke burden across SDI levels, and its impact was slightly more pronounced in lower SDI strata (Table 2).

**Table 2:** Age-standardized mortality and DALY rates per 100,000 population due to stroke attributable to risk factors in both sexes combined across SDI quintile groups of countries in the Americas, 2021. This table presents stroke-related mortality and Disability-Adjusted Life Year (DALY) rates per 100,000 population across **Socio-Demographic Index (SDI) quintiles** in the Americas in 2021. The data are stratified by SDI levels (High, High-Middle, Middle, Low-Middle, and Low) and key risk factors. Mortality and DALY rates are reported with 95% Uncertainty Intervals (UIs) in parentheses. DALY rates represent the sum of years of life lost due to premature mortality and years lived with disability. UI = Uncertainty Interval; SDI = Socio-Demographic Index.

| SDI Quintile | Risk Factor | Death Rate per 100,000, (95% UI) | DALYs Rate per 100,000, (95% UI) |
| --- | --- | --- | --- |
| High SDI | Alcohol use | 1.99 (0.37 to 4.26) | 48.87 (9.61 to 101.19) |
|  | Ambient particulate matter pollution | 0.47 (0.17 to 0.87) | 22.12 (10.95 to 35.67) |
|  | Diet high in processed meat | 0.09 (0.02 to 0.16) | 2.48 (0.58 to 4.38) |
|  | Diet high in red meat | -0.67 (-2.66 to 0.96) | -22.29 (-93.70 to 31.53) |
|  | Diet high in sodium | 0.94 (0.01 to 3.01) | 24.22 (0.60 to 74.30) |
|  | Diet high in sugar-sweetened beverages | 0.03 (0.01 to 0.05) | 0.87 (0.42 to 1.44) |
|  | Diet low in fiber | 0.45 (-0.09 to 1.04) | 6.92 (-1.46 to 14.63) |
|  | Diet low in fruits | 0.49 (0.01 to 0.94) | 28.73 (2.11 to 50.41) |
|  | Diet low in polyunsaturated fatty acids | 0.00 (0.00 to 0.00) | 0.06 (0.02 to 0.12) |
|  | Diet low in vegetables | 0.07 (-0.04 to 0.17) | 3.03 (1.18 to 5.19) |
|  | Diet low in whole grains | 0.33 (-0.34 to 1.25) | 9.72 (-9.39 to 28.14) |
|  | High LDL cholesterol | 3.12 (0.87 to 5.51) | 83.84 (29.32 to 137.45) |
|  | High body-mass index | 2.35 (0.14 to 4.79) | 77.60 (4.45 to 151.21) |
|  | High fasting plasma glucose | 2.33 (1.74 to 2.91) | 104.46 (82.29 to 126.35) |
|  | High systolic blood pressure | 10.42 (7.13 to 13.29) | 335.94 (241.27 to 424.52) |
|  | High temperature | -0.02 (-0.07 to 0.02) | -0.34 (-1.15 to 0.26) |
|  | Household air pollution from solid fuels | 0.00 (0.00 to 0.00) | 0.01 (0.00 to 0.04) |
|  | Kidney dysfunction | 1.60 (0.92 to 2.35) | 32.97 (21.79 to 44.99) |
|  | Lead exposure | 0.60 (-0.08 to 1.33) | 20.58 (-2.66 to 47.82) |
|  | Low physical activity | 0.24 (-0.32 to 0.90) | 6.87 (-0.97 to 16.84) |
|  | Low temperature | 2.16 (1.75 to 2.48) | 30.44 (25.82 to 34.56) |
|  | Secondhand smoke | 0.32 (0.21 to 0.43) | 13.11 (8.73 to 17.94) |
|  | Smoking | 1.69 (1.34 to 2.15) | 93.42 (77.59 to 112.83) |
| High-middle SDI | Alcohol use | 3.20 (0.57 to 6.37) | 71.20 (16.38 to 132.00) |
|  | Ambient particulate matter pollution | 3.99 (2.03 to 6.54) | 90.27 (46.35 to 148.28) |
|  | Diet high in processed meat | 0.27 (0.07 to 0.49) | 3.51 (0.85 to 6.25) |
|  | Diet high in red meat | -1.96 (-8.56 to 3.01) | -71.17 (-323.39 to 101.14) |
|  | Diet high in sodium | 3.70 (0.12 to 10.30) | 59.80 (1.38 to 170.39) |
|  | Diet high in sugar-sweetened beverages | 0.16 (0.08 to 0.26) | 3.30 (1.65 to 5.28) |
|  | Diet low in fiber | 0.64 (-0.13 to 1.49) | 36.53 (-9.22 to 75.95) |
|  | Diet low in fruits | 1.44 (0.07 to 2.69) | 31.87 (0.26 to 60.38) |
|  | Diet low in polyunsaturated fatty acids | 0.00 (0.00 to 0.01) | 0.07 (0.02 to 0.14) |
|  | Diet low in vegetables | 0.20 (-0.04 to 0.42) | 5.75 (1.97 to 9.68) |
|  | Diet low in whole grains | 0.60 (-0.62 to 2.25) | 19.42 (-19.79 to 57.31) |
|  | High LDL cholesterol | 5.25 (1.62 to 9.23) | 183.72 (63.63 to 297.86) |
|  | High body-mass index | 2.78 (0.18 to 5.81) | 87.49 (2.94 to 183.63) |
|  | High fasting plasma glucose | 5.12 (3.98 to 6.35) | 99.29 (76.65 to 121.61) |
|  | High systolic blood pressure | 23.08 (16.81 to 28.10) | 515.03 (379.06 to 630.10) |
|  | High temperature | 0.09 (-0.10 to 0.37) | 0.63 (-1.01 to 3.63) |
|  | Household air pollution from solid fuels | 0.07 (0.00 to 0.64) | 1.61 (0.01 to 13.84) |
|  | Kidney dysfunction | 2.58 (1.68 to 3.56) | 56.11 (39.63 to 73.98) |
|  | Lead exposure | 0.93 (-0.12 to 2.12) | 36.55 (-4.85 to 82.35) |
|  | Low physical activity | 0.43 (-0.31 to 1.21) | 10.39 (0.70 to 22.02) |
|  | Low temperature | 3.54 (3.10 to 4.11) | 54.07 (48.25 to 59.09) |
|  | Secondhand smoke | 1.27 (0.85 to 1.73) | 38.27 (25.92 to 51.17) |
|  | Smoking | 3.74 (3.13 to 4.46) | 127.34 (107.21 to 149.74) |
| **Middle SDI** | Alcohol use | 2.46 (0.45 to 5.21) | 108.18 (25.78 to 213.76) |
|  | Ambient particulate matter pollution | 8.01 (3.73 to 13.72) | 226.94 (80.06 to 421.56) |
|  | Diet high in processed meat | 0.09 (0.02 to 0.16) | 1.39 (0.33 to 2.49) |
|  | Diet high in red meat | -1.26 (-4.88 to 1.72) | -20.82 (-79.19 to 30.40) |
|  | Diet high in sodium | 2.41 (0.01 to 8.32) | 70.50 (0.24 to 241.80) |
|  | Diet high in sugar-sweetened beverages | 0.12 (0.06 to 0.19) | 1.61 (0.75 to 2.70) |
|  | Diet low in fiber | 0.10 (-0.02 to 0.22) | 61.60 (-12.71 to 132.28) |
|  | Diet low in fruits | 1.17 (0.07 to 2.25) | 57.52 (1.63 to 108.90) |
|  | Diet low in polyunsaturated fatty acids | 0.01 (0.00 to 0.01) | 0.12 (0.03 to 0.23) |
|  | Diet low in vegetables | 0.23 (-0.03 to 0.49) | 37.14 (7.00 to 68.11) |
|  | Diet low in whole grains | 0.58 (-0.53 to 1.90) | 38.98 (-41.99 to 120.46) |
|  | High LDL cholesterol | 8.63 (2.61 to 15.06) | 229.68 (74.51 to 394.39) |
|  | High body-mass index | 3.49 (0.21 to 7.50) | 108.92 (5.28 to 227.89) |
|  | High fasting plasma glucose | 8.52 (6.62 to 10.60) | 216.02 (167.51 to 264.70) |
|  | High systolic blood pressure | 27.26 (18.88 to 34.96) | 389.98 (261.21 to 537.52) |
|  | High temperature | 0.35 (0.12 to 0.65) | 7.50 (4.49 to 11.24) |
|  | Household air pollution from solid fuels | 0.32 (0.05 to 1.19) | 6.27 (0.94 to 23.07) |
|  | Kidney dysfunction | 4.22 (2.82 to 5.76) | 150.68 (106.96 to 196.98) |
|  | Lead exposure | 4.92 (-0.67 to 11.04) | 123.88 (-16.07 to 282.75) |
|  | Low physical activity | 1.17 (-0.41 to 2.91) | 33.25 (1.11 to 70.36) |
|  | Low temperature | 0.00 (-0.03 to 0.03) | 0.16 (-0.74 to 1.06) |
|  | Secondhand smoke | 1.81 (1.16 to 2.52) | 17.51 (10.36 to 25.92) |
|  | Smoking | 5.52 (4.31 to 6.85) | 99.83 (77.92 to 126.33) |
| **Low-middle SDI** | Alcohol use | 2.08 (0.52 to 4.16) | 52.27 (11.71 to 103.46) |
|  | Ambient particulate matter pollution | 9.52 (3.67 to 17.46) | 195.93 (77.06 to 360.70) |
|  | Diet high in processed meat | 0.04 (0.01 to 0.06) | 0.67 (0.16 to 1.17) |
|  | Diet high in red meat | -0.54 (-2.12 to 0.83) | -17.83 (-69.46 to 26.68) |
|  | Diet high in sodium | 2.42 (0.01 to 8.10) | 46.57 (0.20 to 158.96) |
|  | Diet high in sugar-sweetened beverages | 0.03 (0.01 to 0.05) | 0.98 (0.40 to 1.82) |
|  | Diet low in fiber | 0.57 (-0.12 to 1.28) | 16.96 (-3.80 to 36.34) |
|  | Diet low in fruits | 0.64 (0.01 to 1.28) | 20.27 (0.07 to 38.81) |
|  | Diet low in polyunsaturated fatty acids | 0.00 (0.00 to 0.01) | 0.07 (0.02 to 0.14) |
|  | Diet low in vegetables | 1.39 (0.22 to 2.60) | 62.40 (9.53 to 115.87) |
|  | Diet low in whole grains | 0.62 (-0.59 to 2.20) | 16.63 (-16.02 to 48.00) |
|  | High LDL cholesterol | 6.34 (1.91 to 11.09) | 144.50 (47.20 to 251.60) |
|  | High body-mass index | 4.18 (0.18 to 8.54) | 115.77 (2.96 to 232.04) |
|  | High fasting plasma glucose | 6.11 (4.65 to 7.71) | 105.64 (80.67 to 132.57) |
|  | High systolic blood pressure | 29.56 (20.63 to 37.16) | 625.18 (394.99 to 940.87) |
|  | High temperature | 0.58 (0.38 to 0.93) | 3.90 (-1.94 to 12.24) |
|  | Household air pollution from solid fuels | 2.42 (0.40 to 7.06) | 49.33 (8.33 to 145.23) |
|  | Kidney dysfunction | 4.60 (3.14 to 6.17) | 95.90 (70.47 to 123.80) |
|  | Lead exposure | 3.10 (-0.39 to 7.13) | 59.63 (-7.40 to 139.42) |
|  | Low physical activity | 0.86 (-0.47 to 2.24) | 18.53 (1.33 to 38.24) |
|  | Low temperature | 0.16 (0.06 to 0.26) | 3.08 (1.14 to 5.07) |
|  | Secondhand smoke | 1.10 (0.70 to 1.57) | 26.98 (17.26 to 38.00) |
|  | Smoking | 3.48 (2.66 to 4.51) | 96.13 (76.14 to 121.76) |
| **Low SDI** | Alcohol use | 8.00 (1.79 to 17.32) | 184.68 (40.99 to 398.60) |
|  | Ambient particulate matter pollution | 7.72 (3.14 to 15.32) | 161.92 (65.97 to 319.52) |
|  | Diet high in processed meat | 0.09 (0.02 to 0.17) | 1.64 (0.44 to 3.05) |
|  | Diet high in red meat | -1.24 (-4.94 to 1.85) | -40.41 (-154.08 to 60.55) |
|  | Diet high in sodium | 7.83 (0.03 to 28.80) | 154.97 (0.47 to 581.79) |
|  | Diet high in sugar-sweetened beverages | 0.03 (0.01 to 0.05) | 0.50 (0.23 to 0.92) |
|  | Diet low in fiber | 3.91 (-0.88 to 8.81) | 114.54 (-27.54 to 254.27) |
|  | Diet low in fruits | 7.34 (0.13 to 14.62) | 213.42 (0.70 to 405.05) |
|  | Diet low in polyunsaturated fatty acids | 0.02 (0.00 to 0.03) | 0.28 (0.07 to 0.58) |
|  | Diet low in vegetables | 13.61 (2.12 to 26.24) | 376.46 (53.66 to 706.84) |
|  | Diet low in whole grains | 2.46 (-2.37 to 8.57) | 60.60 (-64.06 to 180.62) |
|  | High LDL cholesterol | 20.34 (5.99 to 39.86) | 383.06 (122.79 to 715.50) |
|  | High body-mass index | 3.61 (0.31 to 7.97) | 110.29 (7.46 to 238.26) |
|  | High fasting plasma glucose | 20.66 (14.41 to 28.27) | 356.91 (246.19 to 493.56) |
|  | High systolic blood pressure | 109.33 (71.33 to 149.17) | 2395.53 (1570.32 to 3249.86) |
|  | High temperature | 0.72 (0.49 to 1.02) | 15.32 (10.33 to 21.91) |
|  | Household air pollution from solid fuels | 81.95 (57.47 to 112.59) | 1717.30 (1196.00 to 2384.66) |
|  | Kidney dysfunction | 16.03 (9.71 to 23.85) | 334.31 (212.82 to 485.01) |
|  | Lead exposure | 25.42 (-3.25 to 55.09) | 516.03 (-66.48 to 1121.69) |
|  | Low physical activity | 3.21 (-0.66 to 8.32) | 67.44 (14.65 to 141.92) |
|  | Low temperature | 0.45 (0.22 to 0.76) | 9.68 (4.64 to 16.09) |
|  | Secondhand smoke | 2.62 (1.48 to 3.96) | 64.42 (35.84 to 97.94) |
|  | Smoking | 5.45 (3.66 to 7.88) | 156.50 (105.45 to 221.11) |

#### 1.5. Sex- and Age-Specific Disparities

Both sexes were most affected by high systolic blood pressure, but males bore a greater in burden (22.29 deaths [95% UI 16.57–27.54] vs 19.41 deaths [95% UI 14.04–24.15] per 100,000 in females; 504.76 DALYs [95% UI 372.54–625.02] vs 404.91 DALYs [95% UI 295.49–497.78]). High fasting plasma glucose and high LDL cholesterol similarly affected both sexes, though more prominently in males. Stroke risk attributable to alcohol use and smoking was also disproportionately greater in males (e.g., smoking-related DALYs of 128.77 [95% UI 108.86– 151.53] vs 62.76 [95% UI 52.48–75.93] in females). While dietary risk factors showed smaller differential effects by sex, males generally had slightly higher attributable burdens (Supplementary Table 4).

This male–female pattern persisted across all subregions and most countries; for instance, in southern Latin America, males experienced 619.75 DALYs [95% UI 458.97–756.94] per 100,000 from high systolic blood pressure compared with 417.12 DALYs [95% UI 308.99–513.95] in females (Supplementary Table 5).

Among age groups, older adults (≥70 years) consistently exhibited the highest stroke-related deaths and DALYs, followed by those aged 50–69 years, with substantially lower burdens in younger groups regionwide (Figure 3, Supplementary Table 6, Supplement Figure 7-12) and at country level (Supplementary Table 7, Supplement Figure 13-51).

**Figure 3:**
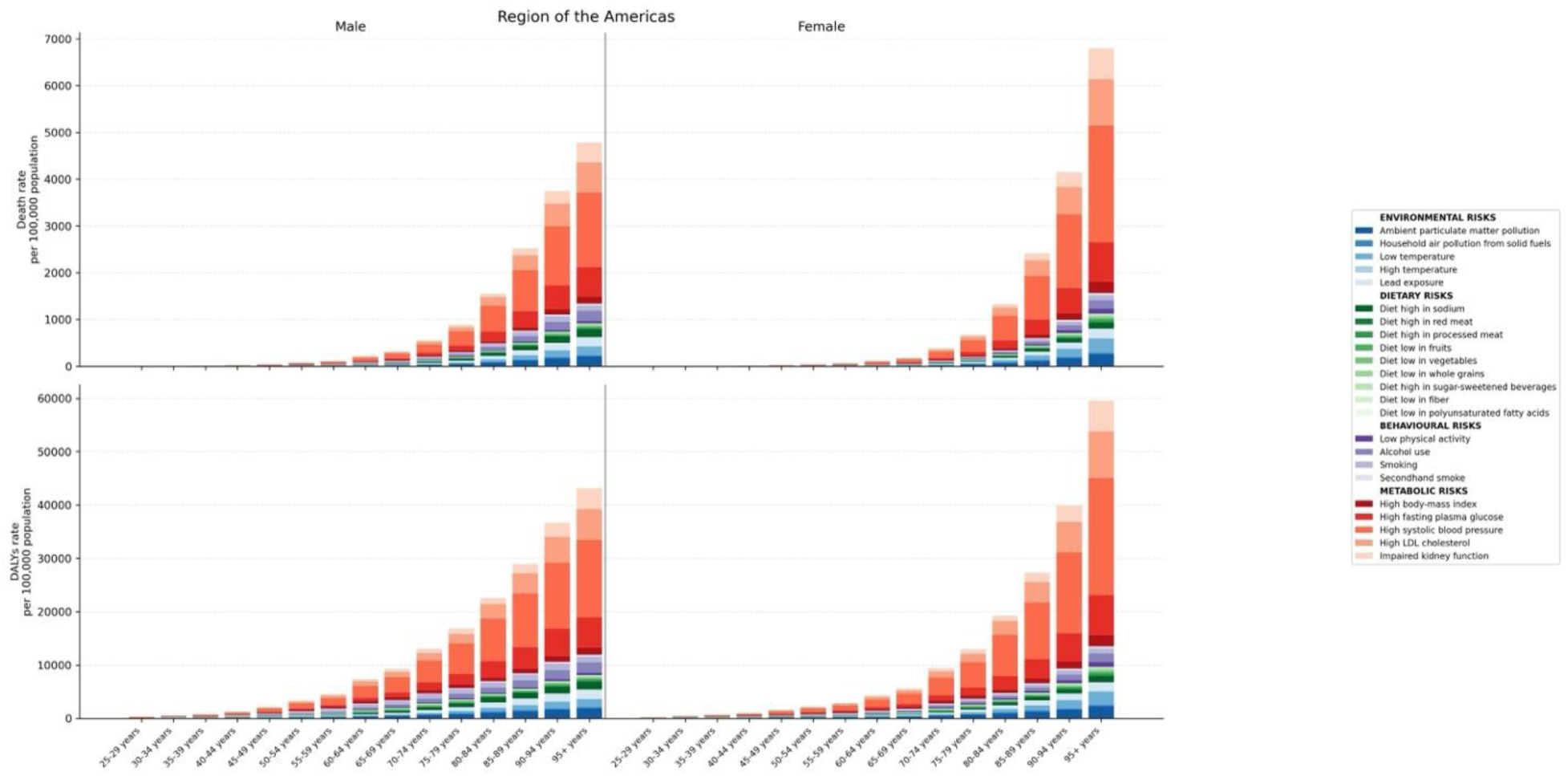
Age- and Sex-Specific Stroke Burden Attributable to Risk Factors in the Americas, 2021. **Legend:** This stacked bar chart illustrates the age- and sex-specific burden of stroke, measured in deaths and DALYs per 100,000 population in the Americas for 2021. The data are split by male (left) and female (right), with different colors representing various risk factors categorized into environmental, dietary, behavioral, and metabolic groups.

### 2. Temporal Trends in Stroke Burden (1990–2021)

#### 2.1. Overall Changes in Deaths and DALYs

Age-standardized deaths and DALYs rates from stroke attributable to most risk factors declined between 1990 and 2021. Household air pollution from solid fuels showed the steepest reduction in both deaths (AAPC −5.09% [95% CI −5.17 to −5.02]; p<0.001) and DALYs (AAPC −4.92% [95% CI −4.98 to −4.86]; p<0.001). Stroke deaths and DALYs attributable to smoke also declined sharply (AAPC around −3.30%; p<0.001). High systolic blood pressure-related effects exhibited a more moderate decrease (deaths: AAPC −1.89% [95% CI −1.93 to −1.85]; DALYs: AAPC −1.84% [95% CI −1.89 to −1.80]; both p<0.001). High body-mass index and high fasting plasma glucose demonstrated smaller reductions in attributable deaths (AAPC −0.54% [95% CI −0.59 to −0.49] and −1.10%l [95% CI −1.14 to −1.06], respectively) and DALYs (AAPC −0.42% [95% CI −0.46 to −0.37] and −0.99% [95% CI −1.05 to −0.93], respectively; all p<0.001). (Table 3)

**Table 3:**
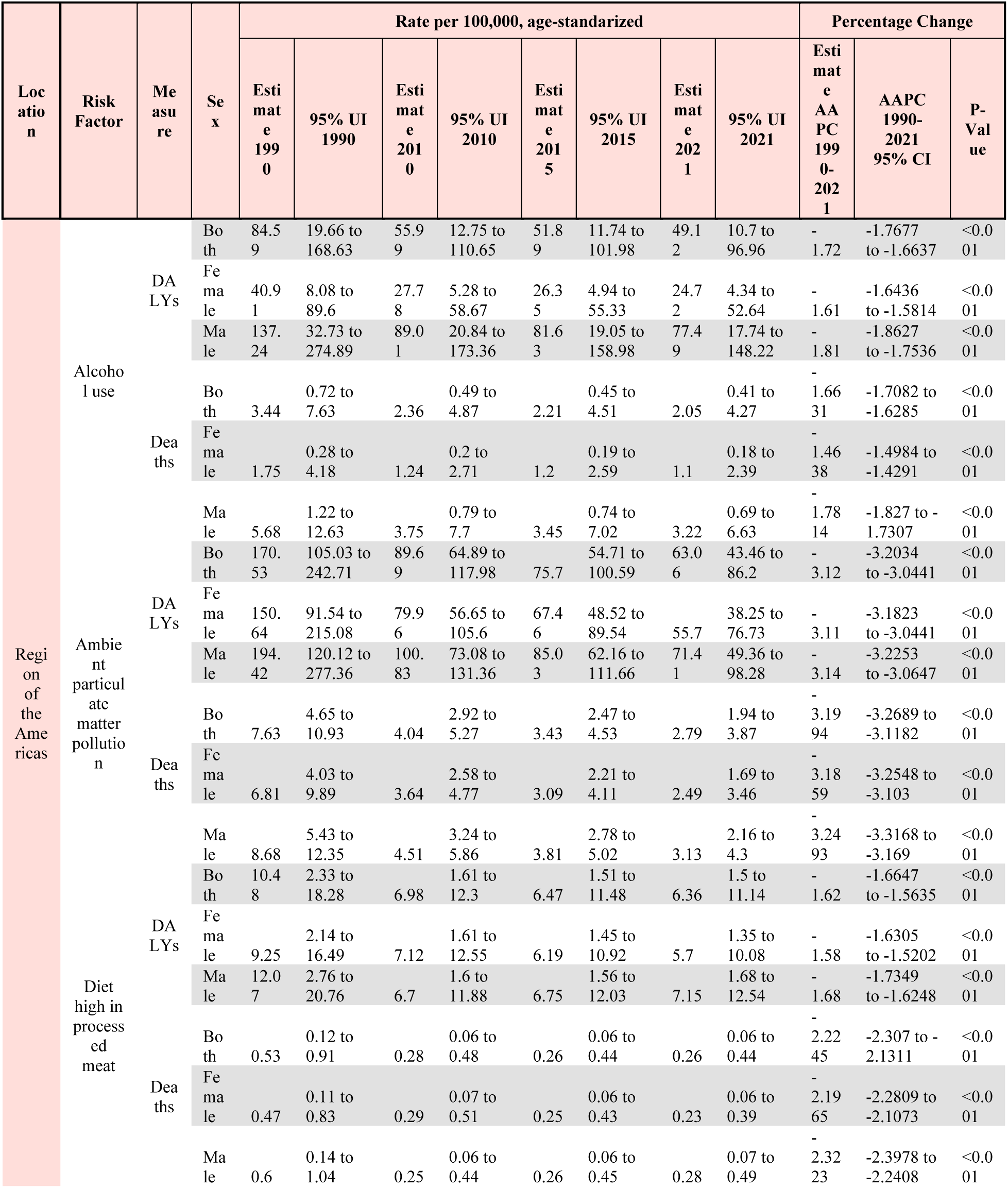
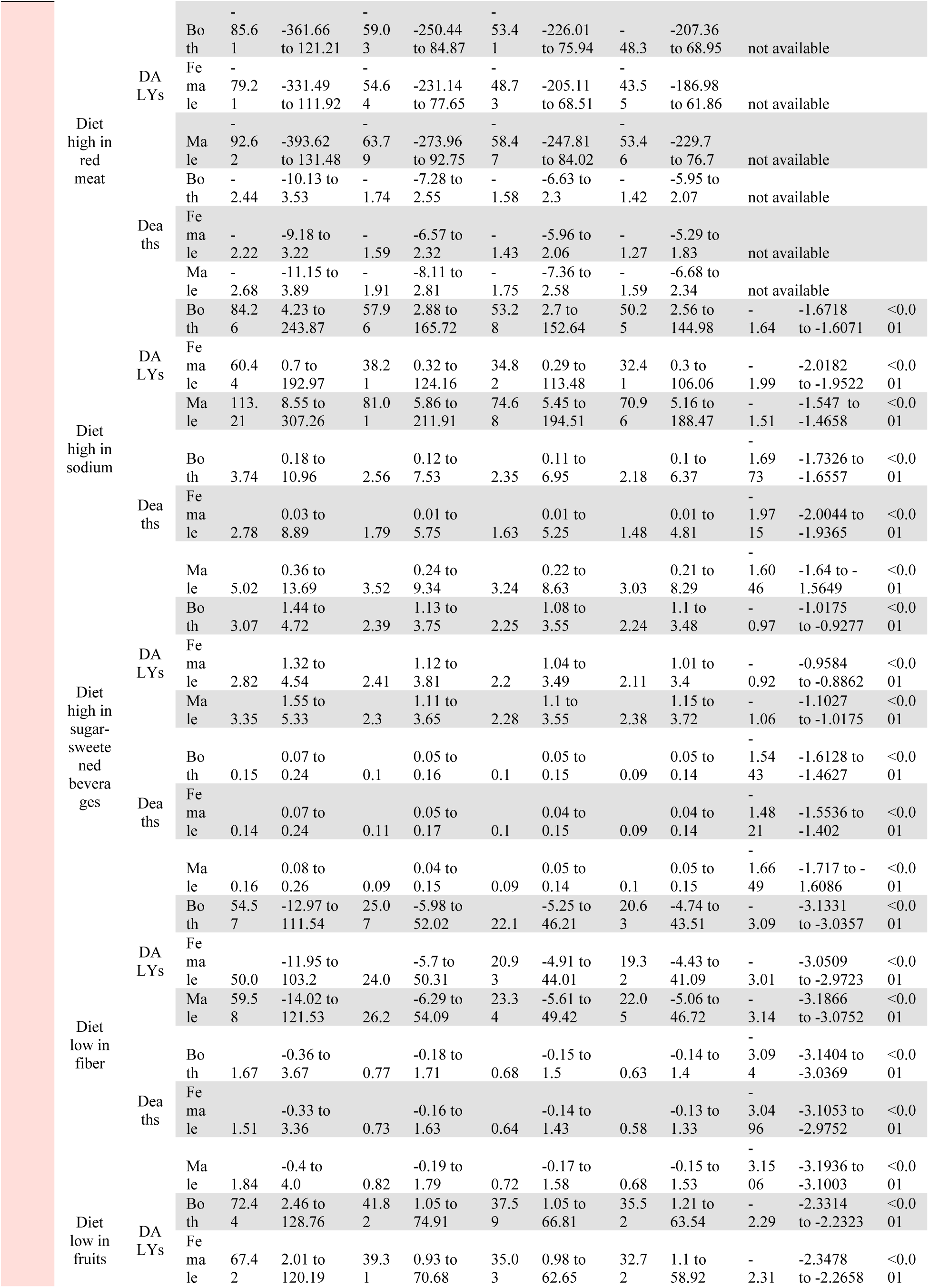
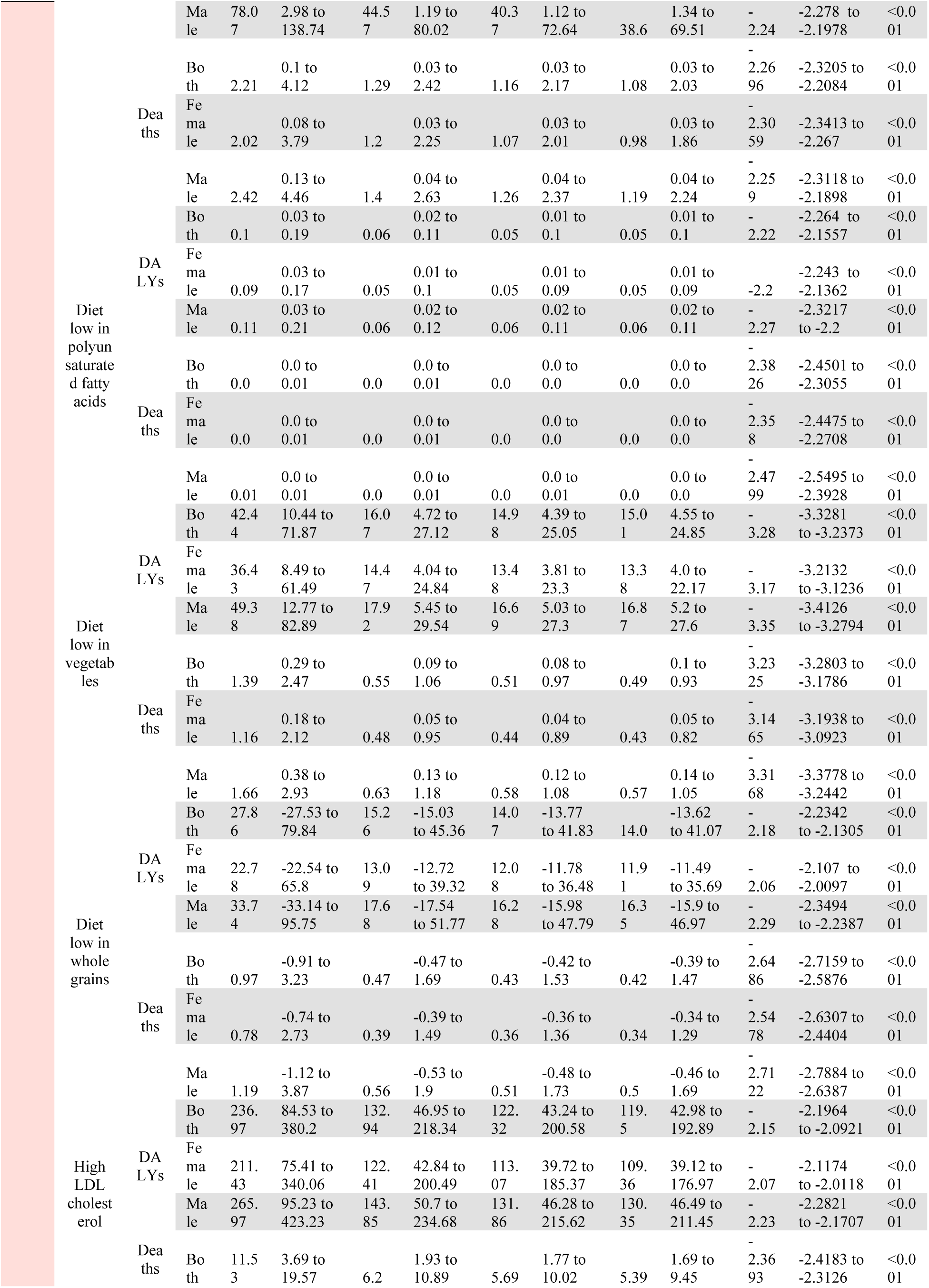
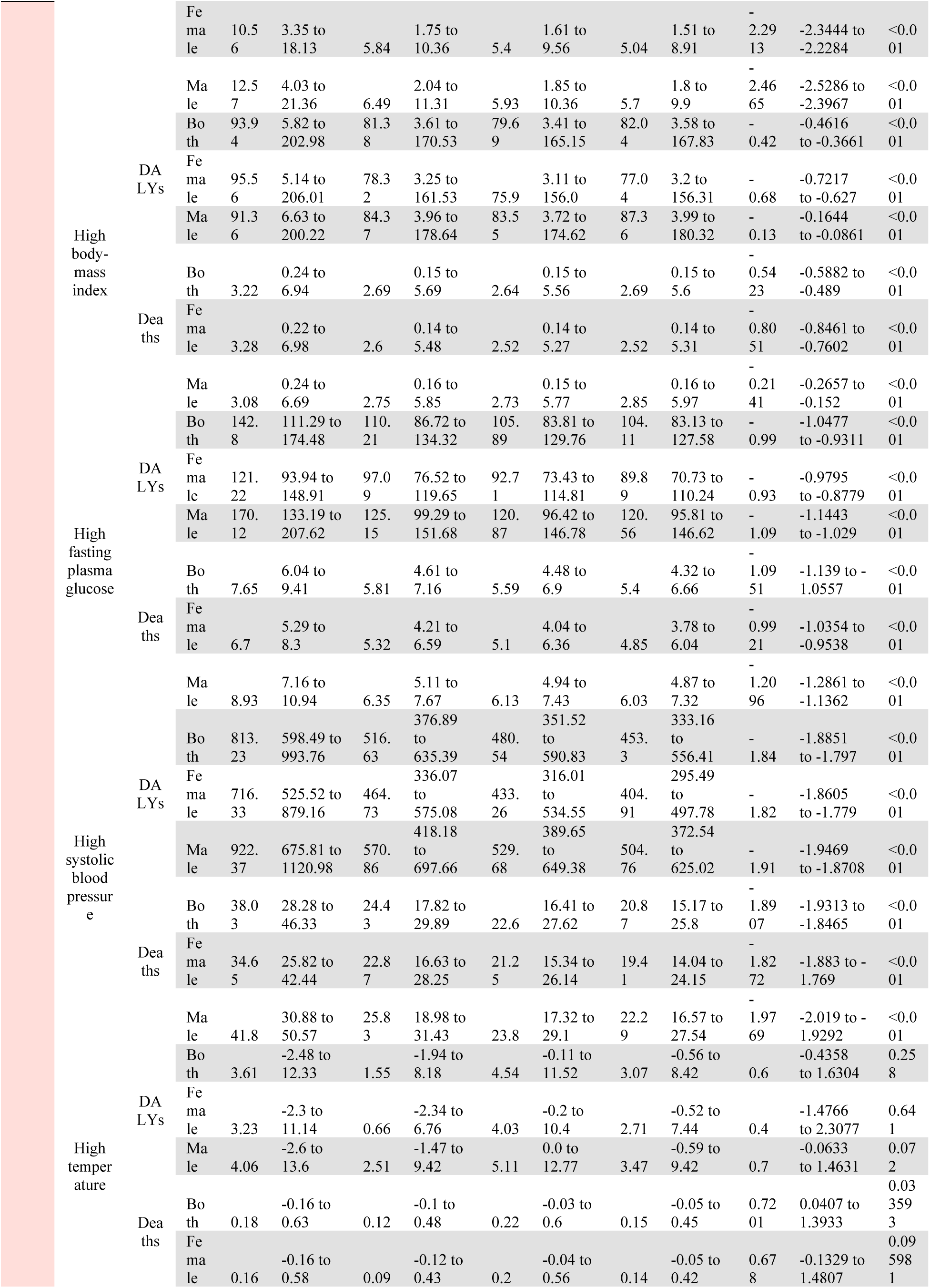
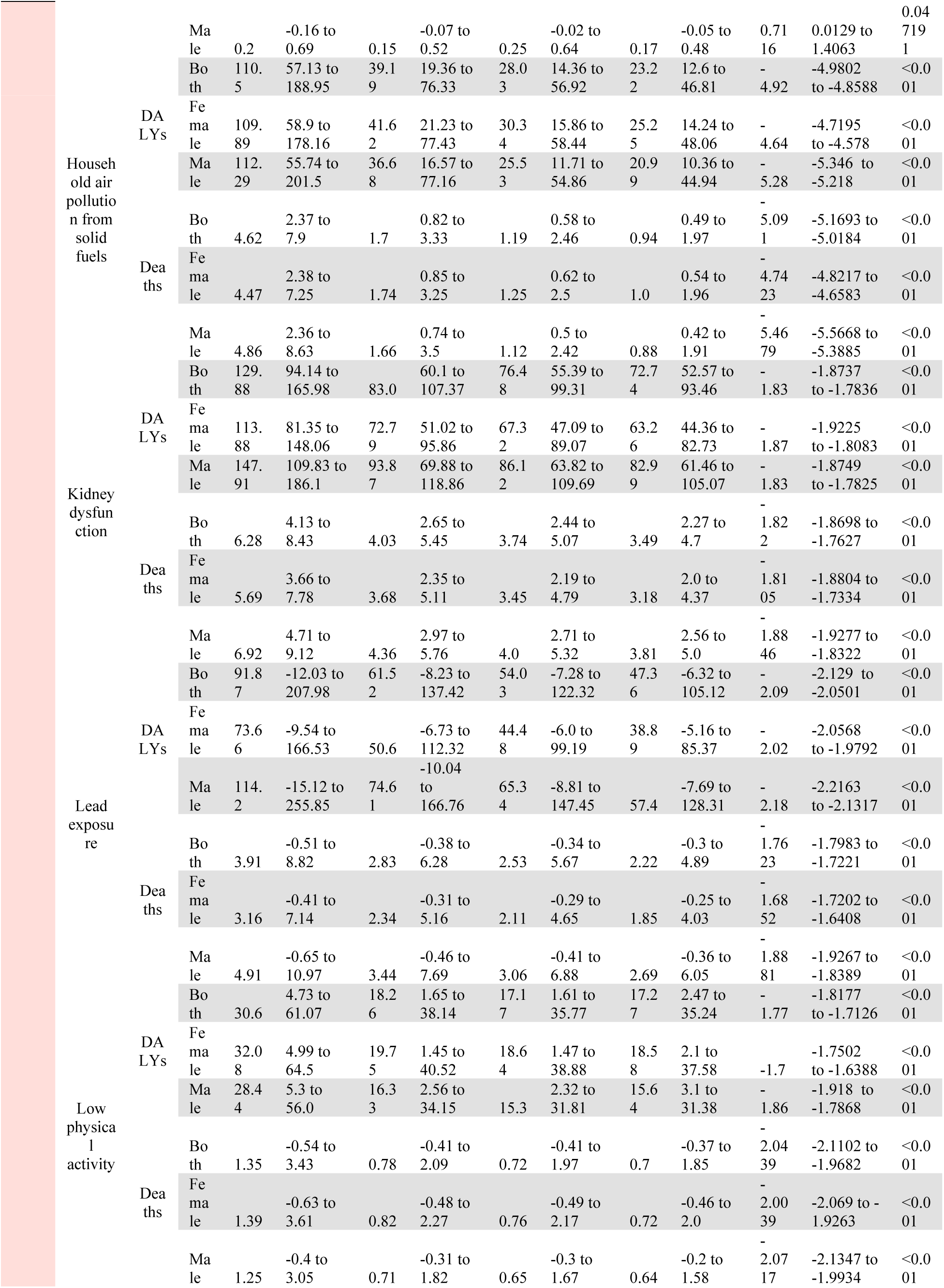
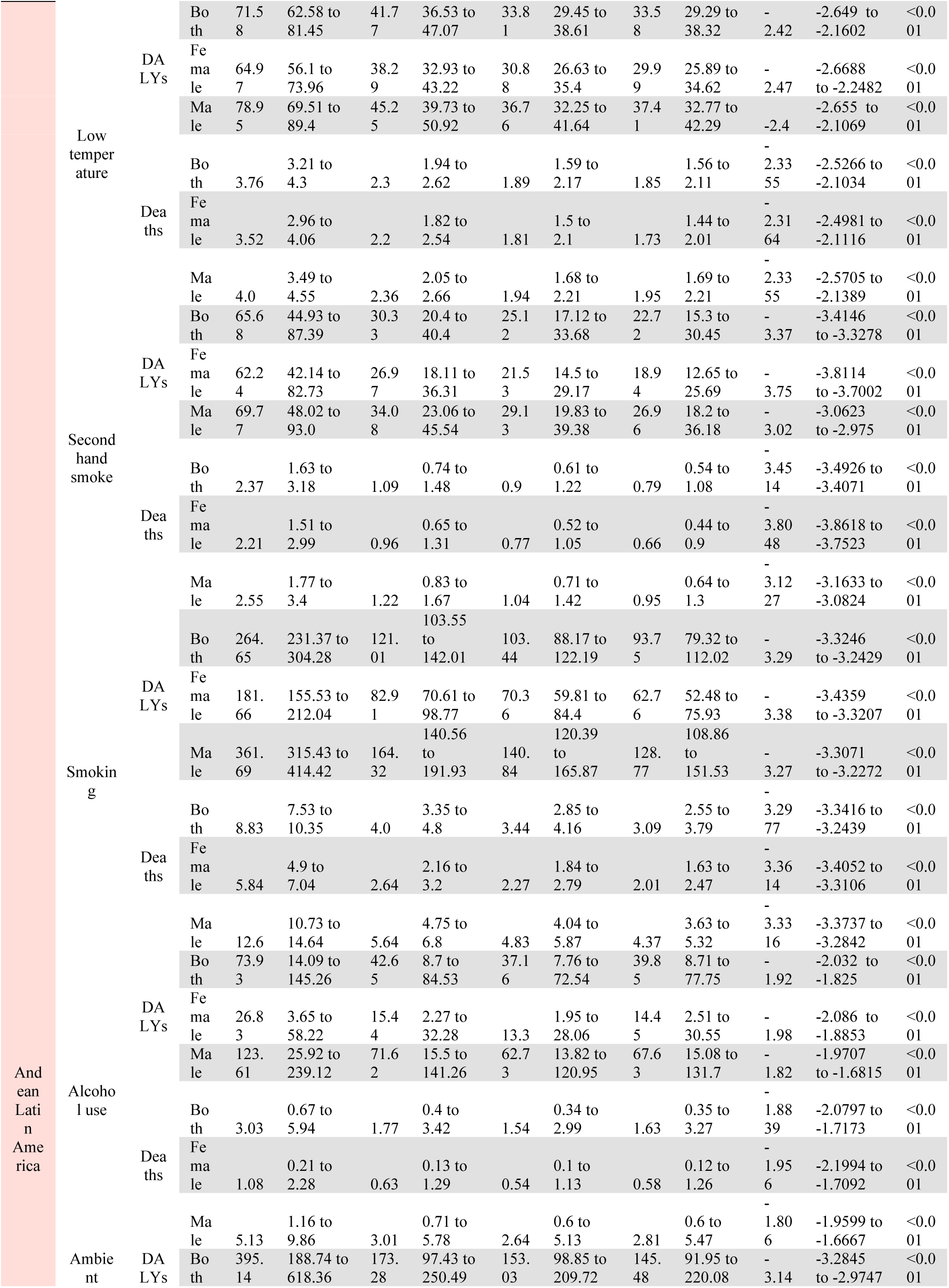
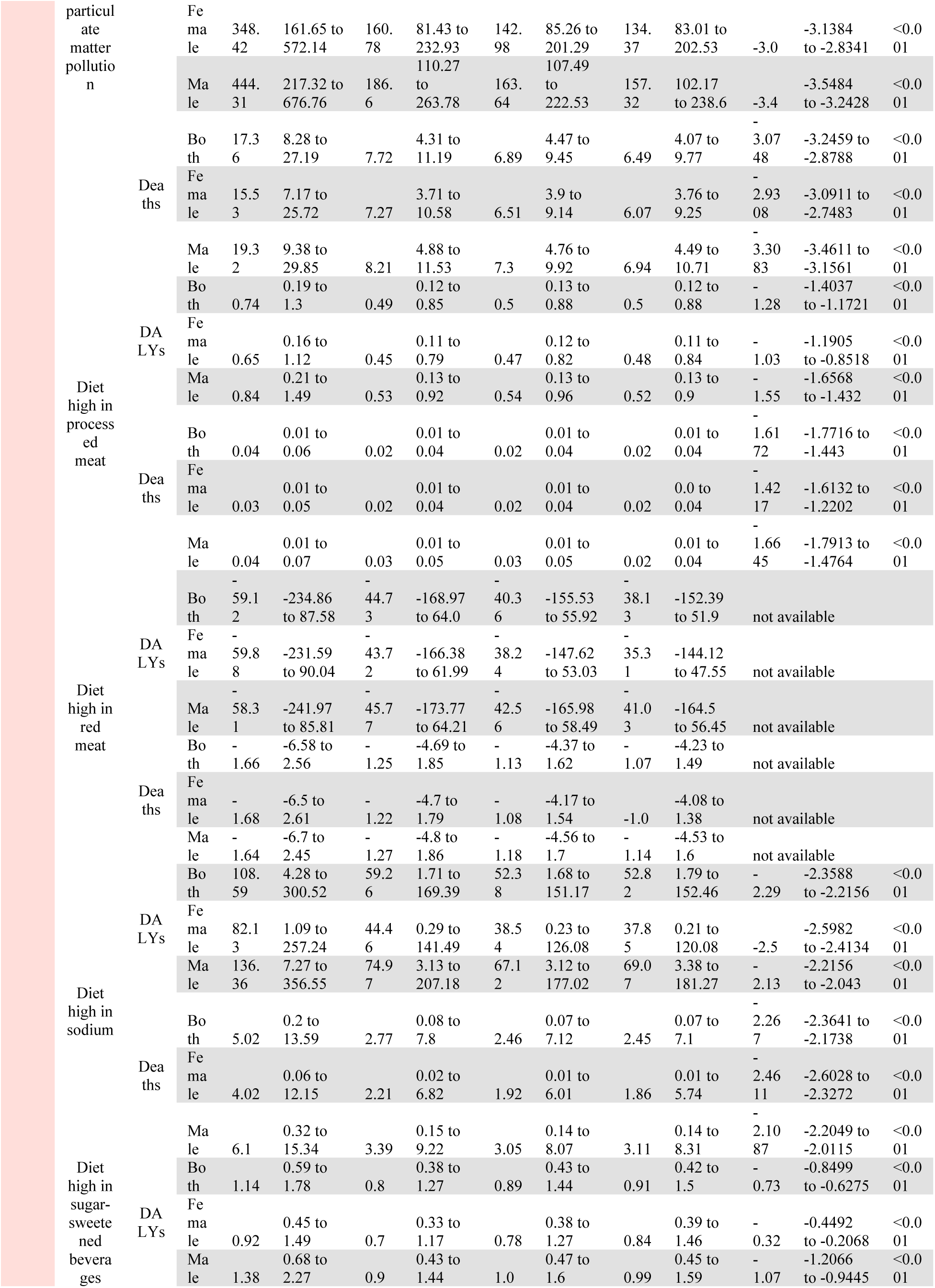
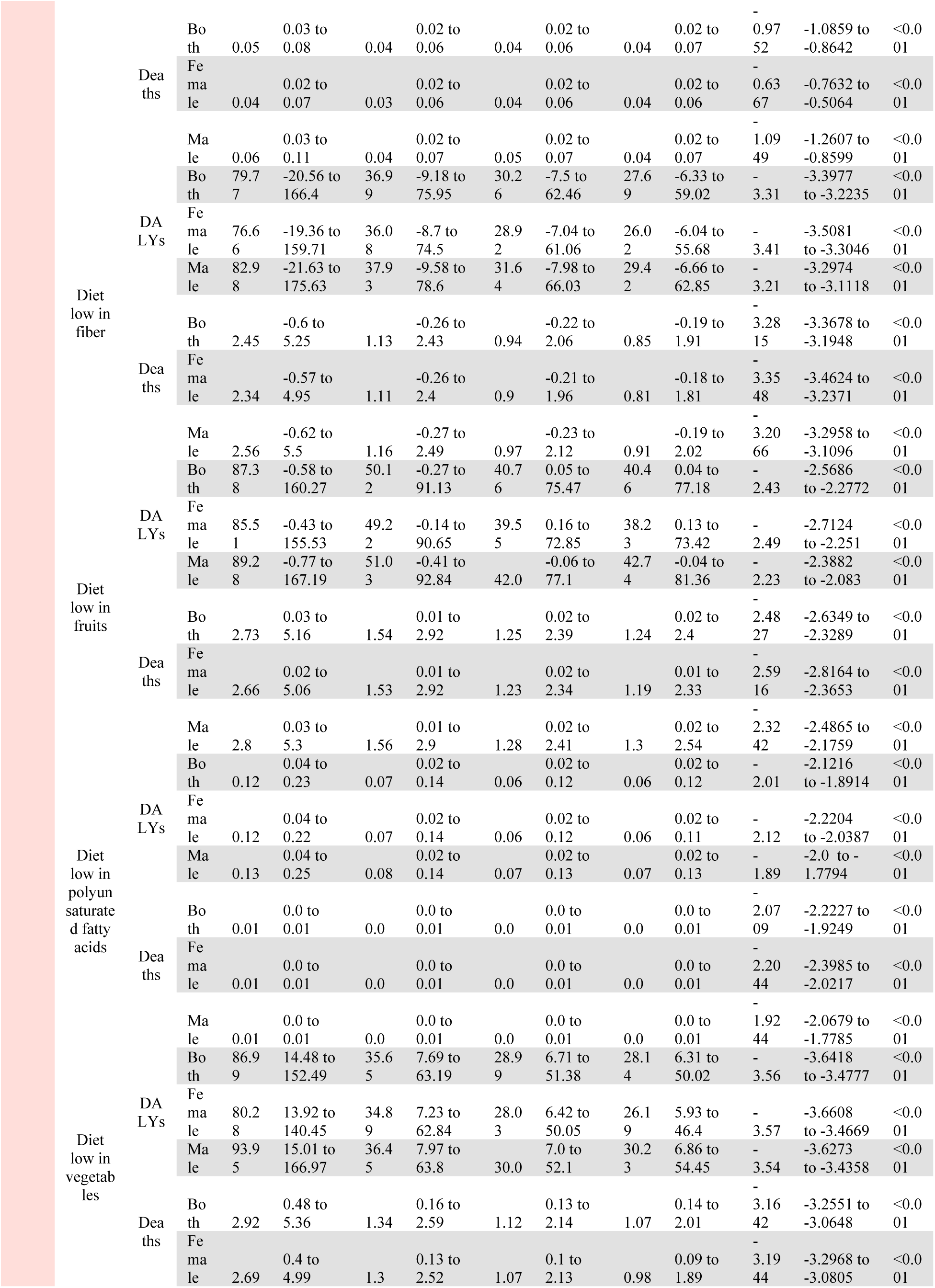
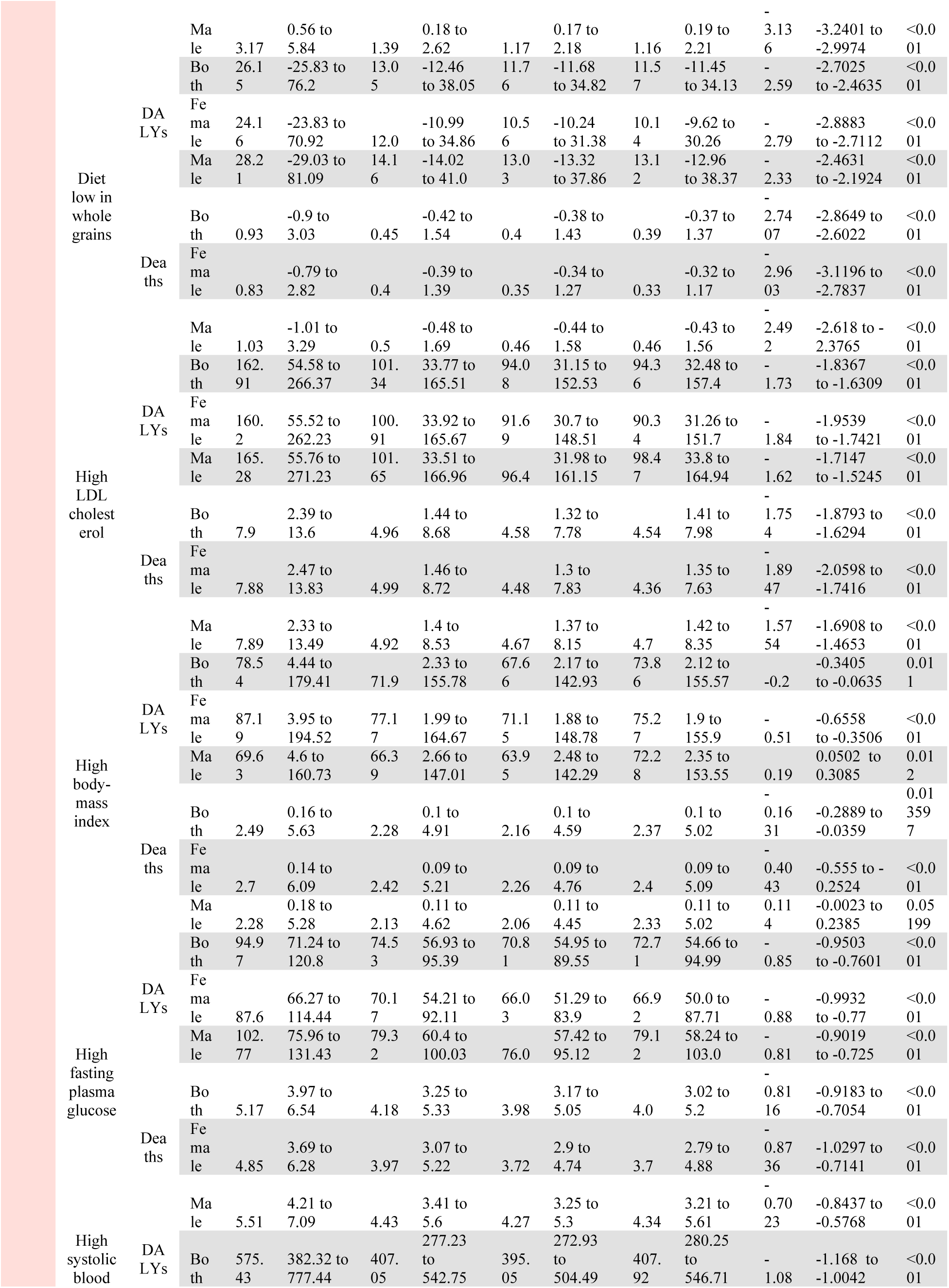
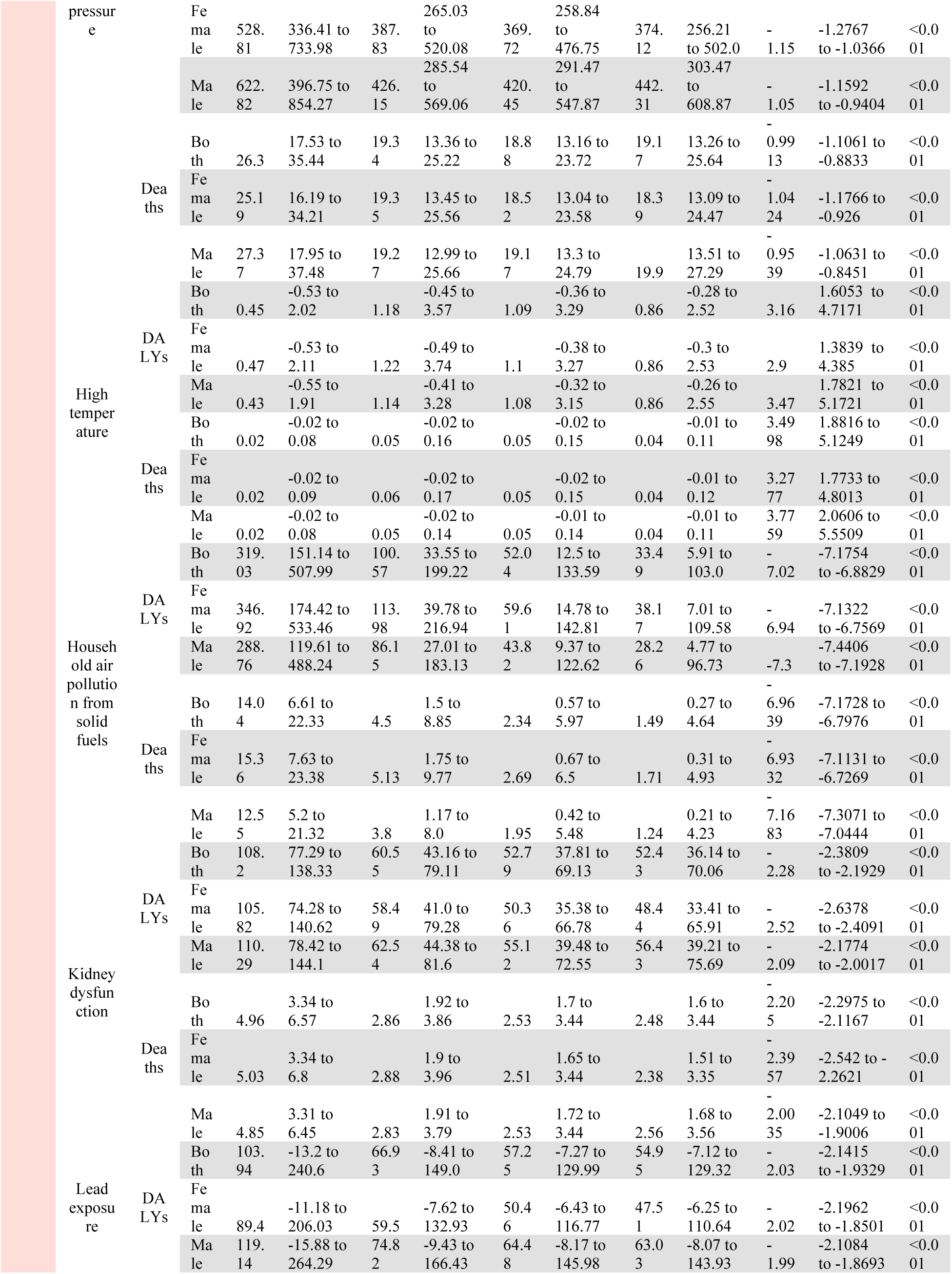
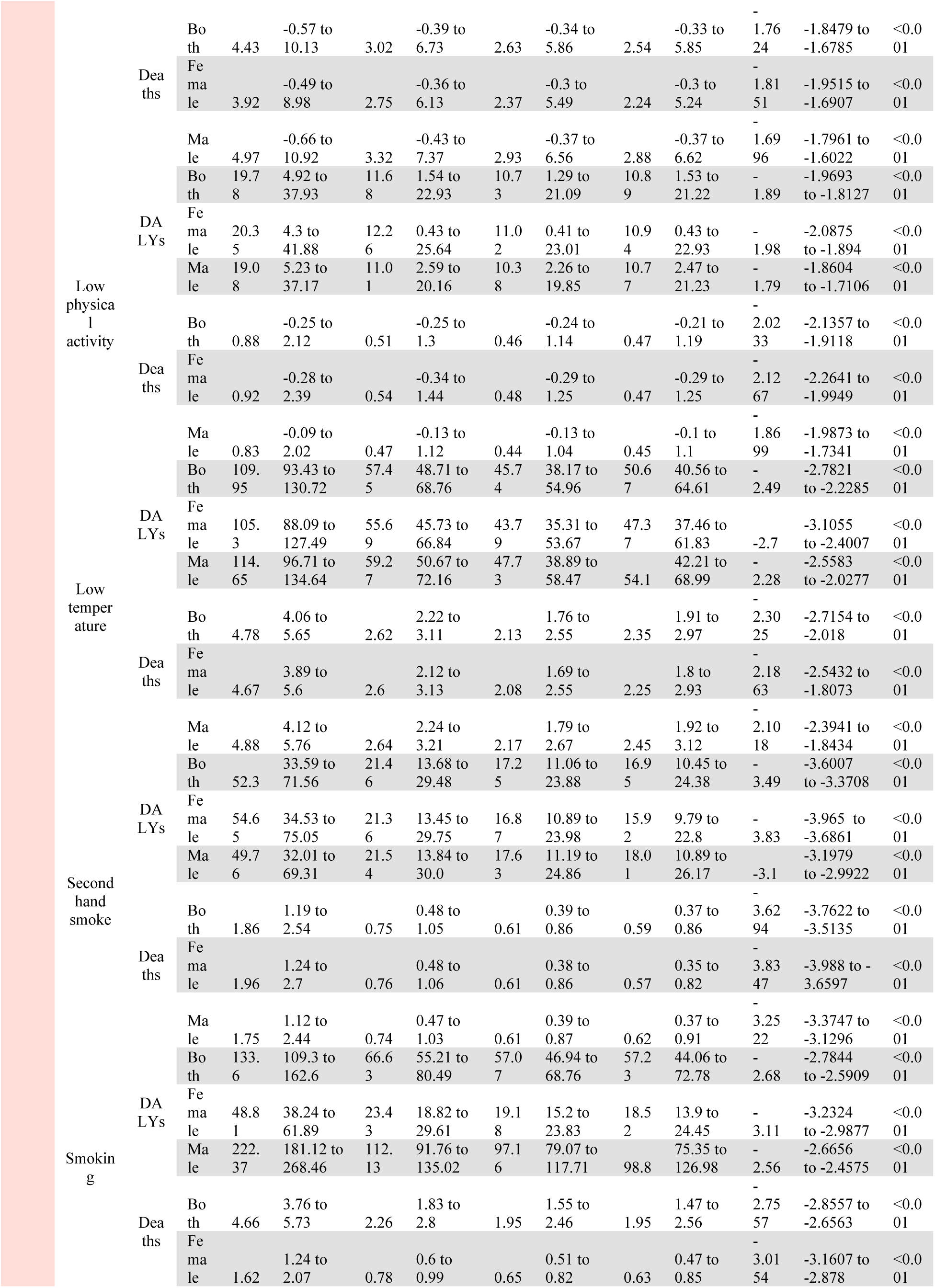
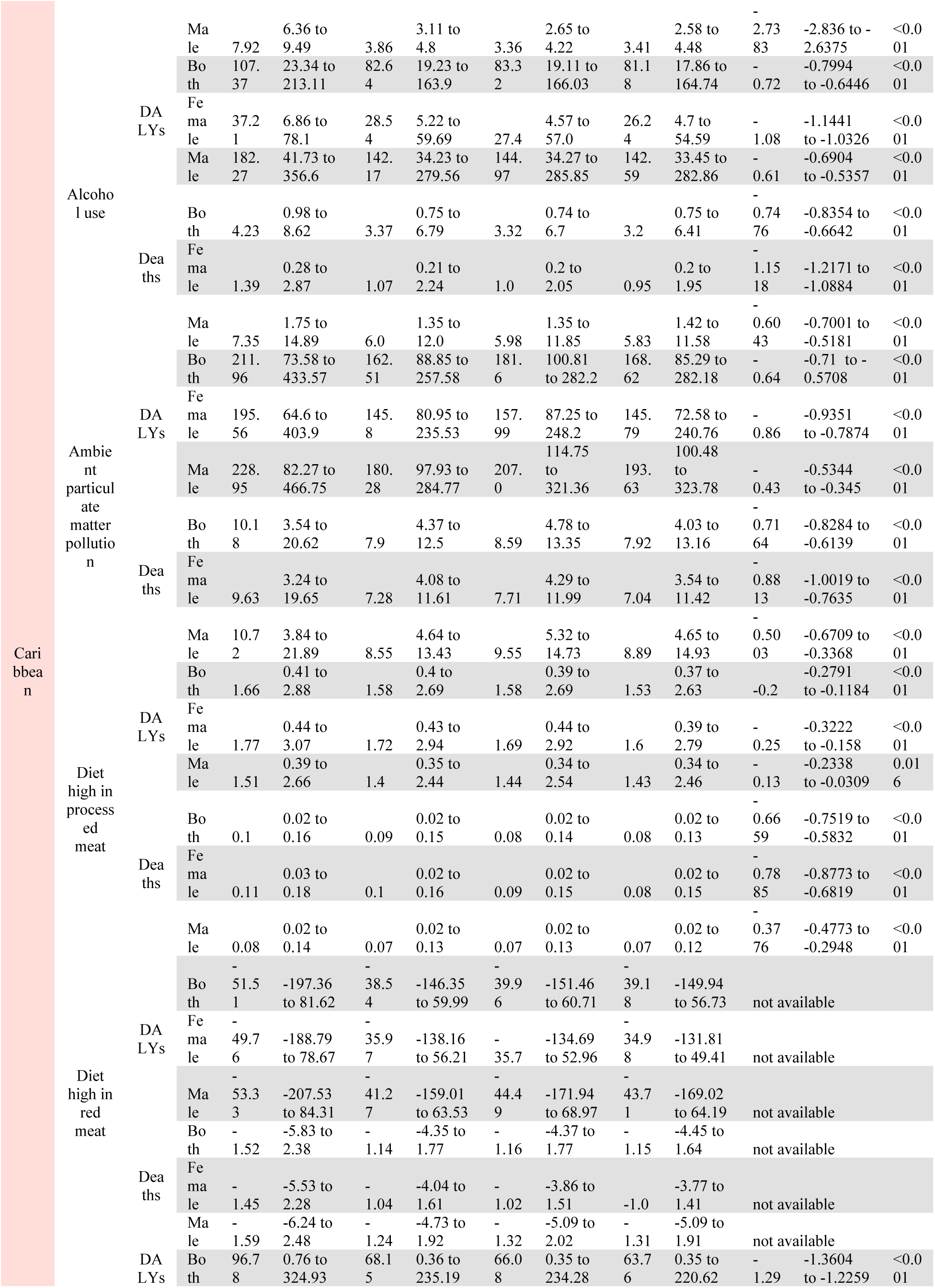
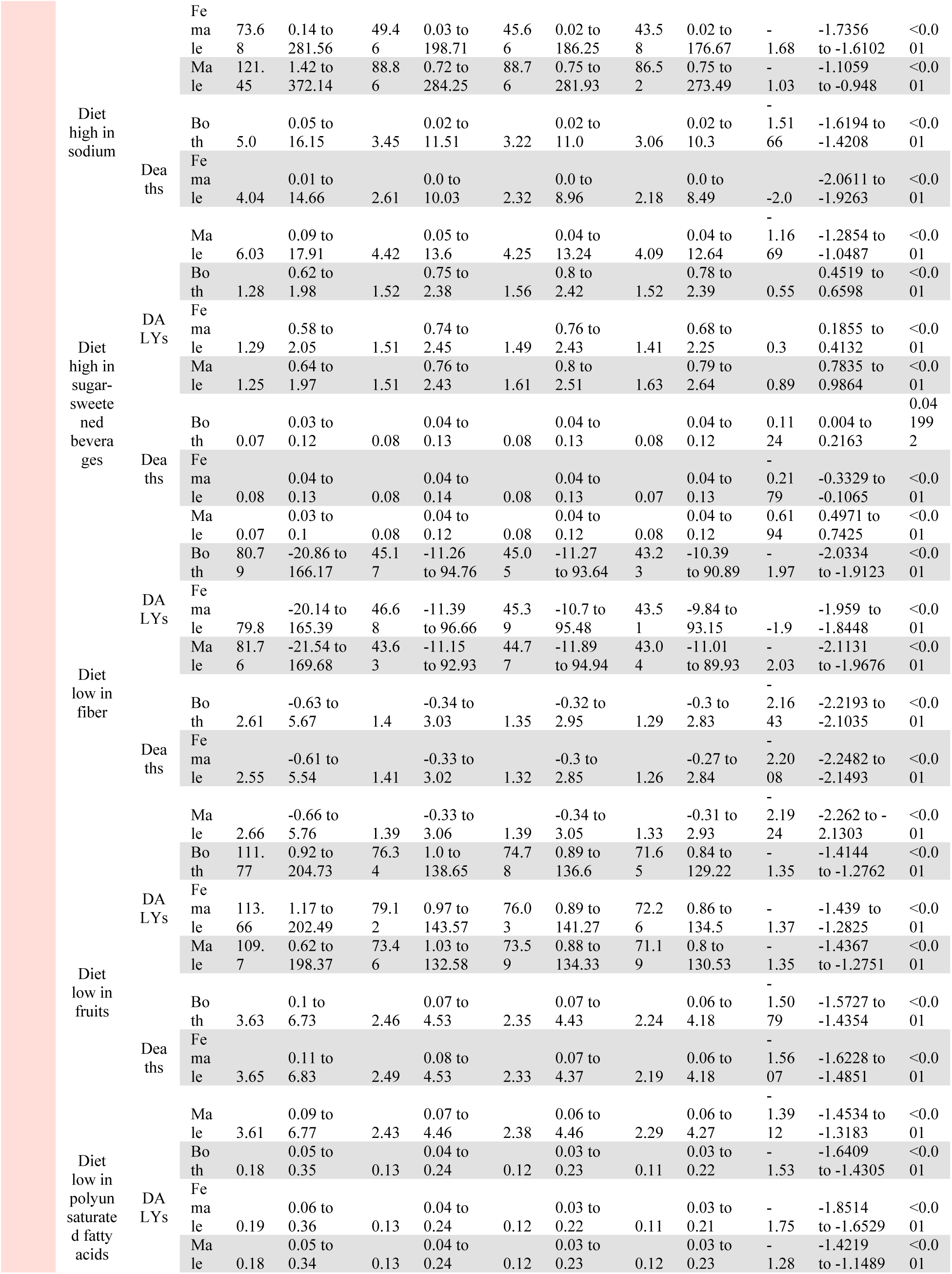
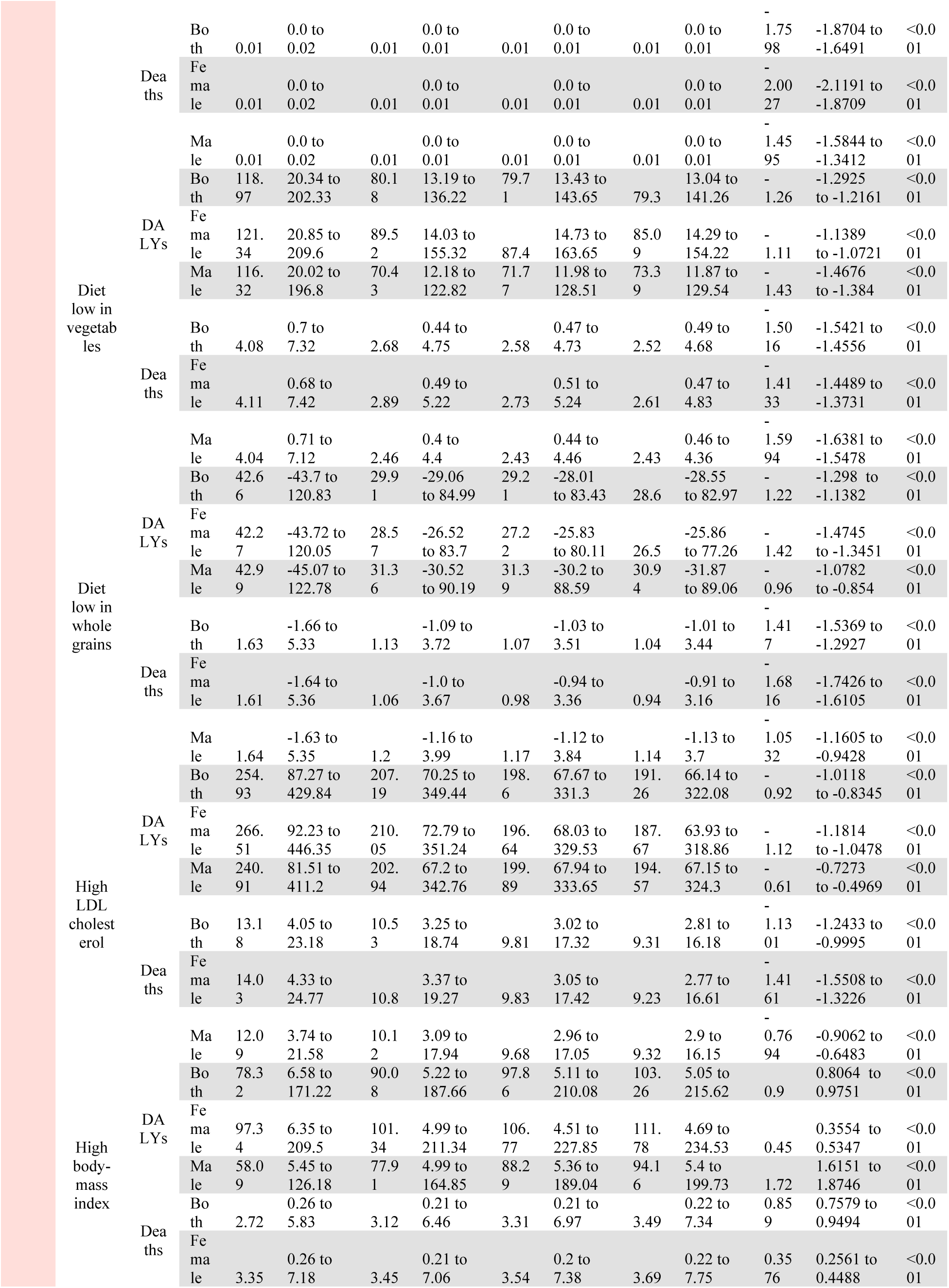
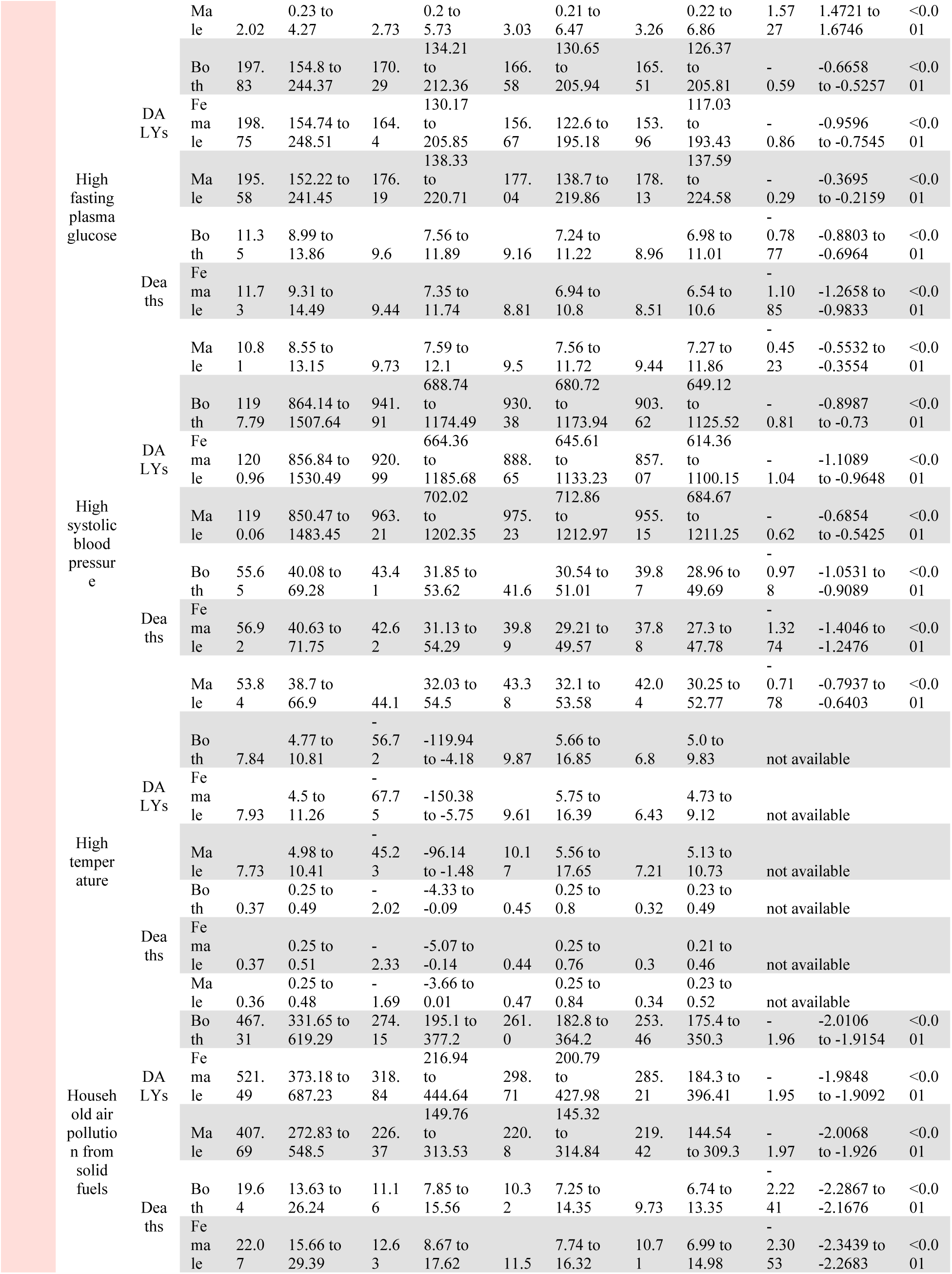
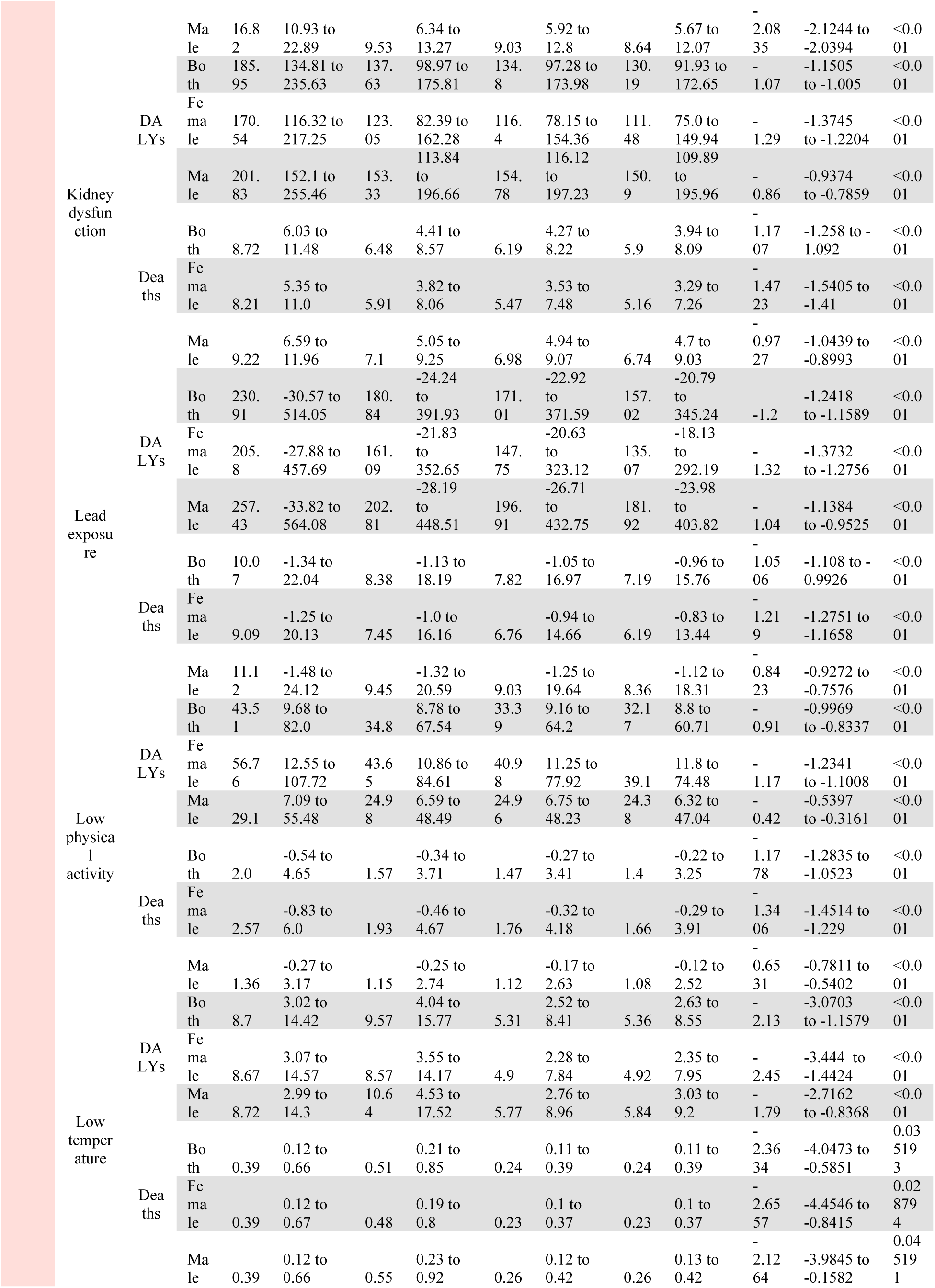
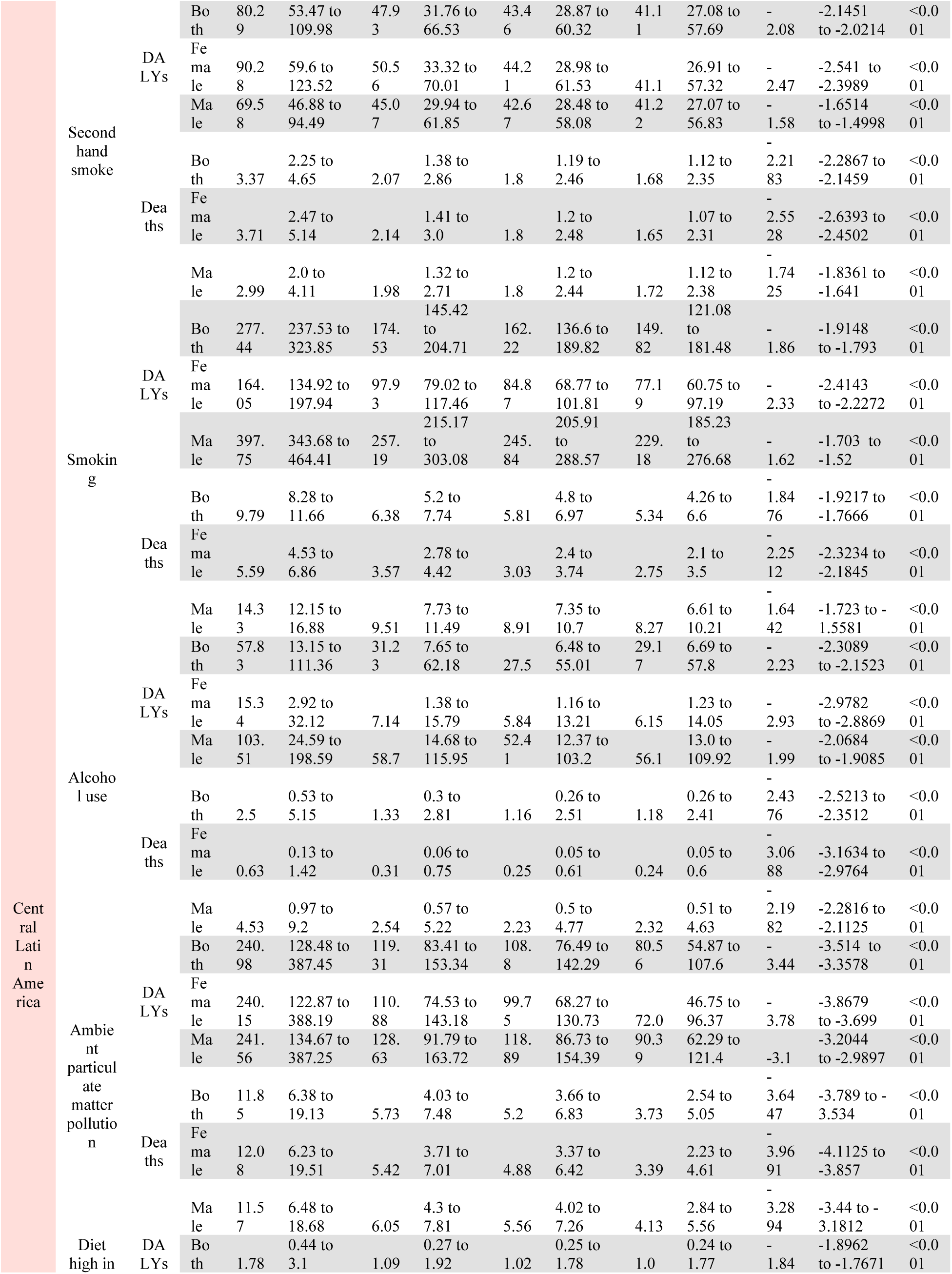
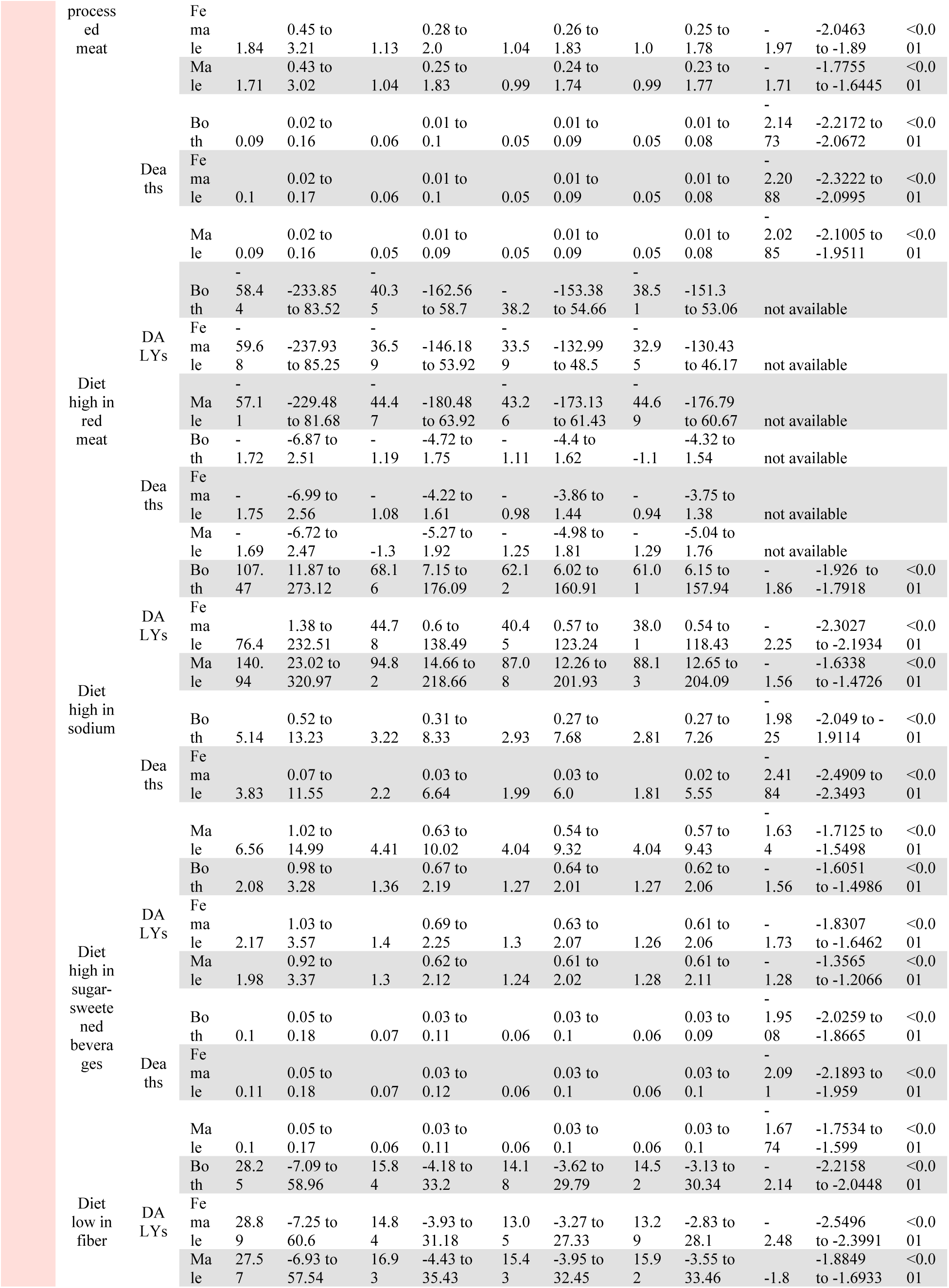
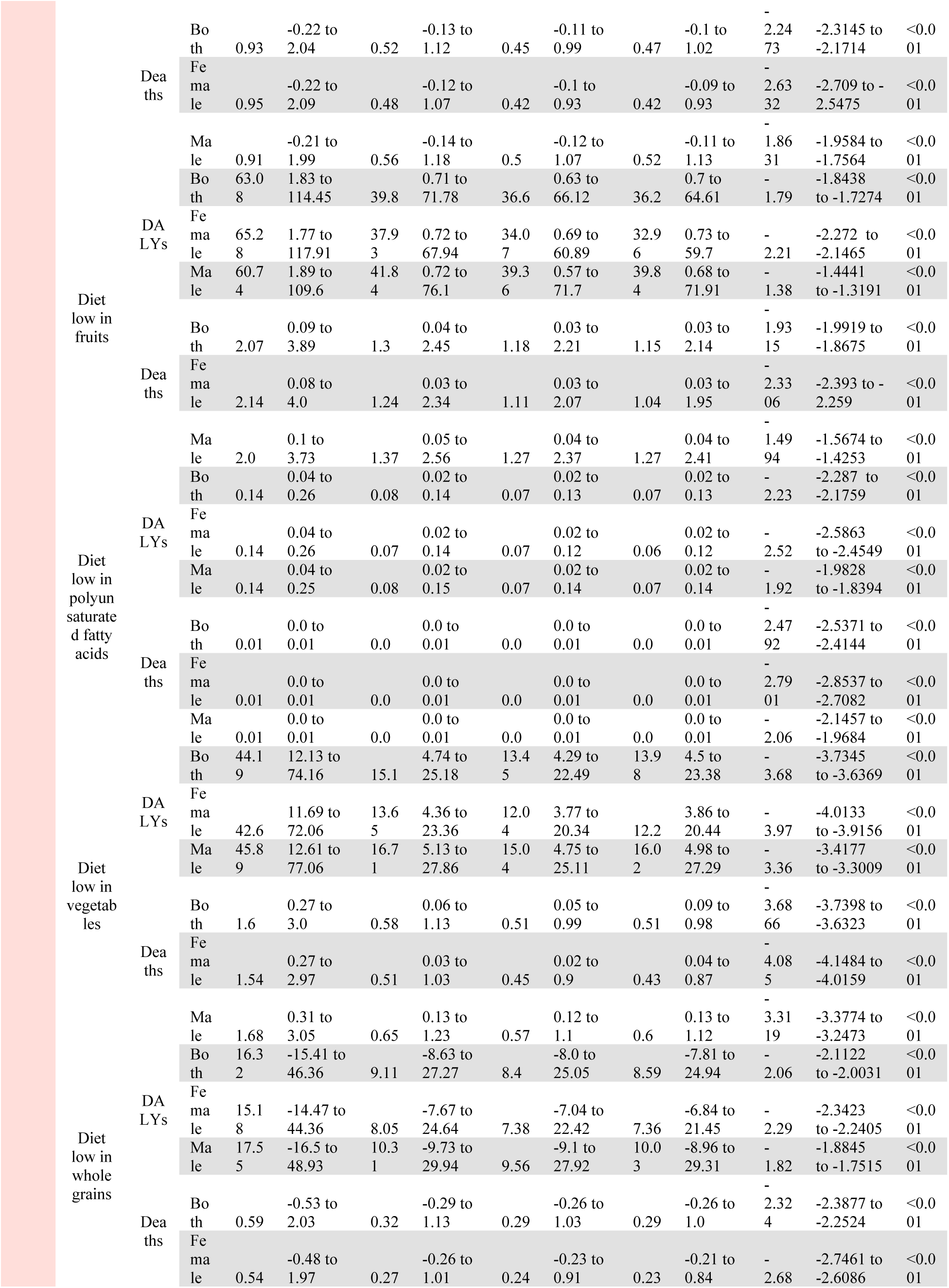
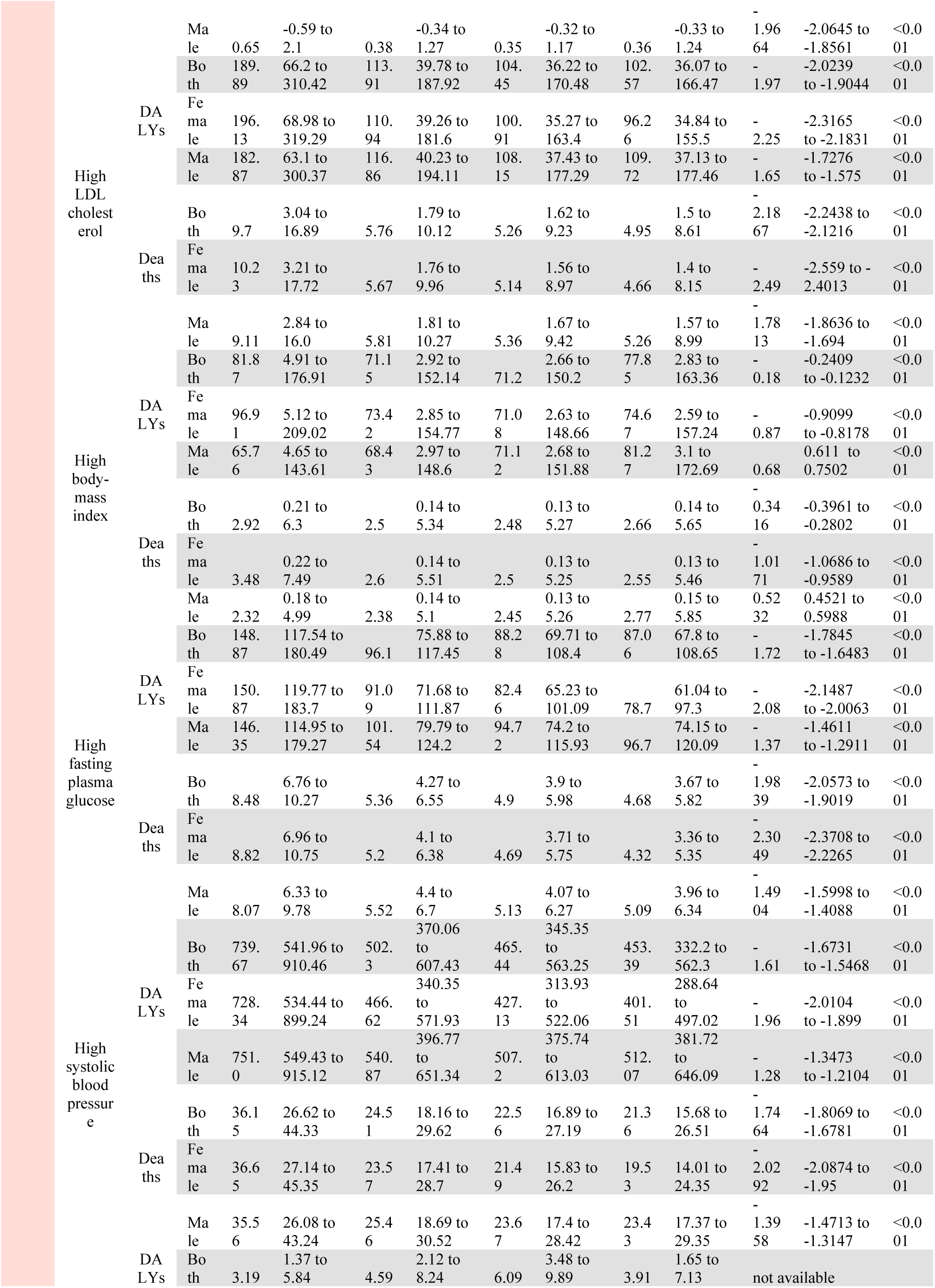
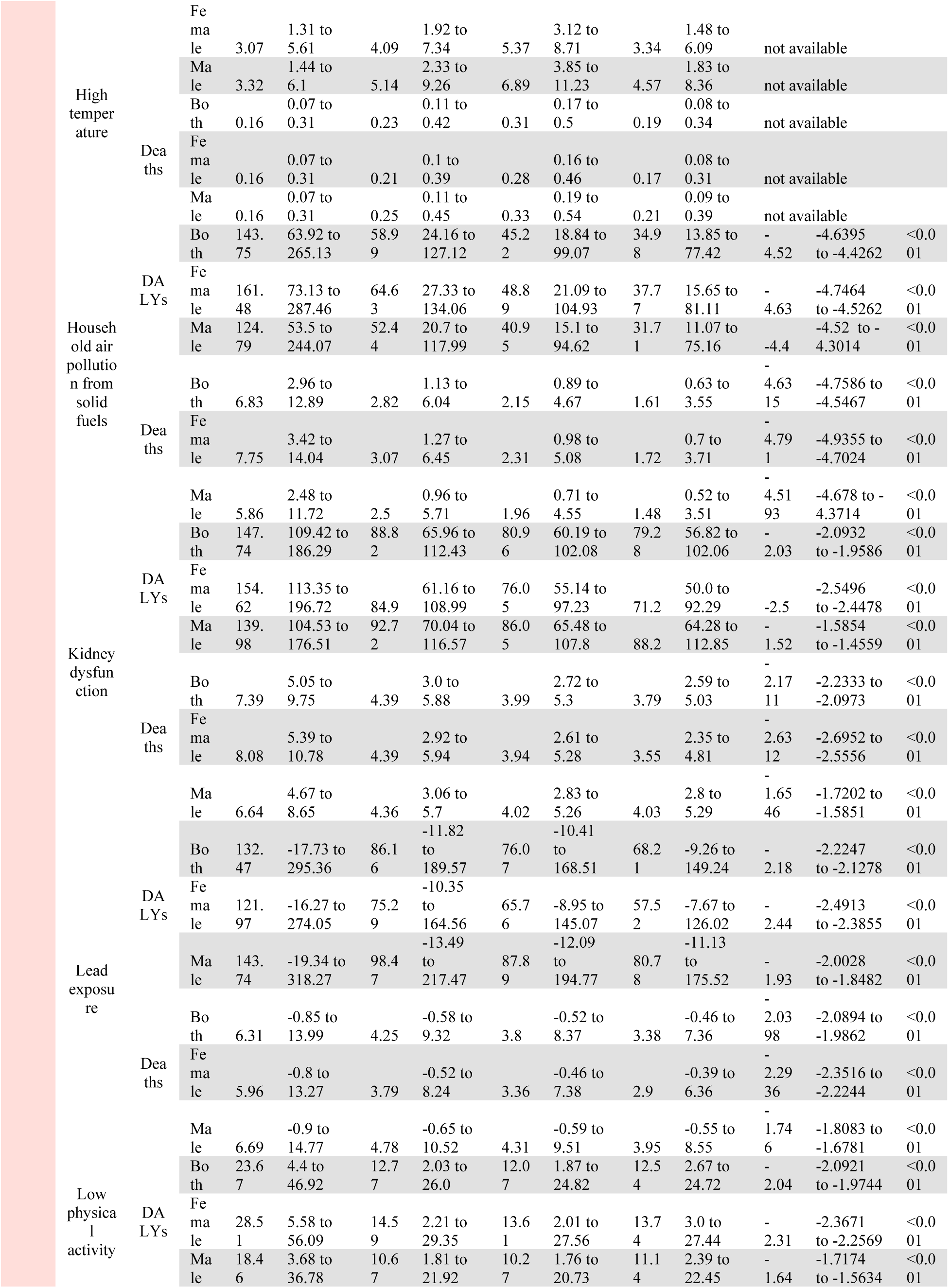
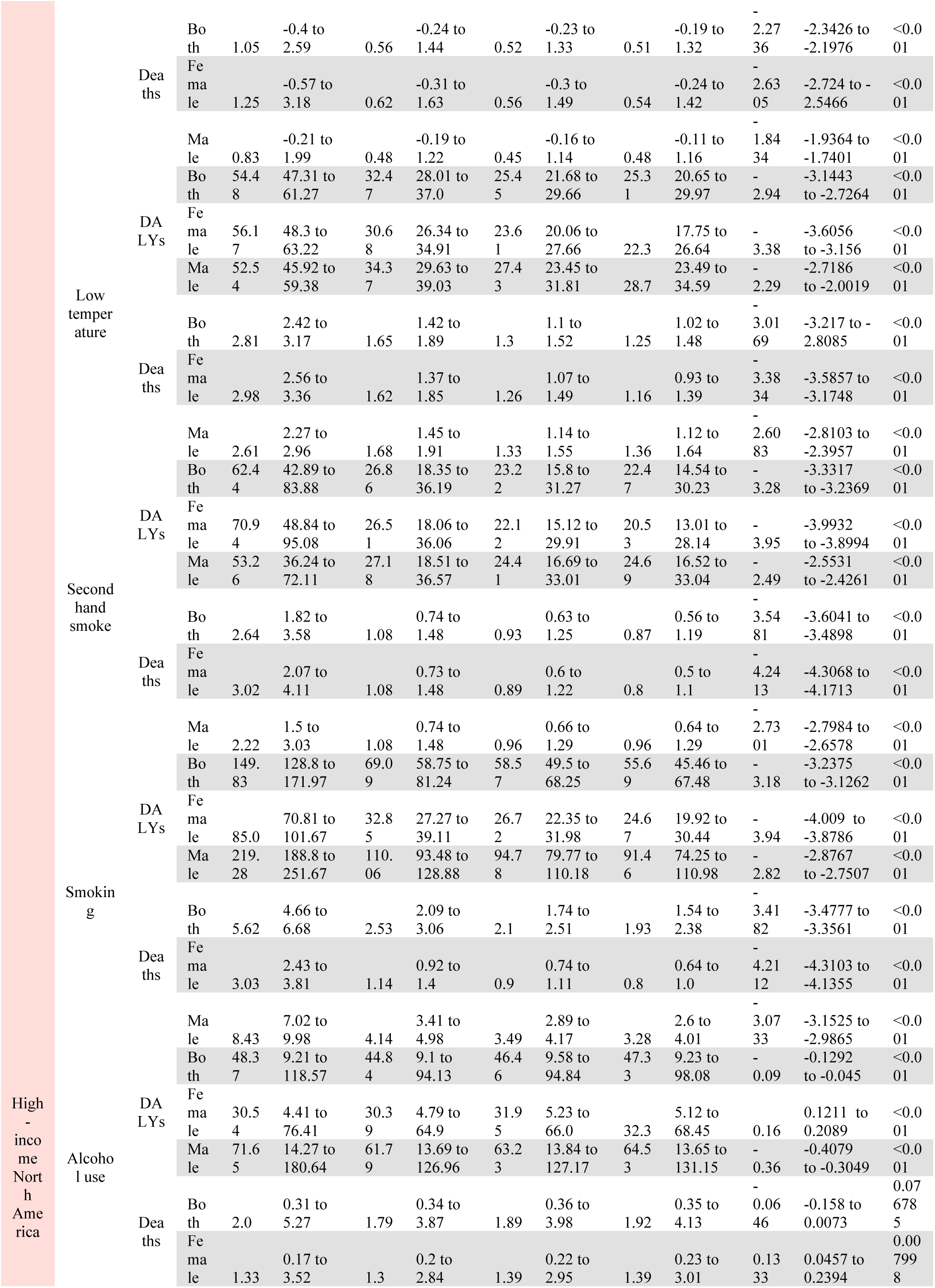
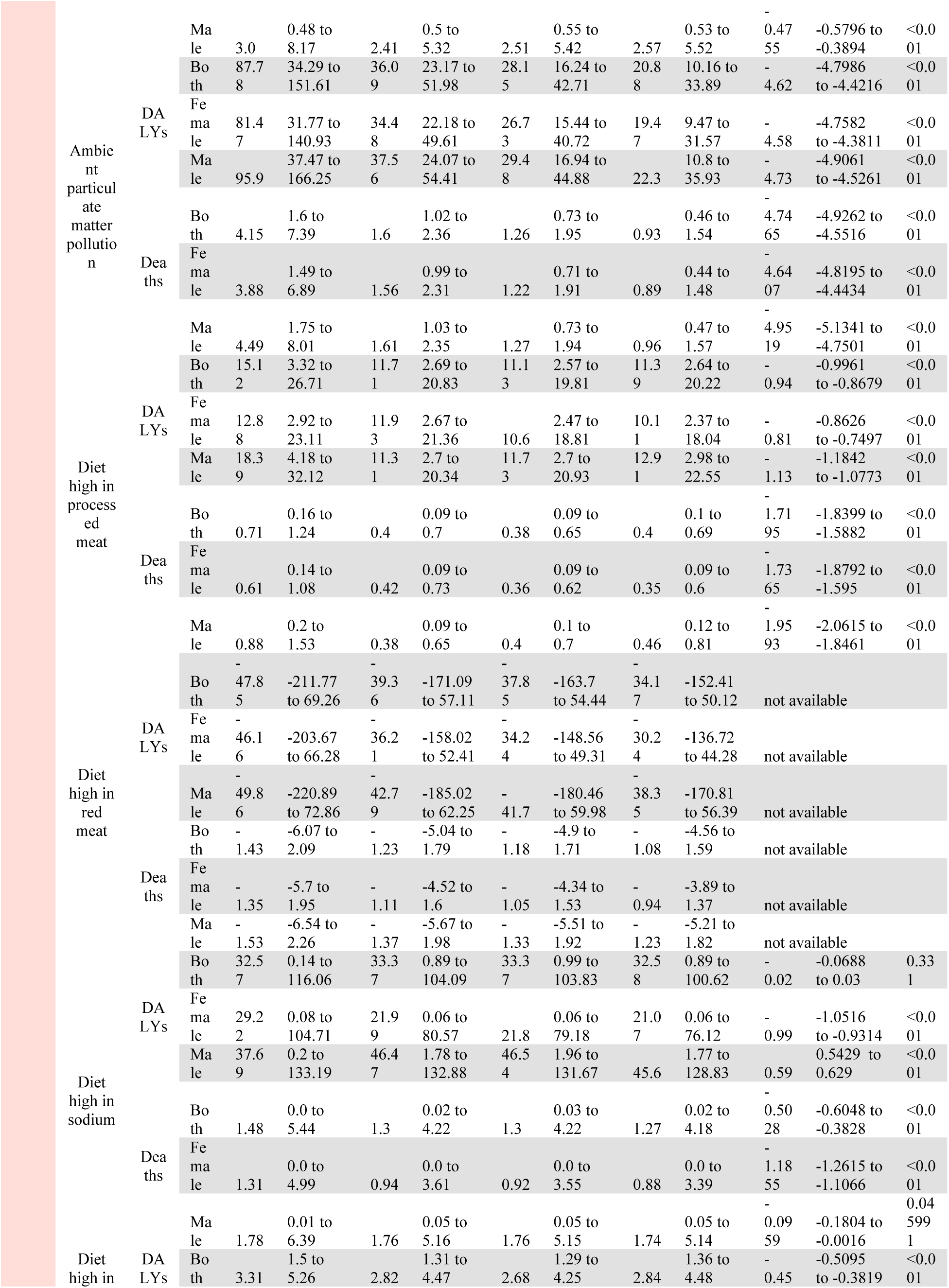
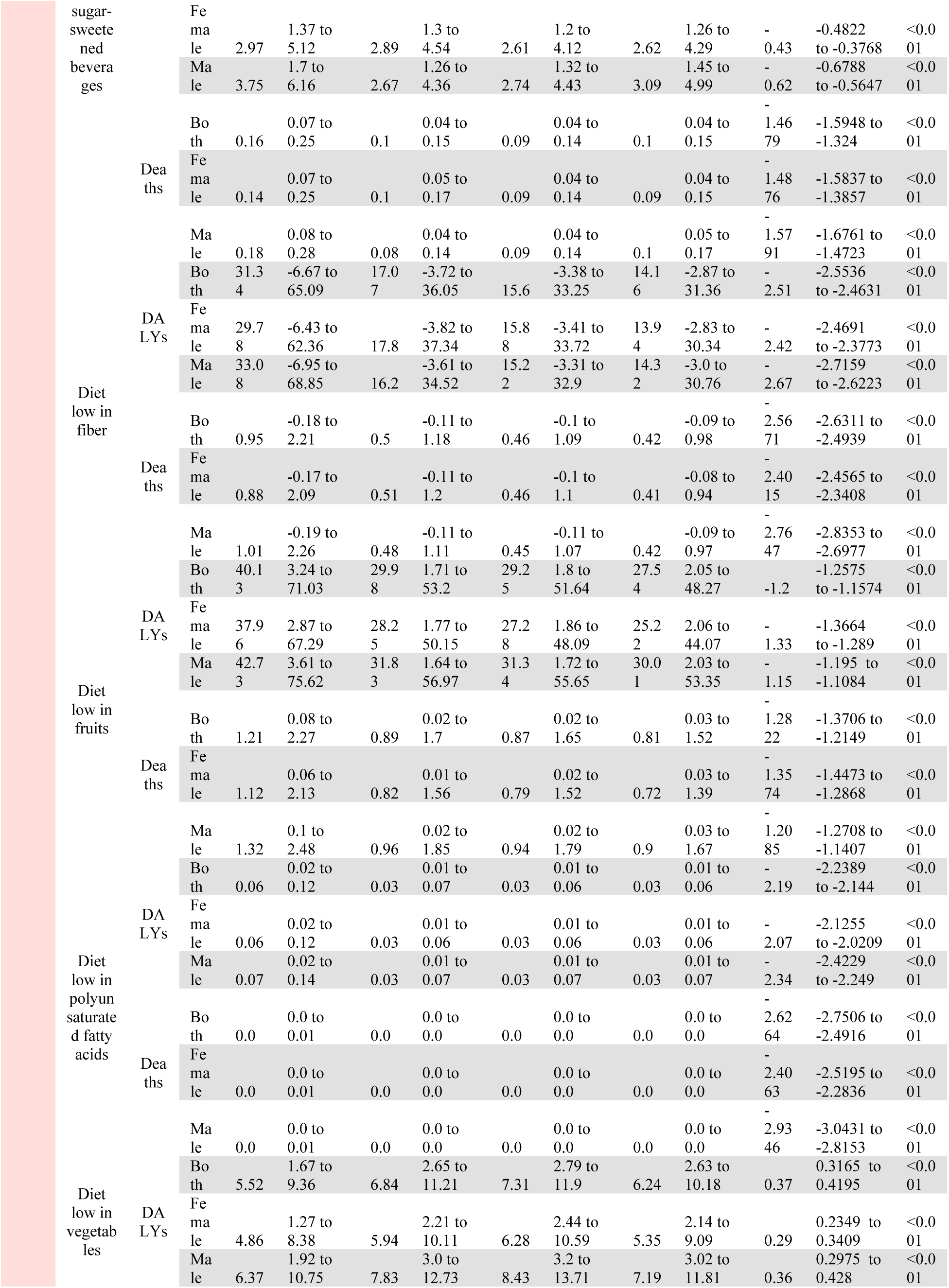
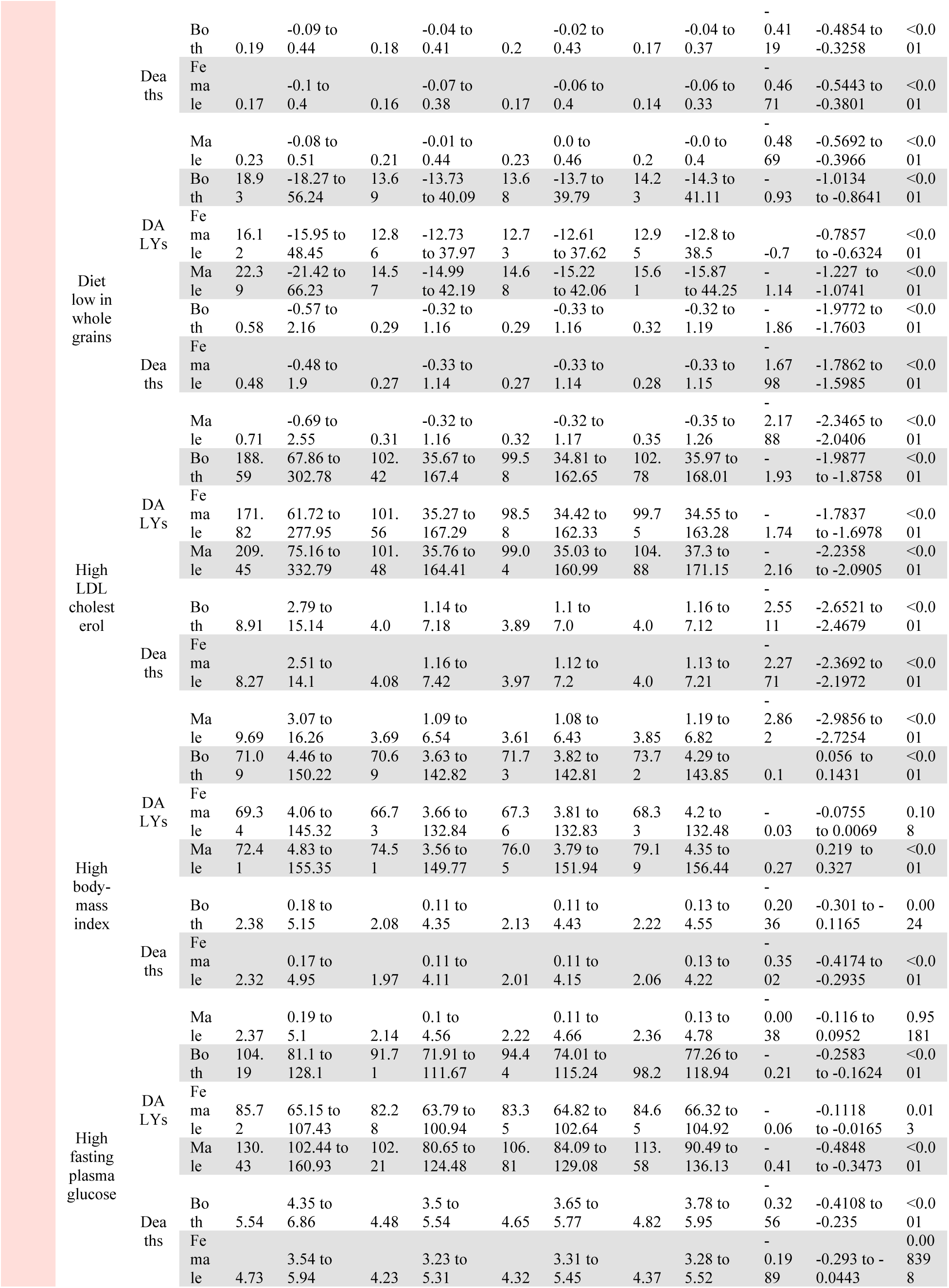
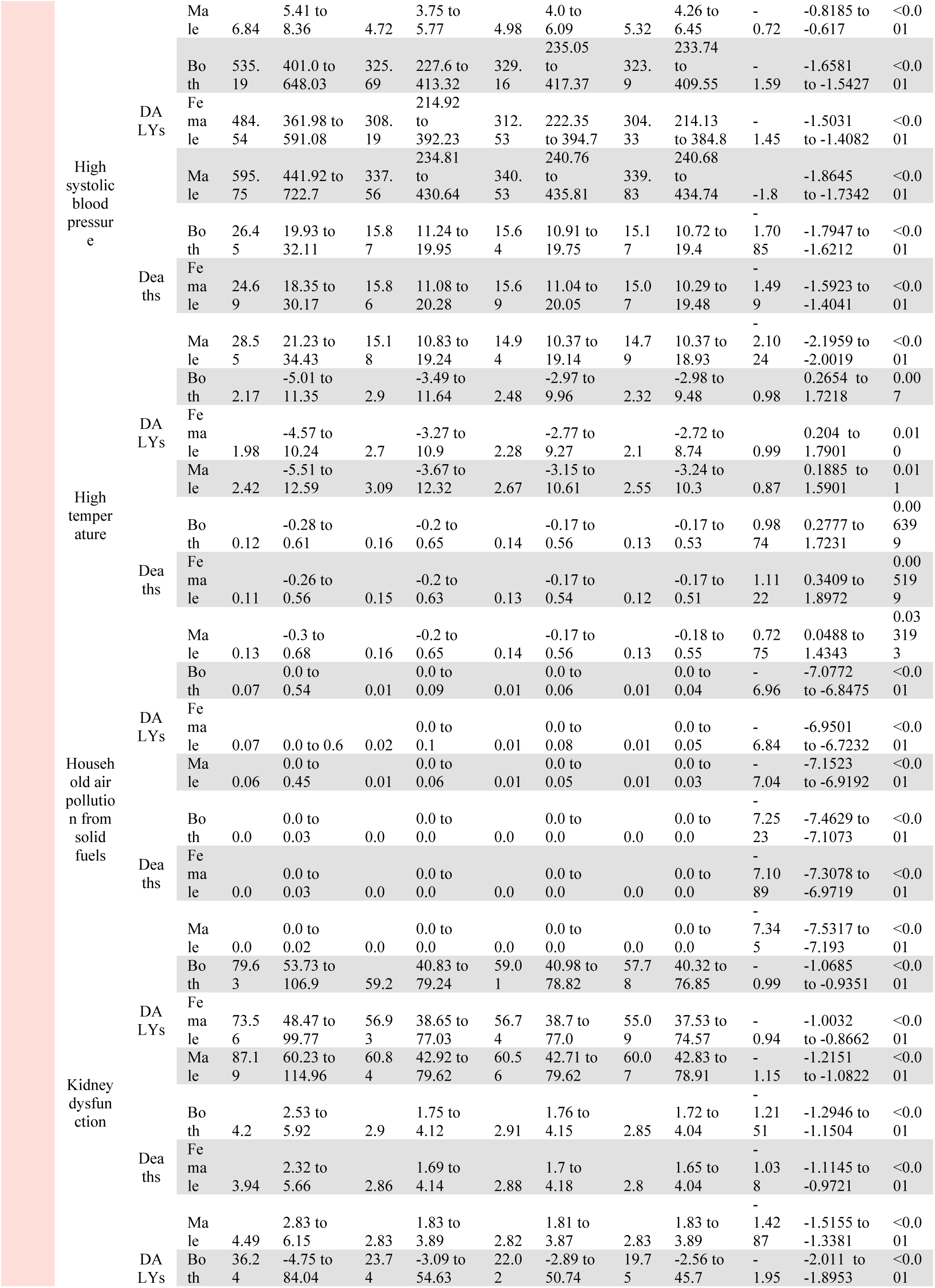
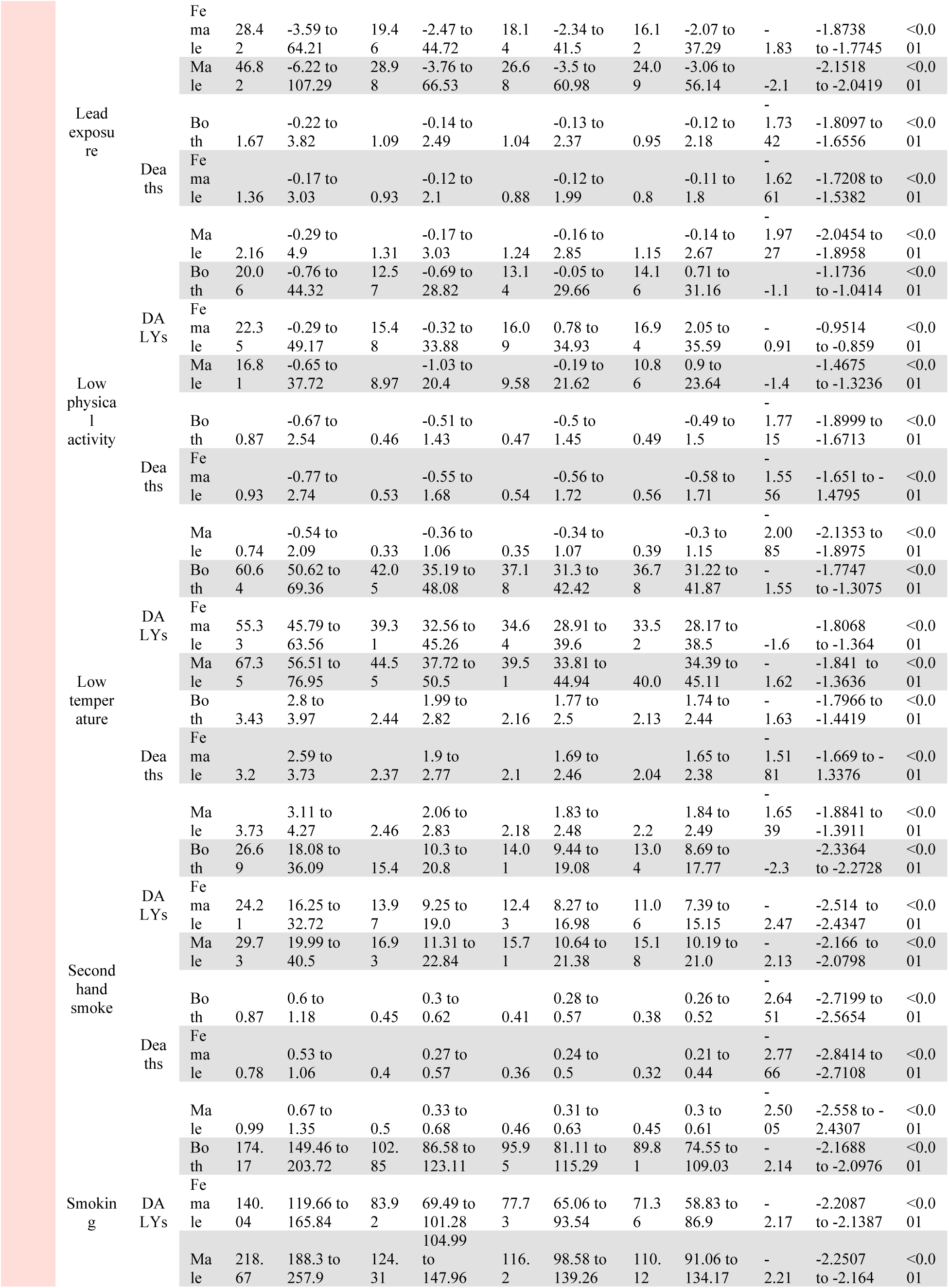
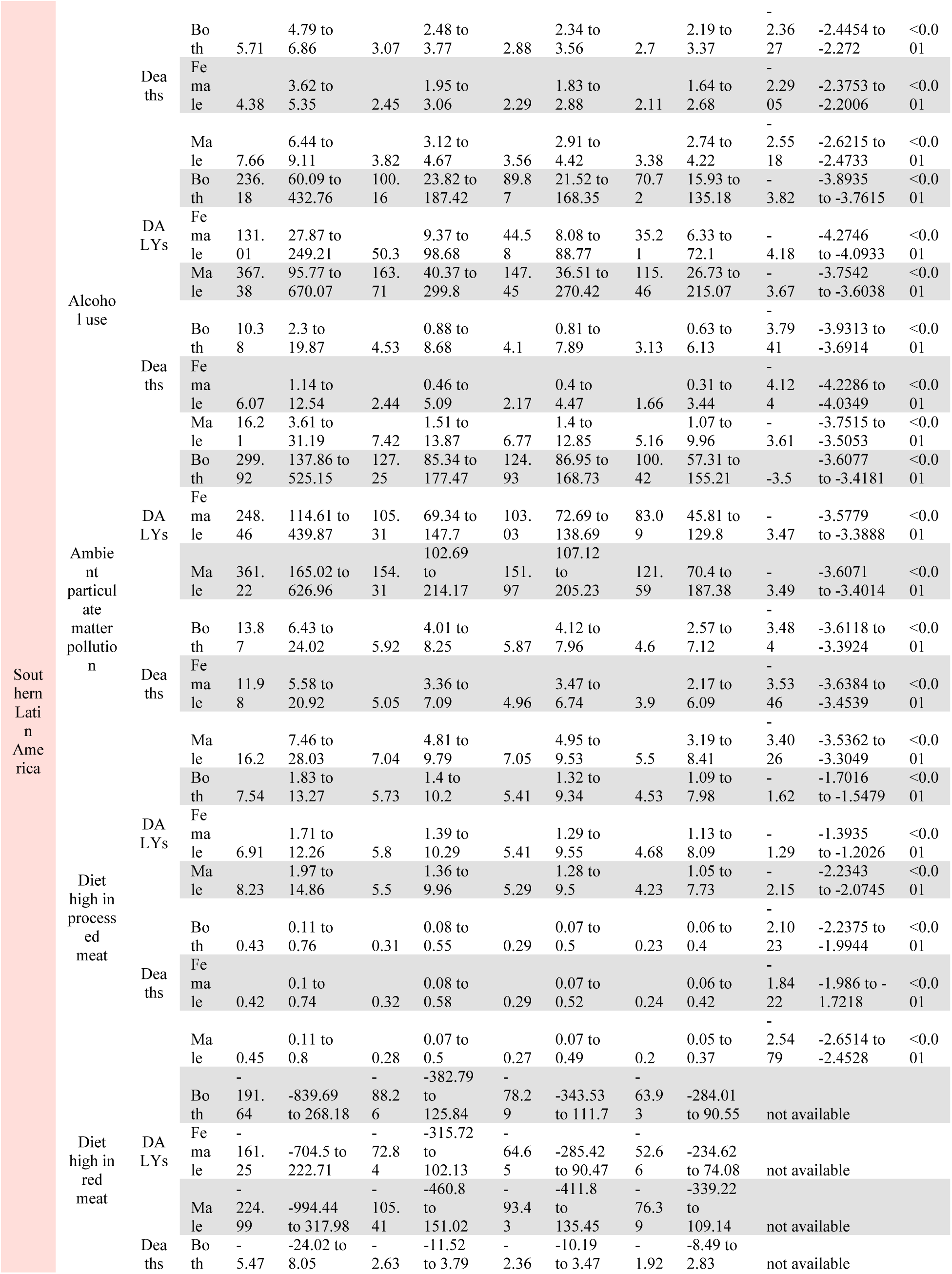
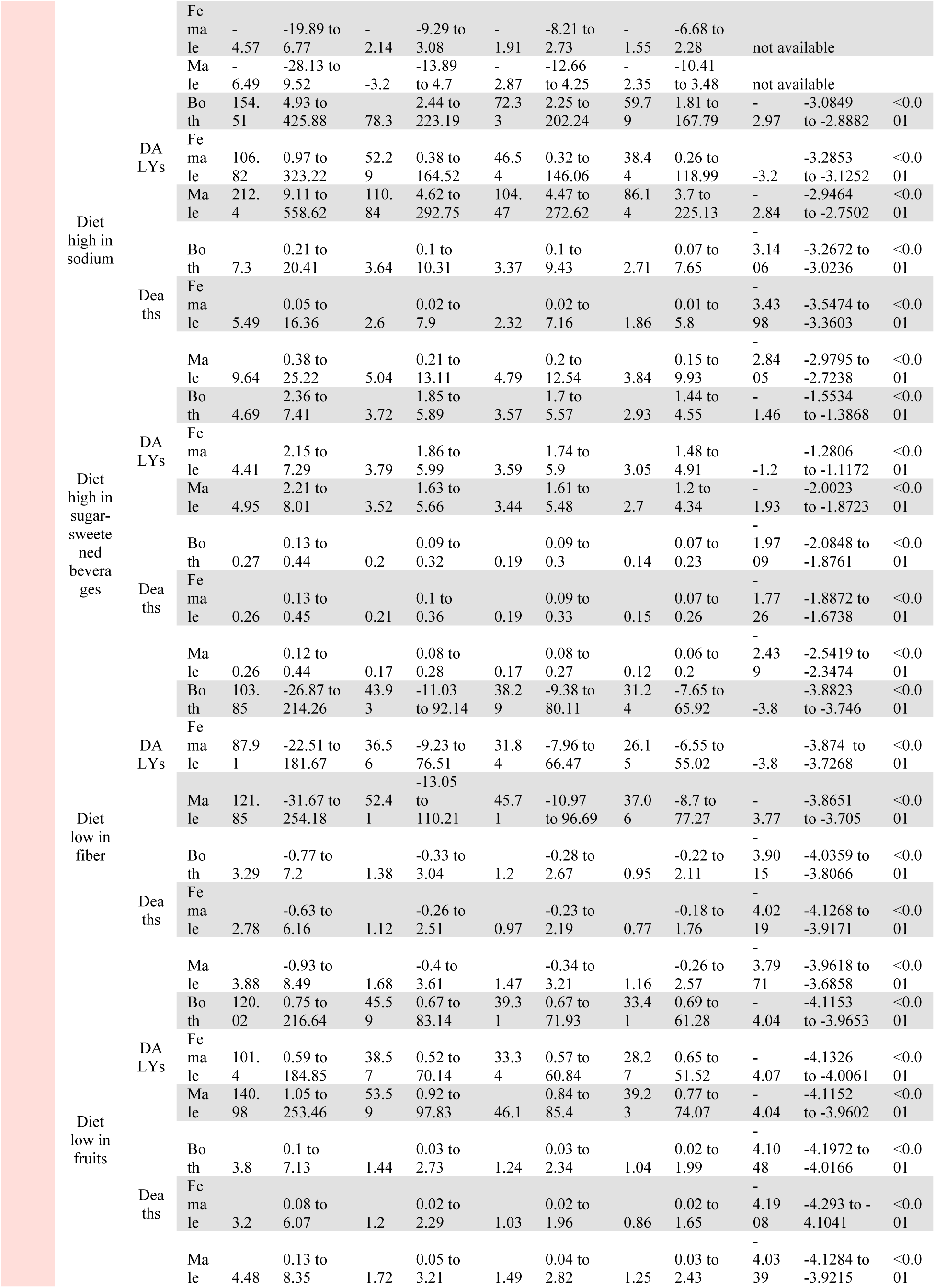
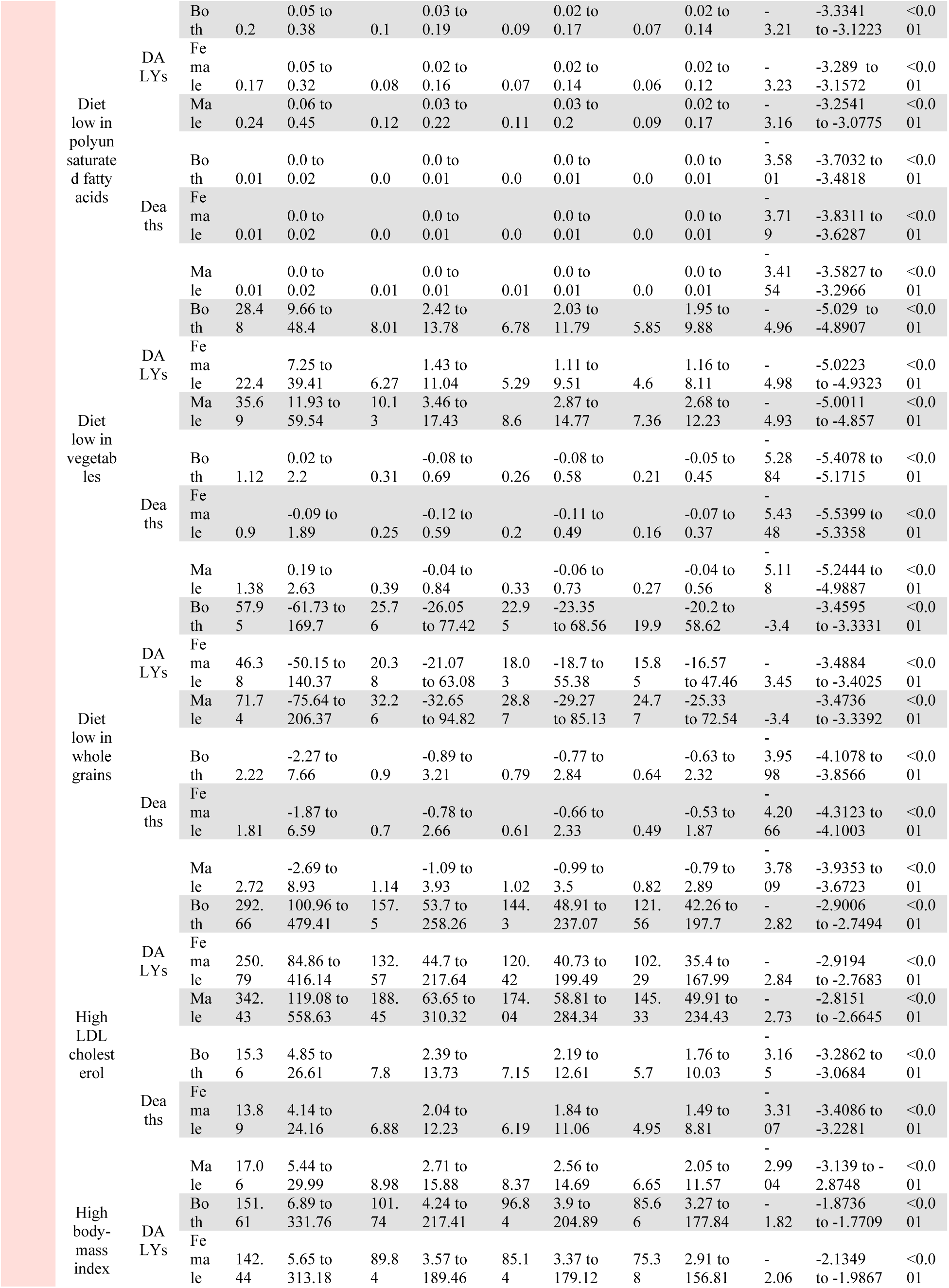
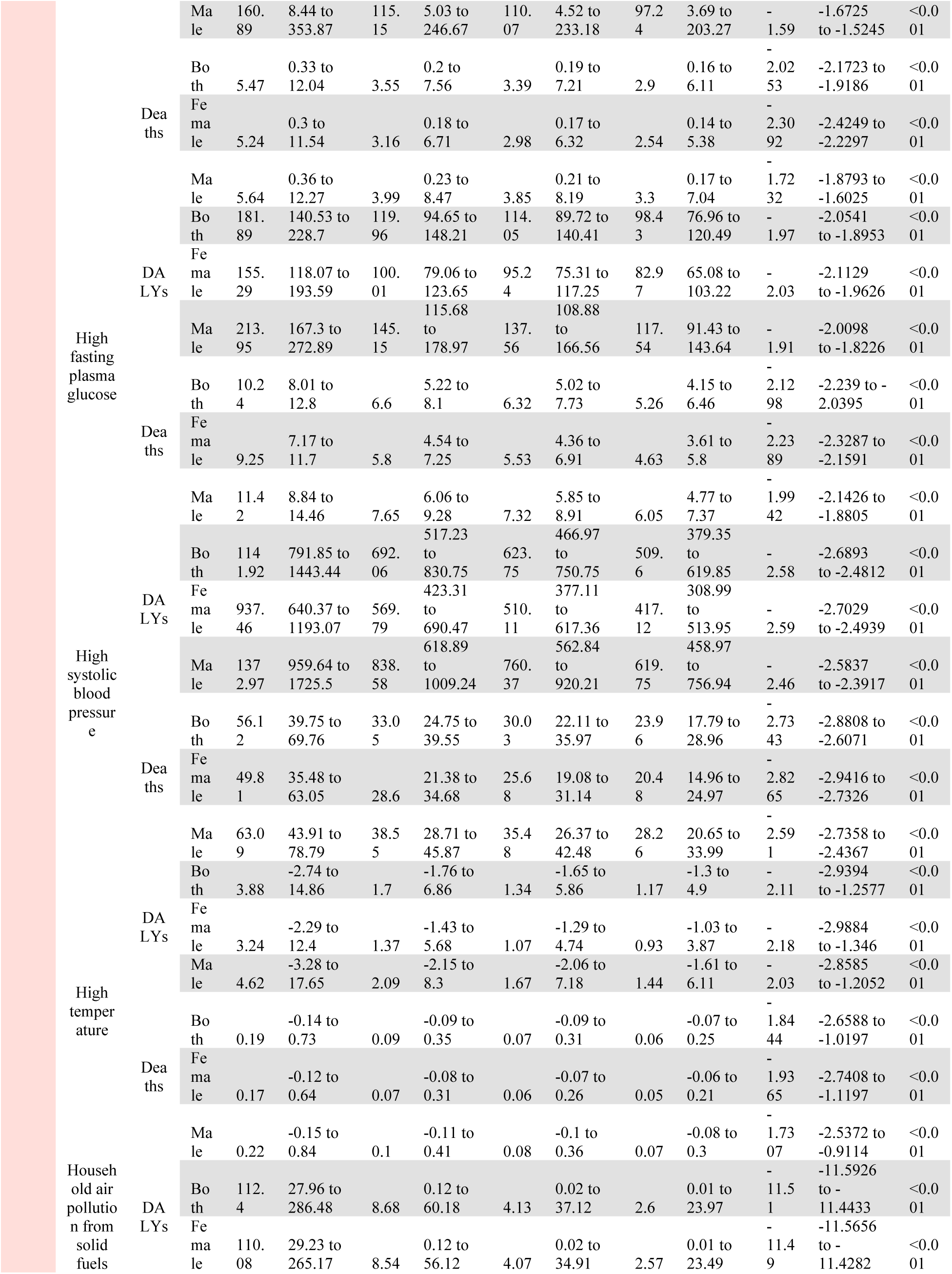
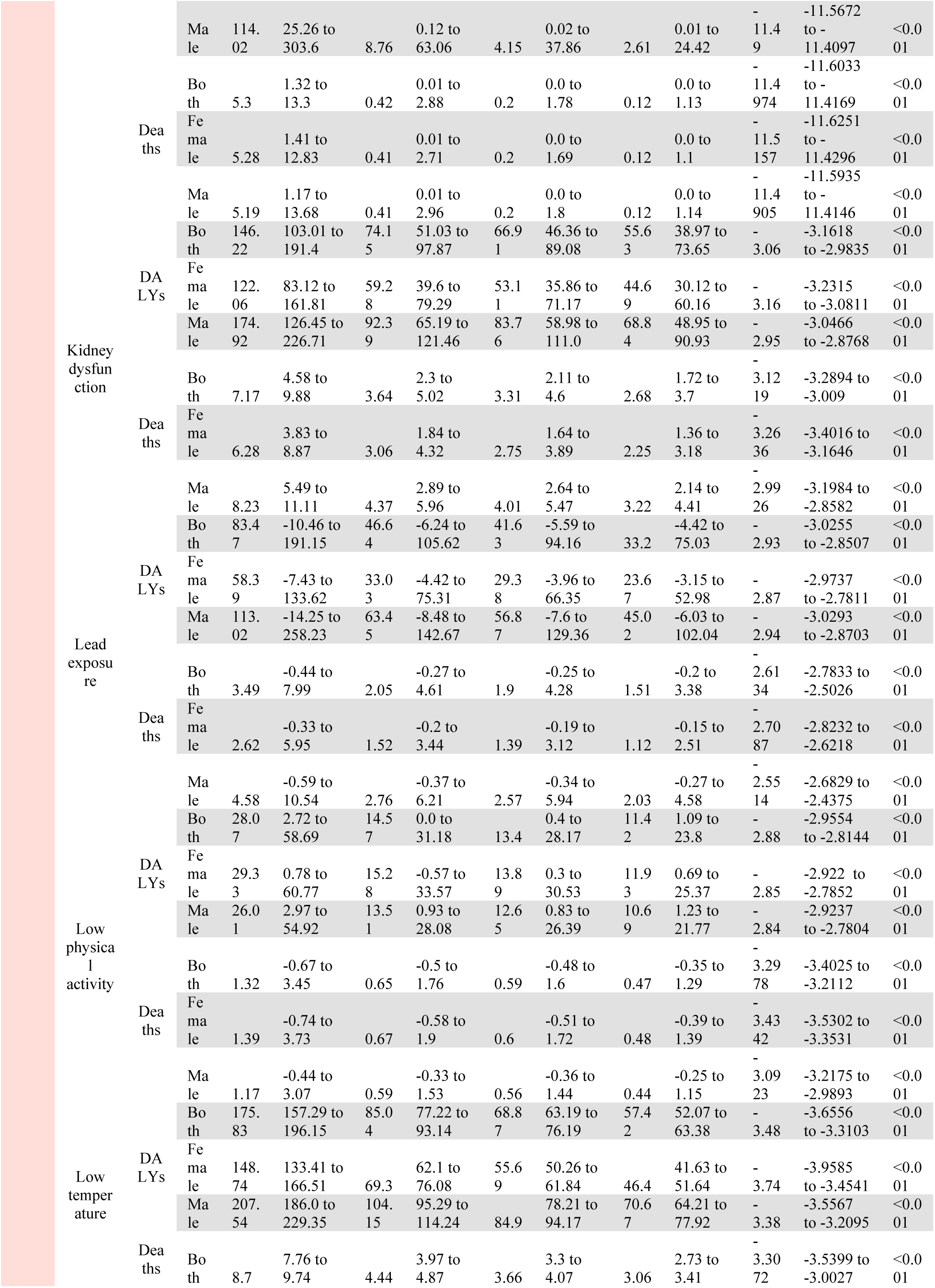
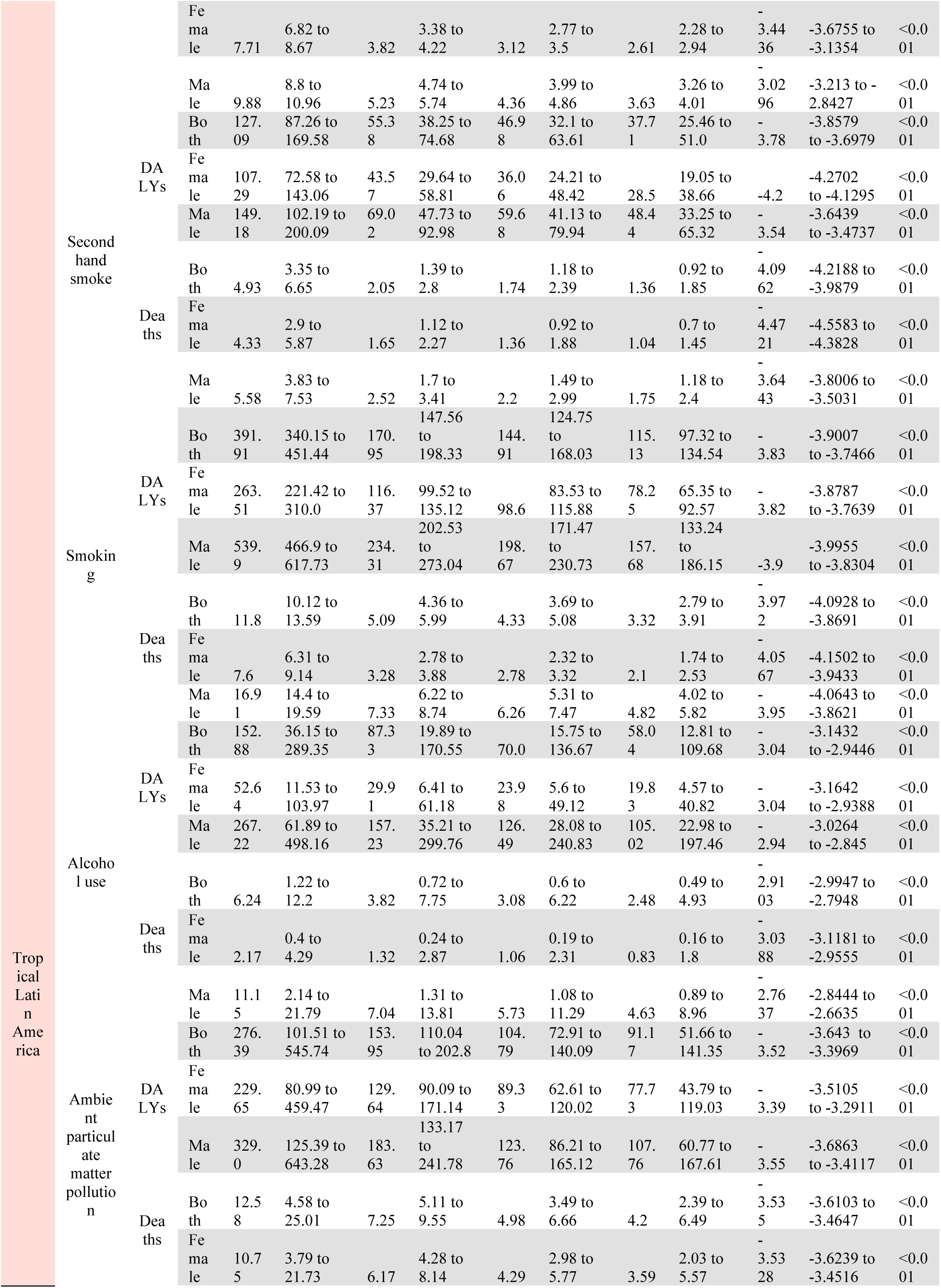
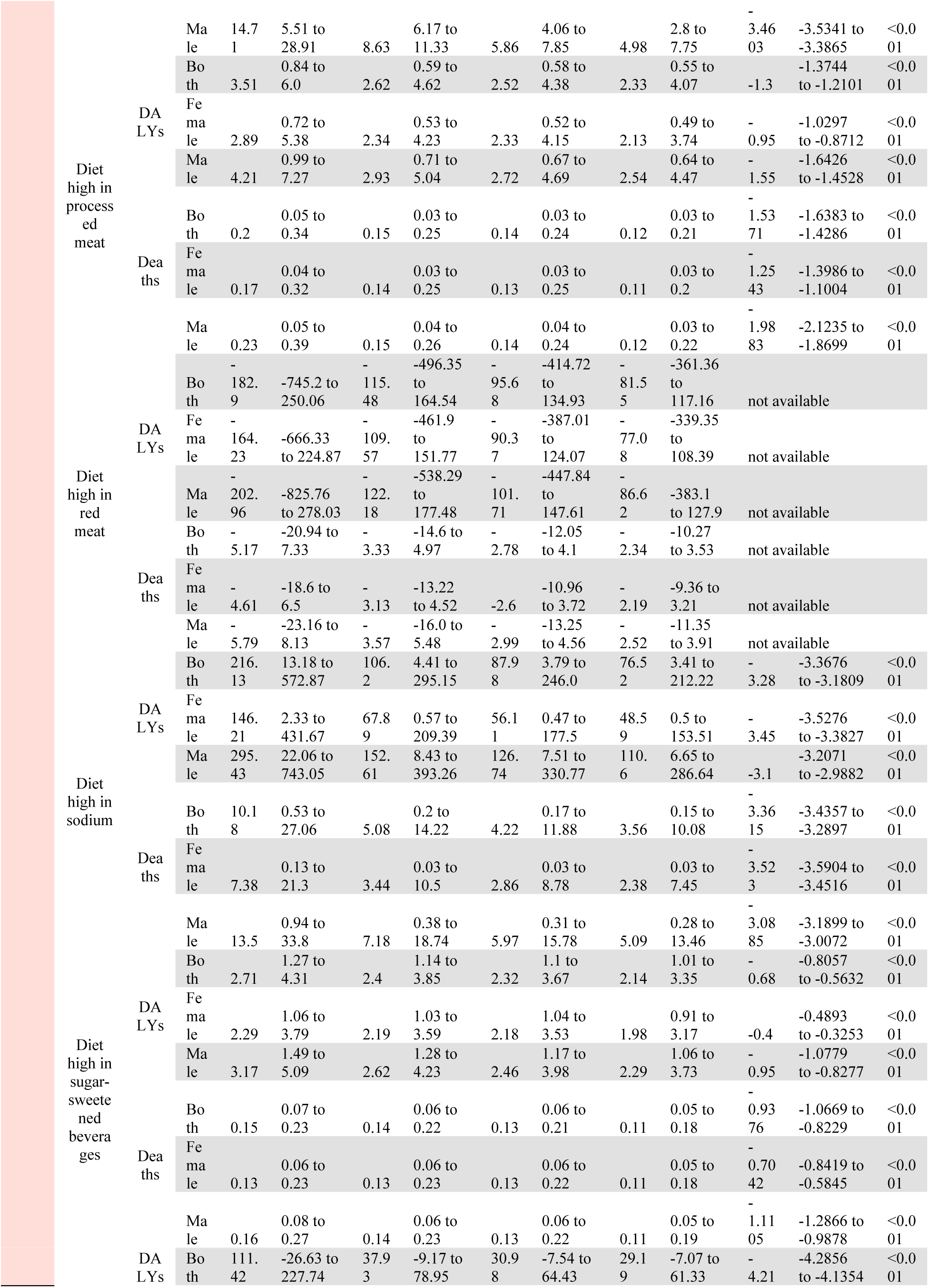
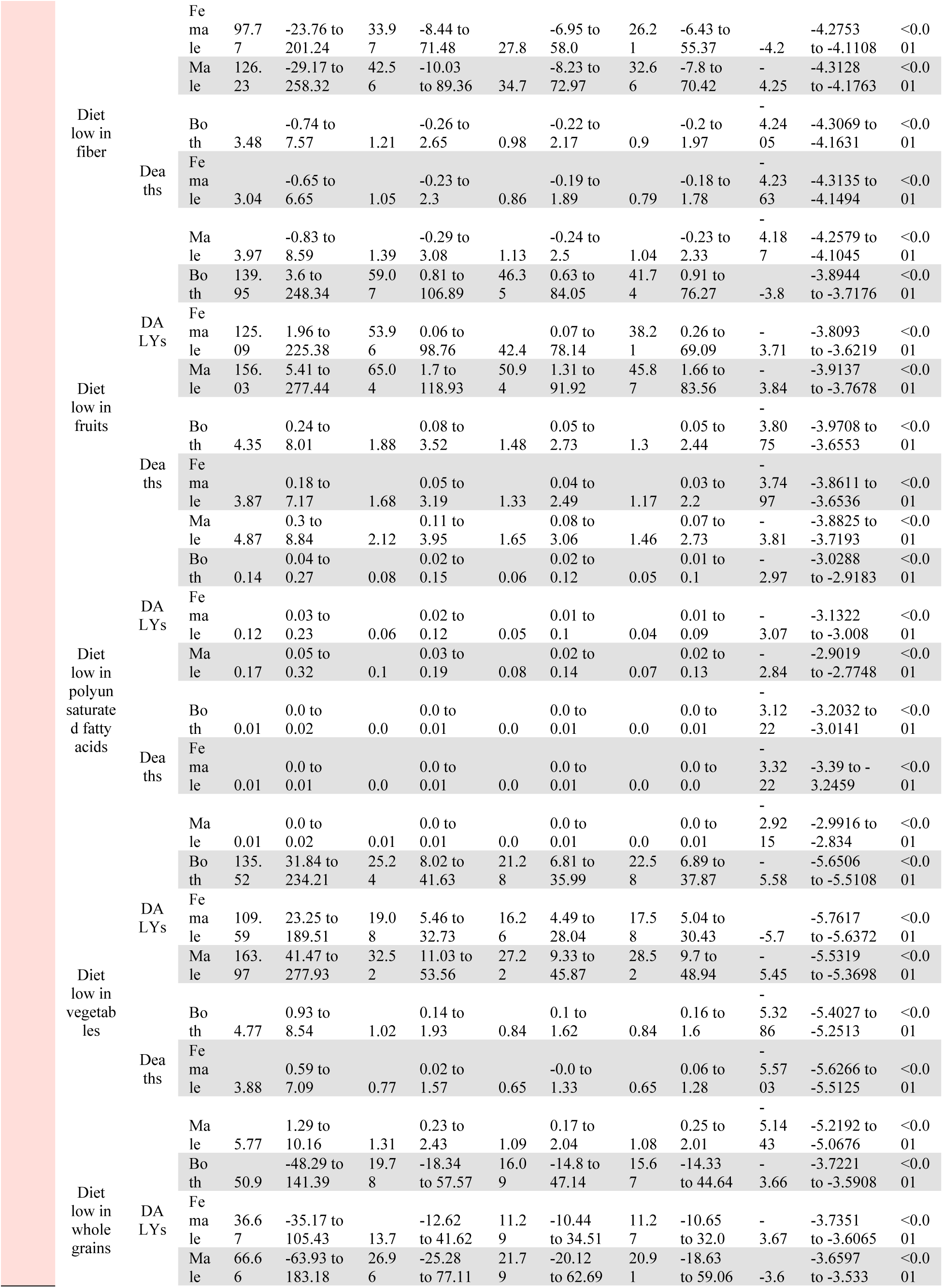
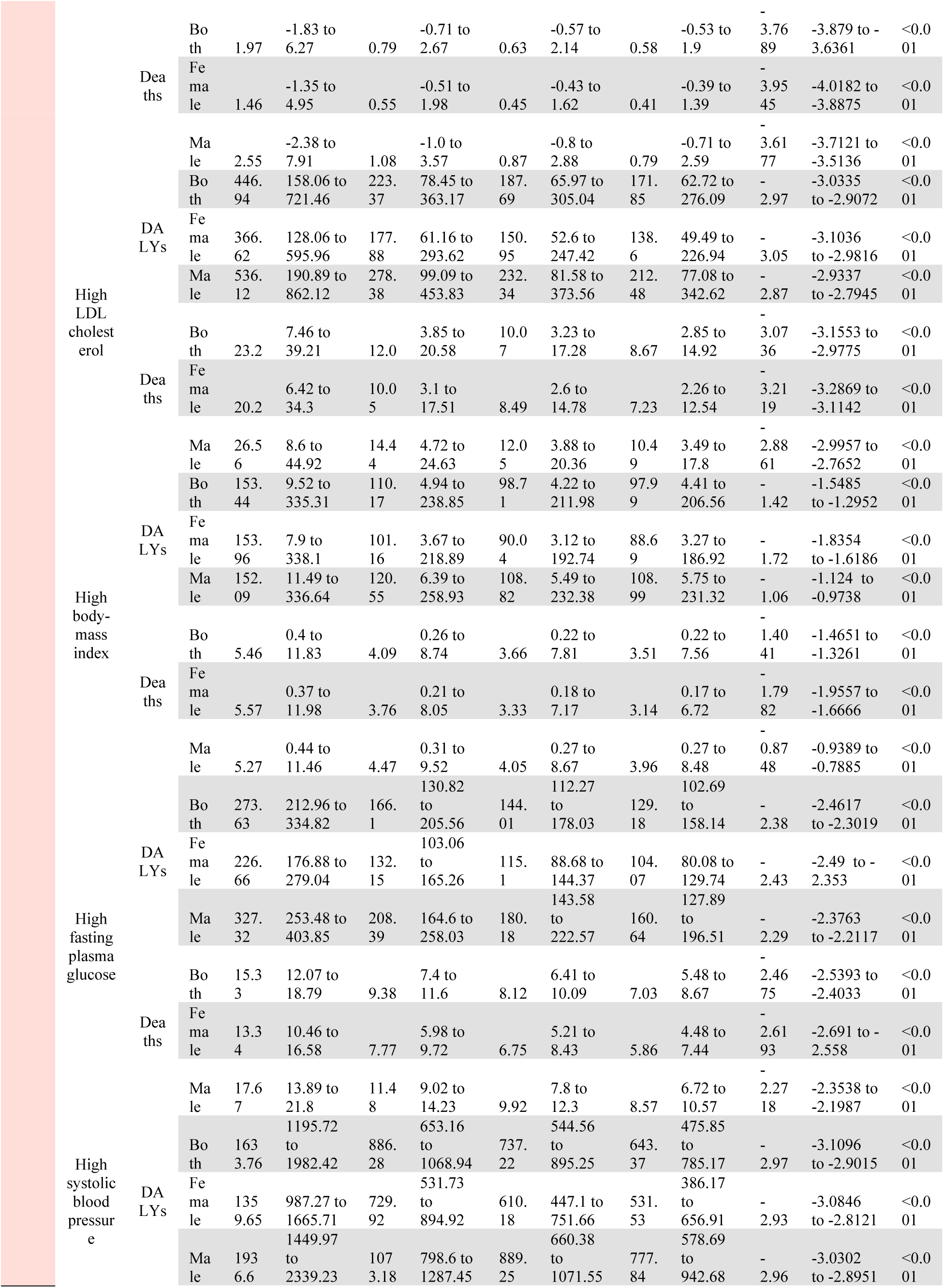
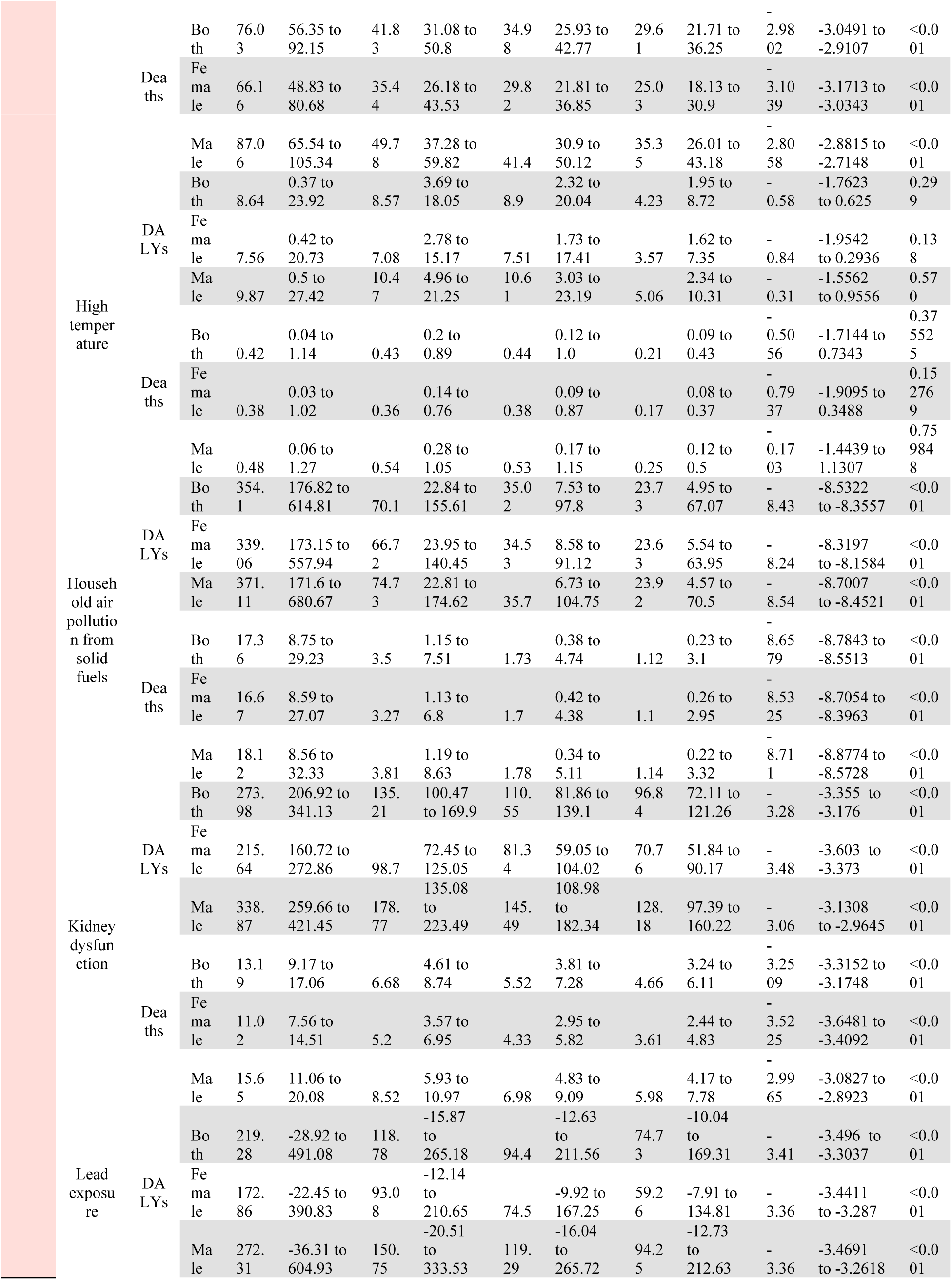
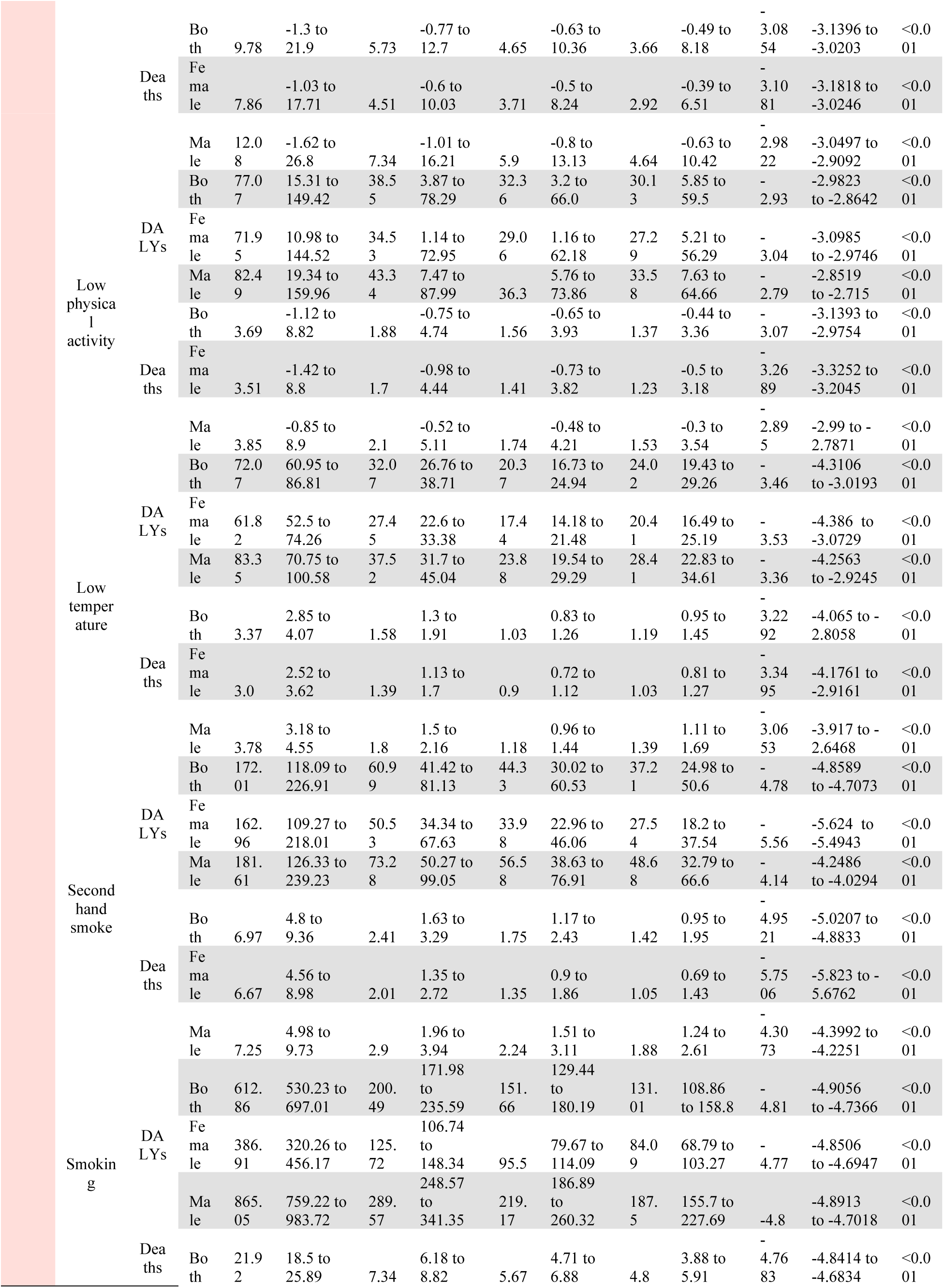
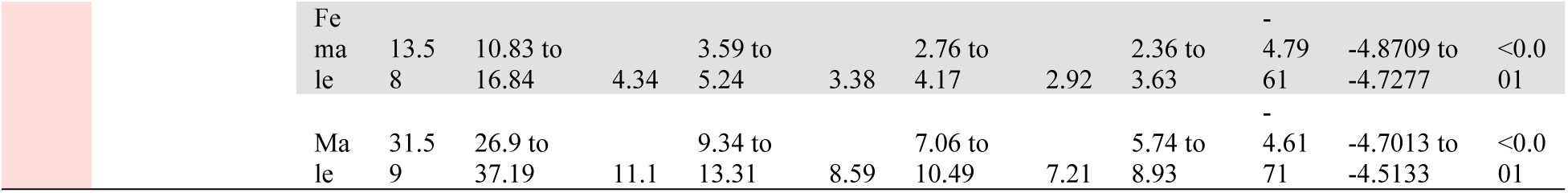
Trends in Stroke Burden Attributable to Risk Factors by Region in the Americas, 1990–2021. (Data are presented as rate per 100,000, both sexes, age-standardized, with 95% Uncertainty Intervals (UIs) in parentheses.) This table presents the temporal trends in stroke-related mortality and Disability-Adjusted Life Year (DALY) rates per 100,000 population in the Americas from 1990 to 2021, stratified by risk factors and sex. Data include age-standardized estimates for stroke burden at four time points (1990, 2010, 2015, and 2021) along with the Annual Average Percentage Change (AAPC) from 1990–2021 and its corresponding 95% Confidence Interval (CI). Negative AAPC values indicate a declining trend, whereas positive values indicate an increasing burden over time. AAPC p- values indicate statistical significance. CI = Confidence Interval; UI = Uncertainty Interval.

#### 2.2. Subregional and Country-Specific Trends

Andean Latin America showed moderate declines, with the largest reduction observed for household air pollution from solid fuels (DALYs AAPC −7.02 [95% CI −7.18 to −6.88]; p<0.001), while high temperature contributed to an increase in deaths (AAPC 3.5 [95% CI 1.9 to 5.12]; p<0.001). The Caribbean experienced the smallest overall declines with notable sex depairtities. In the Caribbean, the stroke burden attributable to high body-mass index increased substantially more in males than in females. While male DALYs rose by an AAPC of 1.72 (95% CI: 1.62 to 1.87), female DALYs increased by only 0.45 (95% CI: 0.36 to 0.53). Similar patterns were seen for stroke deaths, with AAPCs of 1.57 (95% CI: 1.47 to 1.67) in males versus 0.36 (95% CI: 0.26 to 0.45) in females. Despite an overall rise in DALYs due to high consumption of sugar-sweetened beverages (AAPC 0.55 [95% CI: 0.45 to 0.66]), stroke deaths attributable to this risk declined in females (AAPC −0.22 [95% CI: −0.33 to −0.11]) while rising in males (AAPC 0.62 [95% CI: 0.50 to 0.74]). In Central Latin America, declines were driven by improvements in dietary risks, including diet low in fiber (AAPC, deaths −2.25 [95% CI −2.31 to −2.17]; p<0.001) and diet low in whole grains (AAPC, deaths −2.32 [95% CI −2.39 to −2.25]; p<0.001). Reductions in high-income North America were observed for household air pollution from solid fuels (AAPC, deaths −7.25 [95% CI −7.46 to −7.1]; p<0.001), while increases were noted for high temperature (AAPC, deaths 0.99 [95% CI 0.28 to 1.72]; p=0.011) and diet low in vegetables (AAPC 0.41 [95% CI 0.49 to 0.33]; p<0.001). Southern and Tropical Latin America recorded the largest declines, primarily driven by reductions in household air pollution from solid fuels and diet low in fiber; Tropical Latin America had also significant reductions in household air pollution from solid fuels and diet low in vegetables (Table 3) (Supplement Figure 52, 53).

At the country level, high temperature (Jamaica, Dominican Republic, Panama) and high body-mass index (Haiti, Dominican Republic) were major drivers of increased stroke deaths/DALYs. Countries like Trinidad and Tobago, Grenada, and Chile showed the most pronounced reductions, predominantly due to large falls in household air pollution (AAPCs <−12%). (Supplementary Table 8) (Supplement Figure 54, 55)

#### 2.3. Sex-Stratified Temporal Patterns

Both females and males experienced decreasing stroke-related deaths and DALYs overall, although the magnitude varied by risk factor. Males had steeper declines in risks attributable to alcohol use (deaths: AAPC −1.78% [95% CI −1.83 to −1.73]; p<0.001 vs −1.46% [95% CI −1.50 to −1.43]; p<0.001 in females) and high fasting plasma glucose (males: −1.21% [95% CI −1.29 to −1.14] vs females: −0.99% [95% CI −1.04 to −0.95]; all p<0.001), while females showed slightly larger reductions in risks attributable to diet high in sodium and high body-mass index.

Regionally, these sex differences were most pronounced in Central Latin America (where risk declined more in females), whereas the Caribbean exhibited smaller overall improvements (Table 3).

### 3. Risk Factors with Notable Increases, Minimal Changes, or Significant Declines

#### 3.1. Emerging or Growing Risks

High temperature consistently drove increases in stroke-related deaths and DALYs in several countries (e.g., Jamaica, Dominican Republic, and Panama). High body-mass index also rose notably in Haiti and the Dominican Republic. Certain dietary factors (e.g., sugar-sweetened beverages) increased in low- and middle-SDI settings. (Supplementary Table 8)

#### 3.2. Persistent or Minimal Changes

Several countries exhibited negligible variation in risk factor trends. For instance, alcohol use in the Dominican Republic (AAPC near zero), ambient particulate matter in Belize, and certain dietary factors (e.g., low vegetables in the Bahamas) remained unchanged over time. (Supplementary Table 8)

#### 3.3. Significant Declines

Household air pollution from solid fuels showed the most uniform and substantial decline across nearly all subregions and countries (e.g., Trinidad and Tobago, Grenada, Chile, Saint Kitts and Nevis), with AAPCs often greater than 10%. Smoking also decreased significantly, albeit to varying degrees across the Americas. (Supplementary Table 8)

## Discussion

This analysis revealed a decline in stroke-related deaths and DALYs attributable to risk factors in the Americas between 1990 and 2021, with distinct epidemiological patterns that were strongly influenced by SDI levels and varied across geographical regions and sexes. High systolic blood pressure was consistently the leading risk factor for stroke-related deaths and DALYs attributable to risk factors, contributing to 55% of all stroke deaths and 52% of all stroke DALYs regionwide, and with higher proportions in some subregions—particularly in lower SDI countries. Similarly, other metabolic factors such as high fasting plasma glucose and high LDL cholesterol, and certain behavioral and dietary risk factors such as smoking, alcohol, high sodium intake also shaped these outcomes considerably. Our findings align with global trends based on 2021 GBD data, where high systolic blood pressure emerged as a primary stroke risk factor, corroborating prior findings for Latin America and the Caribbean region from the GBD 2019 study.^1,4^

High SDI regions often have systems in place that allow for rapid response and timely initiation of treatment, resulting in a notably shorter time to diagnosis and treatment compared to low and lower-middle-SDI countries.^17,18^ These countries also have greater access to preventive care and effective acute stroke interventions, allowing more people to survive with stroke-related disabilities. By contrast, low SDI and low-middle SDI countries have higher death rates likely attributed to poor access and quality of preventive care as well as delays in diagnosis and limited access to timely medical intervention.^17^ For instance, in the Americas countries with the highest levels of population hypertension control tend to have low stroke mortality rates.^6^ Thus, disparities in healthcare access are a key contributing factor to the differences observed in stroke outcomes across SDI levels.^19,20^

Our findings underscore a complex relationship between food consumption patterns and stroke burdens across different SDI context. Greater stroke deaths attributed to high consumption of processed meat was noted across SDIs, but the impact on DALY outcomes was variable in more affluent regions, suggesting differential health impacts and possible variations in disease rehabilitation and lifestyle modifications post-stroke. This observation invites further exploration into local dietary habits and their protective health implications particularly for stroke.

Sex-based disparities were observed in death- and DALYs-related stroke across the Americas, with men bearing a disproportionately higher impact from key risk factors such as high systolic blood pressure, high fasting plasma glucose, smoking, and alcohol. These disparities were more pronounced in some regions. For instance, in Southern Latin America, the DALY rate from high systolic blood pressure was significantly higher in males than in females.

Our findings are consistent with long-term trends observed in previous reports, reflecting a global reduction in stroke mortality and DALYs.^21^ ^22^ This reduction has been more pronounced in males, who showed a slightly faster annual decrease in stroke-related mortality and DALYs as compared to females. Multiple factors, including age, stroke type, comorbidities, and access to care may influence this observed difference in trends between sexes. A meta-analysis that included 36 studies and over 8 million subjects demonstrated higher mortality risk in women during the in-hospital period due to stroke of unspecified type, and highlighted an unadjusted estimate of 10% higher mortality post ischemic stroke in females at all follow-up points.^23^ Furthermore, historical data, such as those from the International Stroke Trial, suggest a 5.2% higher raw mortality in females at six-months, though mortality was lower among females (odds ratio 0.90; 95% CI 0.83–0.98) after adjusting for potential confounders such as age, stroke severity, atrial fibrillation, and high systolic blood pressure^24^. Consequently, women with vascular risk factors should be a focus of future stroke prevention strategies.

Interestingly, our analysis indicated minimal change in stroke-related outcomes associated with some behavioral factors, such as alcohol use, high dietary sodium intake, and high body mass index, even in high-income North American regions where the temporal changes did not exceed a modest -0.02% over nearly three decades. This surprising stagnation occurred despite extensive public health initiatives and policy shifts aimed at improving diet quality globally. The lack of progress may reflect entrenched eating habits, socio-economic constraints, or ineffectiveness in current health policy implementations. Our findings underscore the need for more pragmatic, tailored, and culturally adapted strategies to achieve meaningful changes in these regions.

Although this study provides a comprehensive and longitudinal analysis of stroke burden across the Americas and offers policymakers robust, comparable data that enhance the understanding of regional stroke epidemiology and support the development of targeted prevention strategies, the study also tampered by some limitations. The reliance in the GBD data on mixed data sources, including hospital records, surveys, and verbal autopsies, introduces variability in data quality, particularly in low SDI regions, which may lead to underestimating the real effect of risk factors and stroke association. The methodological challenges of attributing stroke risks to specific factors when multiple risk factors coexist (e.g., hypertension and dyslipidemia) add to the uncertainty of our estimates. Additionally, the ecological design of our analysis focuses on population-level trends and does not account for individual-level causation or clinical practice variations.

Despite progress in the region, this study highlights that efforts to prevent and reduce the burden of stroke remain insufficient and uneven across countries. However, there are clear opportunities. For example, 33 countries in the region are implementing HEARTS, the regional adaptation of WHO Global HEARTS, coordinated by the Pan American Health Organization (PAHO). The program currently reaches 34.2 million adults, with 4.5 million receiving hypertension treatment across over 6,500 primary healthcare centers. HEARTS aims to transform health services and clinical practice to improve hypertension control and integrated cardiovascular disease risk management in primary healthcare.^25^ Additionally, the WHO, in collaboration with the World Stroke Organization and others, has published the “Framework for the Care of Acute Coronary Syndrome and Stroke”. This framework outlines a health systems approach, covering care models, service packages, planning, and the role of public health authorities, emphasizing the importance of a continuum of care—from community awareness and prehospital care to acute hospital care, rehabilitation, and ongoing management.^26^

## Conclusions

The overall burden of stroke in the Americas attributable to modifiable risk factors decreased between 1990 and 2021. However, significant disparities by sex and socioeconomic status persist, disproportionately affecting countries with lower SDI. High systolic blood pressure remains the leading modifiable risk factor, responsible for more than half of all stroke-related deaths in some subregions, followed by high fasting plasma glucose and high LDL cholesterol. Minimal progress has been made in improving stroke- related outcomes associated with alcohol use, high dietary sodium intake, and high body mass index, even in high-income countries. These findings underscore the urgent need to prioritize and improve the reach and quality of public health and health system interventions aimed at the prevention and effective management of traditional risk factors, especially hypertension and other cardiometabolic conditions.

## Data Availability

All data used in this study are publicly available from the Global Burden of Disease (GBD) 2021 study. The datasets can be accessed through the Institute for Health Metrics and Evaluation (IHME) Global Health Data Exchange (GHDx) at http://ghdx.healthdata.org/gbd-results-tool.

## Disclaimer

RM, PO and AR are staff members of the Pan American Health Organization. The authors alone are responsible for the views expressed in this publication, and they do not necessarily represent the decisions or policies of the Pan American Health Organization.

## Funding

None

to address disparities and socio-economic factors influencing health outcomes in the Americas.

## References

1 Feigin VL, Abate MD, Abate YH, et al. Global, regional, and national burden of stroke and its risk factors, 1990–2021: a systematic analysis for the Global Burden of Disease Study 2021. The Lancet Neurology 2024; 23: 973–1003.

2 Martínez R, Venturelli PM, Ordunez P, et al. Burden of Stroke Across 38 Countries and Territories of the Americas from 1990 to 2021: A Population-Based Trends Analysis from the Global Burden of Disease Study 2021. 2024; published online Aug 21. DOI:10.2139/ssrn.4929800.

3 GBD 2019 Stroke Collaborators. Global, regional, and national burden of stroke and its risk factors, 1990-2019: a systematic analysis for the Global Burden of Disease Study 2019. Lancet Neurol 2021; 20: 795–820.

4 Pacheco-Barrios K, Giannoni-Luza S, Navarro-Flores A, et al. Burden of Stroke and Population-Attributable Fractions of Risk Factors in Latin America and the Caribbean. Journal of the American Heart Association 2022; 11: e027044.

5 Fan J, Ma W, Liu J, et al. Associations between socioeconomic status and stroke in American adults: A population-based study. Preventive Medicine Reports 2023; 35: 102354.

6 Martinez R, Soliz P, Campbell NRC, Lackland DT, Whelton PK, Ordunez P. Association between population hypertension control and ischemic heart disease and stroke mortality in 36 countries of the Americas, 1990-2019: an ecological study. Rev Panam Salud Publica 2022; 46: e143.

7. 7 Institute For Health Metrics And Evaluation. Global Burden of Disease Study 2021 (GBD 2021) Cause, REI, and Location Hierarchies. 2024. DOI:10.6069/G44H-ZC04.

8. 8 Global Burden of Disease (GBD) data and tools guide | Institute for Health Metrics and Evaluation. https://www.healthdata.org/research-analysis/about-gbd/gbd-data-and-tools-guide (accessed Nov 12, 2024).

9 Murray CJ, Ezzati M, Flaxman AD, et al. GBD 2010: design, definitions, and metrics. The Lancet 2012; 380: 2063–6.

10 GBD 2021 Risk Factors Collaborators. Global burden and strength of evidence for 88 risk factors in 204 countries and 811 subnational locations, 1990-2021: a systematic analysis for the Global Burden of Disease Study 2021. Lancet 2024; 403: 2162–203.

11 GBD 2019 Risk Factors Collaborators. Global burden of 87 risk factors in 204 countries and territories, 1990-2019: a systematic analysis for the Global Burden of Disease Study 2019. Lancet 2020; 396: 1223–49.

12 GBD 2021 Diseases and Injuries Collaborators. Global incidence, prevalence, years lived with disability (YLDs), disability-adjusted life-years (DALYs), and healthy life expectancy (HALE) for 371 diseases and injuries in 204 countries and territories and 811 subnational locations, 1990-2021: a systematic analysis for the Global Burden of Disease Study 2021. Lancet 2024; 403: 2133–61.

13 Aho K, Harmsen P, Hatano S, Marquardsen J, Smirnov VE, Strasser T. Cerebrovascular disease in the community: results of a WHO collaborative study. Bull World Health Organ 1980; 58: 113–30.

14. 14 Global Burden of Disease 2021 risk factor factsheets | Institute for Health Metrics and Evaluation. https://www.healthdata.org/research-analysis/diseases-injuries/factsheets-overview/about-risk-factor (accessed March 2, 2025).

15 Martinez R, Muñoz-Venturelli P, Ordunez P, et al. Risk and impact of stroke across 38 countries and territories of the Americas from 1990 to 2021: a population-based trends analysis from the Global Burden of Disease Study 2021. The Lancet Regional Health – Americas 2025; 43. DOI:10.1016/j.lana.2025.101017.

16. Global Burden of Disease Study 2021 (GBD 2021) Socio-Demographic Index (SDI) 1950– 2021 | GHDx. https://ghdx.healthdata.org/record/global-burden-disease-study-2021-gbd-2021-socio-demographic-index-sdi-1950%E2%80%932021 (accessed Jan 21, 2025).

17 Aguirre AO, Rogers JL, Reardon T, et al. Stroke management and outcomes in low-income and lower-middle-income countries: a meta-analysis of 8535 patients. 2023; published online March 31. DOI:10.3171/2023.2.JNS222807.

18 Lees KR, Bluhmki E, von Kummer R, et al. Time to treatment with intravenous alteplase and outcome in stroke: an updated pooled analysis of ECASS, ATLANTIS, NINDS, and EPITHET trials. Lancet 2010; 375: 1695–703.

19 Feigin VL, Lawes CMM, Bennett DA, Barker-Collo SL, Parag V. Worldwide stroke incidence and early case fatality reported in 56 population-based studies: a systematic review. Lancet Neurol 2009; 8: 355–69.

20 Strong K, Mathers C, Bonita R. Preventing stroke: saving lives around the world. Lancet Neurol 2007; 6: 182–7.

21 Araujo ACB, de Souza OF, de Alencar FEC, et al. Trends in Mortality Due to Stroke in South America between 1990 and 2019. Epidemiologia 2024; 5: 581–91.

22 He Q, Wang W, Zhang Y, et al. Global, Regional, and National Burden of Stroke, 1990–2021: A Systematic Analysis for Global Burden of Disease 2021. Stroke; 0. DOI:10.1161/STROKEAHA.124.048033.

23 Abdel-Fattah A-R, Pana TA, Smith TO, et al. Gender differences in mortality of hospitalised stroke patients. Systematic review and meta-analysis. Clinical Neurology and Neurosurgery 2022; 220: 107359.

24 Niewada M, Kobayashi A, Sandercock PAG, Kamiński B, Członkowska A. Influence of Gender on Baseline Features and Clinical Outcomes among 17,370 Patients with Confirmed Ischaemic Stroke in the International Stroke Trial. Neuroepidemiology 2005; 24: 123–8.

25 Ordunez P, Campbell NRC, DiPette DJ, et al. HEARTS in the Americas: Targeting Health System Change to Improve Population Hypertension Control. Curr Hypertens Rep 2024; 26: 141–56.

26. 26 Organization WH. Framework for the care of acute coronary syndrome and stroke. World Health Organization, 2024 https://iris.who.int/handle/10665/380065 (accessed March 2, 2025).

